# Novel paradigm enables accurate monthly gestational screening to prevent congenital toxoplasmosis and more

**DOI:** 10.1101/2023.04.26.23289132

**Authors:** Y Zhou, K Leahy, A Grose, J Lykins, M Siddiqui, N. Leong, P Goodall, S Withers, K Ashi, S Schrantz, V Tesic, A P Abeleda, K Beavis, F Clouser, M Ismail, M Christmas, R Piarroux, D Limonne, E Chapey, S Abraham, I Baird, J Thibodeau, K Boyer, E Torres, S Conrey, K Wang, MA Staat, N Back, J Gomez Marin, F Peyron, S Houze, M Wallon, R McLeod

## Abstract

**Background:** Congenital toxoplasmosis is a treatable, preventable disease, but untreated causes death, prematurity, loss of sight, cognition and motor function, and substantial costs worldwide.

**Methods/Findings:** In our ongoing USA feasibility/efficacy clinical trial, data collated with other ongoing and earlier published results proved high performance of an Immunochromatographic-test(ICT) that enables accurate, rapid diagnosis/treatment, establishing new paradigms for care. Overall results from patient blood and/or serum samples tested with ICT compared with gold-standard-predicate-test results found ICT performance for 4606 sera/1876 blood, 99.3%/97.5% sensitive and 98.9%/99.7% specific. However, in the clinical trial the FDA-cleared-predicate test initially caused practical, costly problems due to false-positive-IgM results. For 58 persons, 3/43 seronegative and 2/15 chronically infected persons had false positive IgM predicate tests. This caused substantial anxiety, concerns, and required costly, delayed confirmation in reference centers. Absence of false positive ICT results contributes to solutions: Lyon and Paris France and USA Reference laboratories frequently receive sera with erroneously positive local laboratory IgM results impeding patient care. Therefore, thirty-two such sera referred to Lyon’s Reference laboratory were ICT-tested. We collated these with other earlier/ongoing results: 132 of 137 USA or French persons had false positive local laboratory IgM results identified correctly as negative by ICT. Five false positive ICT results in Tunisia and Marseille, France, emphasize need to confirm positive ICT results with Sabin-Feldman-Dye-test or western blot. Separate studies demonstrated high performance in detecting acute infections, meeting FDA, CLIA, WHO ASSURED, CEMark criteria and patient and physician satisfaction with monthly-gestational-ICT-screening.

**Conclusions/Significance:** This novel paradigm using ICT identifies likely false positives or raises suspicion that a result is truly positive, rapidly needing prompt follow up and treatment. Thus, ICT enables well-accepted gestational screening programs that facilitate rapid treatment saving lives, sight, cognition and motor function. This reduces anxiety, delays, work, and cost at point-of-care and clinical laboratories.

**Author’s Summary:** Toxoplasmosis is a major health burden for developed and developing countries, causing damage to eyes and brain, loss of life and substantial societal costs. Prompt diagnosis in gestational screening programs enables treatment, thereby relieving suffering, and leading to > 14-fold cost savings for care. Herein, we demonstrate that using an ICT that meets WHO ASSURED-criteria identifying persons with/without antibody to *Toxoplasma gondii* in sera and whole blood with high sensitivity and specificity, is feasible to use in USA clinical practice. We find this new approach can help to obviate the problem of detection of false positive anti-*T.gondii* IgM results for those without IgG antibodies to *T.gondii* when this occurs in present, standard of care, predicate USA FDA cleared available assays. Thus, this accurate test facilitates gestational screening programs and a global initiative to diagnose and thereby prevent and treat *T.gondii* infection. This minimizes likelihood of false positives (IgG and/or IgM) while maintaining maximum sensitivity. When isolated IgM antibodies are detected, it is necessary to confirm and when indicated continue follow up testing in ∼2 weeks to establish seroconversion. Presence of a positive ICT makes it likely that IgM is truly positive and a negative ICT makes it likely that IgM will be a false positive without infection. These results create a new, enthusiastically-accepted, precise paradigm for rapid diagnosis and validation of results with a second-line test. This helps eliminate alarm and anxiety about false-positive results, while expediting needed treatment for true positive results and providing back up distinguishing false positive tests.

## INTRODUCTION

*Toxoplasma gondii*, infects approximately half of all persons with 16 million people infected congenitally. Congenital toxoplasmosis (CT) causes loss of life, sight, cognitive and motor function [1–5]. In 2013 the World Health Organization (WHO) estimated there are up to 190,100 new cases of CT and 1.20 million disability-adjusted life years each year globally [4–7]. Disease burden is particularly high in Latin America and certain populations in the US and elsewhere with high exposure. Almost all untreated congenitally infected persons develop manifestations [1–17]. In addition to considerable progress toward definitive cure and prevention of *Toxoplasma* infection with novel potential medicines and vaccines [6, 7], a critical part of eliminating the disease and reducing suffering and disease burden of CT requires prompt recognition of seroconversion and expeditious, early treatment of the acutely infected pregnant women with available, effective medicines [3,6–9]. Screening monthly, beginning before or near conception to one month post-partum for development of antibody to the parasite in previously seronegative women can enable treatment to prevent trans-placental transmission of newly acquired maternal *Toxoplasma* infection or treat the fetus to prevent sequelae [3,7–17]. France, Austria, Slovenia, Colombia, Panama, Brazil, Argentina and Morocco have or are developing screening programs [8,10, 11] but the United States does/has not [3,6,7,8,12–14]. Actual and artificially inflated costs to make profit are potential barriers [1, 13–45], even though cost benefit analyses all have found substantial cost savings and benefits with routine testing [13–18]. Introduction of prenatal screening tests that fulfill the WHO ASSURED criteria (Affordable, Sensitive, Specific, User-friendly, Rapid, Robust, Equipment-free, Deliverable) can improve benefit [13–18]. False positive results using currently available commercial test kits for anti-*Toxoplasma* IgM compound problems [19–22, 38]. The United States FDA mandated that a positive result for acute infection (IgM) with a non-reference laboratory (NRL) test should be confirmed at the Palo Alto Medical Foundation /Remington Specialty *Toxoplasma* Serology Laboratory (PAMF-TSL) [22]. Associated delays cause concern for patients and their physicians. Substantial costs (more than $800 per panel of tests in the USA with additional problems for this exceptional testing from insurance denials and capitation of obstetrical health care) have been an argument against screening programs [14]. Therefore, NRL tests with high specificity and low cost are needed. A recently developed *Toxoplasma* ICT IgG-IgM test (LDBIO Diagnostic, Lyon, France, hereafter called ICT) is a promising candidate NRL test that satisfies ASSURED criteria [23–5].

To begin to implement a reasonable, feasible, low cost workflow for USA gestational screening programs, which has currently and historically been problematic (please see **Commentary** in supplemental showing those problems for care), a series of studies described herein were performed. A formal clinical trial feasibility study at the University of Chicago Medical Center (UCMC) began in July 2020. The goal was to evaluate a sufficient number of verifiable ICT results to complete the U.S. Food and Drug Administration (FDA) 510(k) clearance and Clinical Laboratory Improvement Amendments (CLIA) regulations waiver process. This study involves comparing results of the ICT to an already cleared serum test, also called predicate test (Bio-Rad Platelia Toxo Enzyme Immunoassay). When we encountered difficulties with false positive IgM results with the predicate comparator, but not the ICT, we were faced with the unanticipated constraint of cost of positive confirmation of a number of tests at PAMF-TSL, and recognition that this type of cost from the frequent false positive IgM results could de-rail screening programs in the United States. Given the true negative with ICT in our setting, we queried whether the ICT could be part of a paradigm to rule-out false positive IgM results with NRL test, both at the point-of-care and in the hospital clinical laboratory. Further, we tested samples with the ICT that were suspected false positives from local laboratories that had been referred to reference laboratories. We placed these data in the context of practical clinical problems we encountered and collated our results with ongoing and reported similar studies to define whether this could be a paradigm helpful in addressing false positive predicate NRL test results. Solving this problem emphasized how the ICT can be used in screening programs to benefit pregnant women and their families, creating a new paradigm to approach the problem of need for accurate screening and of false positive tests. This highly accurate test may help enable screening for acquisition of *T. gondii* in gestation [23–39] and thereby contribute to saving sight, cognition, motor function and lives and improve quality of life [1-10, 12-15, 17, 24, 35].

## METHODS

### Hypothesis

Our hypothesis was that a lateral immunochromatography test (ICT) we previously found sensitive and specific could meet criteria specified by USA FDA and CLIA to document that this test is useful for serologic and whole blood point of care testing to detect *Toxoplasma* infection. We performed a series of studies (**Figure 1**) to test this hypothesis in the USA, France, and Colombia. In so doing, we discovered paradigm shifting approaches and utility, proving and extending beyond our original hypothesis, in studies using methods that follow and in the supplement in more detail. **Figure 1** lists and provides a “roadmap” to 12 studies/analyses in this decades-long work. A succinct overview of methods follows:

**Figure 1.**
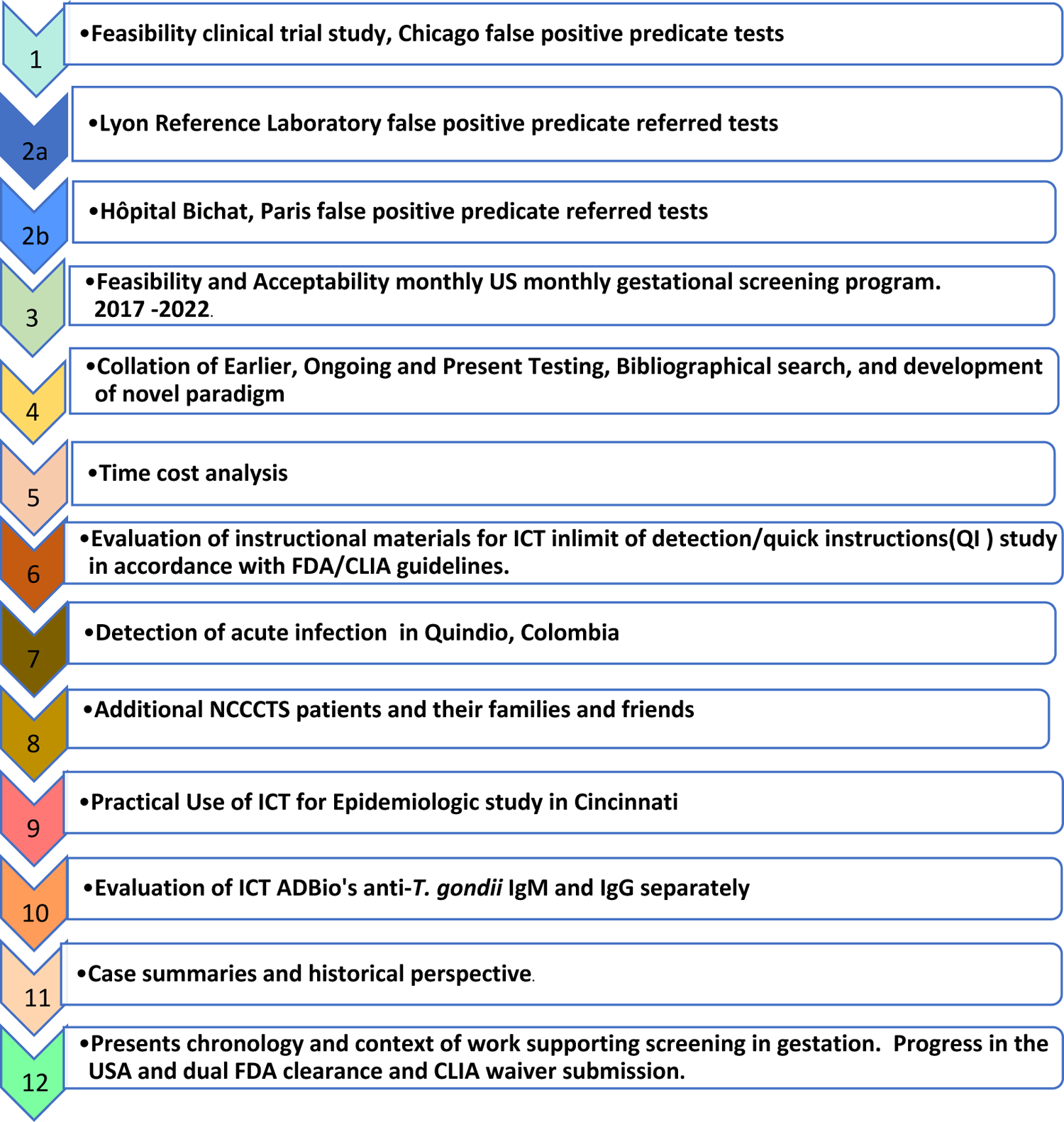
“Roadmap” to Studies Performed herein. Key shows corresponding Supplemental figures and tables.

### Study 1: Feasibility clinical trial study

The design of this study (Clinical_Trials.gov number NCT04474132) and how it is related to earlier work is shown in **Supplemental Figure S1A-E.** Serologic samples for the UCMC feasibility study (ongoing as of April 2023) were obtained from 41 pregnant women, 40 undergoing regular prenatal appointments at the UCMC (23 in first trimester, 12 in second trimester, four in third-trimester) and from seventeen non-pregnant volunteers. Each subject’s whole blood and sera were tested with ICT; subjects’ sera were also tested at the UCMC’s CLIA-approved Clinical Laboratory, which uses a Bio-Rad Platelia Toxo Enzyme Immunoassay as its FDA-cleared standard predicate test to detect IgG and IgM *Toxoplasma* antibodies. There were three testers in 3 sites. All discrepant results between ICT and predicate were sent to Remington Specialty Laboratory-PAMF-TSF or Lyon Reference laboratory for confirmation immediately using a panel of tests described elsewhere [27, 37].

### Study 2a: Additional samples from Lyon Reference Laboratory that had been referred when erroneously reported/referred by local laboratories with positive IgM

A set of 32 samples obtained at the Parasitology Laboratory of the University Hospital of Lyon, France (Institut des agents infectieux, Hôpital de la Croix-Rousse, Lyon, France) were selected for being reported as false positive with at least one first-line, NRL automated assay and confirmed to be negative by a panel of additional tests in the laboratory. They were additionally tested at LDbio Diagnostics using ICT and WB ToxoII IgG and IgM [40–45].

### Study 2b. Testing of other samples at Hôpital Bichat, Paris

A total of 558 US serum samples that would otherwise be discarded were tested at Hôpital Bichat.,Paris (Bichat-Claude Bernard Hôpital, Laboratory of Parasitologie, Paris, France [30]). Another set of samples also was tested at Hôpital Bichat in Paris. Results are being presented in detail in a separate report describing a variety of tests from Hôpital Bichat (Abraham et al, 30, and in submission 2023).

### Study 3: Testing of US Samples in a study to Determine Feasibility and Acceptability of fingerstick in monthly US gestational screening program 2017-2022

This separate study was to determine whether this ICT testing could be performed monthly for pregnant women in an academic obstetrical setting in the USA and whether it would be acceptable for patients and their physicians. This study took place in the outpatient Obstetrics and Gynecology Practice at an urban academic medical center between September, 2017 and September 2018. Patients were identified at their first outpatient obstetrical visit, between 8-12 weeks gestation, by their primary obstetrical care provider. Patients not infrequently attended their obstetrical visit with their partners. They were provided an educationl pamphlet [33] and were able to ask any questions. Patients then were offered an opportunity to participate in the monthly screening study and if they wished to do so to sign an informed consent. If the patient indicated interest in participating in the study, voluntary consent was obtained by the research team. All patients who were asked expressed interest and willingness to participate. The original intent of the study was to follow 20 women to term with monthly testing through the sixth week post-partum obstetrical visit. Each month, at the patients’ regularly scheduled appointment or shortly thereafter the patient was tested with the whole blood-variant *Toxoplasma* ICT IgG-IgM POC test. Methods for testing have been discussed in our previous work [23] and above. Serum was tested with another high-functioning test, i.e.,with the ARCHITECT-, and /or VIDAS (VITEK® ImmunoDiagnostic Assay System) as an automated enzyme-linked fluorescent immunoassay (ELFA) and/or Western Blot-Toxo-IgG and IgM systems (LDBio diagnostics) performed in Lyon, France and/or Quindio Colombia Reference Laboratories [10, 25]. We also tested an additional 25 participants in the National Collaborative Congenital Toxoplasmosis Study (NCCCTS) and our other studies during this time frame who wanted to participate.

Providers joined the study as collaborators following an Obstetrics Department Grand Rounds and Obstetrics Sectional Educational informational meeting for those who missed the Grand Rounds. Both informational meetings were presented by RMc. Providers were provided the same educational pamphlet that their patients also received. All had the opportunity to ask questions of RMc. As described above under “Patient Recruitment”, providers then mentioned the study to their patients. At the initial and subsequent visits the medical student (JL), Maternal Fetal Medicine Nurse (KL), or PI (RMc) obtained the samples after coordinating with the patient and practitioner at the time of a subsequent monthly obstetrical visit. Providers were told the results they could discuss with their patients.

Surveys, designed to assess patient satisfaction with the gestational screening program were created to use at the end of the study. Responses were based on a 5-point Likert scale, ranging from “strongly disagree” to “strongly agree.” There was also an opportunity for free response regarding strengths and potential areas of improvement for the screening program. The detail of questions is in a figure in the results section. Surveys were provided by the research nurse or others working in the study to the study participants at the 6-week postpartum visit or shortly before this visit. Contact with provision of the brief questionnaire was missed for five study participants at the 6-week postpartum visit. All five were asked and two of those five completed the questionnaire at a later time. Surveys were anonymized, so correlation of survey data to individual respondents was not possible. As part of this intent-to-study, as above, we had enrolled 22 pregnant women, and 19 continued monthly. In the Lyon, France reference laboratory, the 155 samples were tested with Abbott ELISA IgG/IgM. When Abbott Architect (France) IgG/IgM had either an IgG or IgM that was positive, backup testing was performed with VIDAS in Lyon laboratory, and LDBio Western Blot IgG/IgM (IgM performed for three tests at LDBio). In the Quindío, Colombia reference laboratory that uses VIDAS family (VITEK® ImmunoDiagnostic Assay System) as an automated enzyme-linked fluorescent immunoassay (ELFA) test, the last 88 of the 155 samples were tested in parallel.

### Study 4. Collation of Earlier Testing, Bibliographical search, and development of novel paradigm

We collated results of all of our earlier work, both published already [23-27, 29-31, 35] and other separate studies ongoing at present on related topics that are being separately submitted for publication currently [Abraham et al, in submission 2023; 30], and our current work herein. Now that the ICT is CE marked and available in a Europe, to determine whether we had overlooked any other study we were not otherwise aware of, we performed a bibliographical search on Pubmed using terms for evaluations of the *Toxoplasma* ICT IgG-IgM test. This was to make certain that we had included all reported tests in our analysis. Only English literature was reviewed. For evaluations found, full text was retrieved and searched for potential false positive samples. Additionally, results of evaluations presented in congress that were known by the authors including those in submission to other journals were added to the totals in this analysis. Earlier and ongoing studies and reports were arranged chronologically and the total collated data are reported herein.

#### Summary diagram of Novel paradigm the work presented herein develops

Difficulties we encountered initially in our clinical trial inspired organizing the algorithm we created and show graphically in **Figures S2.I** and **3** to prevent problems like those we had to address.

#### Study/Analysis 5: Time cost analysis

We found a number of approaches including reference laboratory tests with varying costs, ease of use, and considered whether they meet WHO ASSURED criteria. Advantages and disadvantages of those approaches are summarized in tabular format including a time cost analysis.

### Study 6: Evaluation of instructional materials for ICT with whole blood at point of care in limit of detection/ quality of instructions (QI) study in accordance with FDA/CLIA guidelines

The following study was performed to determine the precision of the ICT with samples at the limit of detection and whether never experienced testers could read, understand, perform and interpret instructional material for use of the ICT with whole blood. Samples were prepared in accordance with FDA/CLIA requirements and guidance for instructional material for CLIA waiver for a point of care test (Recommendations for Clinical Laboratory Improvement Amendments of 1988 [CLIA] Waiver Applications for Manufacturers of *In Vitro* Diagnostic Devices ([version of January 30, 2008 – in force and updated in 2020]). The following limits of detection, “Quick Instructions” (“QI”) study was then performed as follows: Nine testers were identified. Testers were three practicing physicians, three nurse/nurse practitioners, two medical students and one medical resident. They were not experienced with this type of ICT using whole blood or this cassette. None worked as a certified laboratory technician. They were selected to reflect categories of potential users of this test who were unfamiliar with and unskilled with this test. Each tester took the University of Chicago blood-borne pathogens and universal precautions training to work with whole blood, and their competence in understanding and using this material currently was formally documented for the study, as was IRB-required. The ability of three groups of testers with different clinical roles to read the instructions, to perform the test in accordance with the instruction, and to distinguish negative and positive results at the pre-established limits of detection were evaluated. Additional methodologic details are in the Supplement.

### Study 7. Detection of acute infection and seronegatvity in Quindio, Colombia by using ICT

Sera from 22 patients who had recently acquired their *T.gondii* infection in Quindio Colombia and 12 patients who were seronegative had sera that were tested with Vidas ELIFA IgG, IgM and with ICT.

### Study 8. Additional NCCCTS patients and their families had testing with ICT while at follow up visits in Chicago to add data determine antibody present for many years is still detectable by ICT

Between March and December 2018 20 participants in the NCCCTS whose serologic status was known from earlier reference laboratory testing and family members traveled to Chicago. Number of tests were 20 seropositive and 8 family members or other controls for separate studies such as multimodal neuroimaging studies were found to be seronegative but did not have other reference laboratory testing. Time from acquisition of infection was noted, in a similar approach to earlier studies of Begeman and Lykins.

### Study 9. Use of ICT for Epidemiologic study in Cincinnati

Sera and demographic data from a maternal-infant cohort in Cincinnati were available for 265 women; 264 had data on the variables of interest. Variables of interest included residential address (longitude and latitude), age, education, race, income and pet ownership as part of the original cohort study. Sera were tested with ICT and if positive then were tested with IgM and IgG western blots at LDBio. A logistic regression model on the results for the 264 samples was used to estimate the ICT *Toxoplasma* infection positive status by including independent variables such as, maternal age, marital status, Neighborhood Deprivation Score (a higher value means more deprived and missing values are extrapolated from 5 nearest neighbors), latitude, longitude, race, i.e., White or not, and an interaction term between maternal age and Marital status.

### Study 10. Evaluation of lateral chromatography test AdBio that detects anti-*T. gondii* IgM and IgG separately

To determine whether a USA made immunochromatography test (called ADBio) would function as well as the ICT or whether samples from Colombia that had very high performance with ICT would function as well with a different USA made test, an additional set of known positive IgM positive or IgM negative samples was tested. The tests that were used were the VIDAS (Quindío, Colombia) test and another commercially available but not FDA cleared or CLIA waived test that detects *Toxoplasma* specific IgM in the Colombian Reference laboratory. A total of 147 serum samples were included, selected from the biobank of past studies at the University of Quindío in Armenia, Quindío, Colombia. All samples were previously tested using the reference test VIDAS (VIDAS Toxo-IgG Avidity kit; bioMérieux, Marcy-l’Etoile, France). Samples were divided into the following three groups as defined by VIDAS testing: (1) IgG negative and IgM negative (n=65), (2) IgG positive and IgM negative (n=55), and (3) IgG positive and IgM positive (n=27).

### Analysis 11: Representative Case Summaries illustrative of practical clinical problems where solutions are needed and potential utility of ICT

Representative case vignettes with concepts they illustrate were collected and were summarized to illustrate impact and need of this paradigm and its historical context (as illustrate in “**Studies 2a,b, and 10”**). These brief case summaries are from The National Collaborative Chicago-Based Congenital Toxoplasmosis Study and Consultations to the Toxoplasmosis Center and Toxoplasmosis Research Institute. They illustrate representative, frequent clinical problems incurred from false positive IgM tests. Representative examples of benefit and novel utility of ICT in addressing this problem are also presented. A case summary also presents use of ICT for early detection confirming pre-conception infection when sequential samples obtained in the context of *in vitro* fertilization (IVF) were available. Commentary about screening programs and their absence in the USA further place our findings in an historical perspective in the Supplemental.

#### Ethics

The ongoing UCMC study, under the name “Prevention of CT: Feasibility of prenatal screening using point-of-care tests,” is conducted with ethical standards for human experimentation established in the Declaration of Helsinki. Research received approval from the University of Chicago Institutional Review Board (University of Chicago IRB Protocols 20-0442, 19-0505, 21-1446, 8793, 8794, 8795, 8796, 8797, 8798, 16708A and met the standards of the Health Insurance Portability and Accountability Act. Results were or will be reviewed by/discussed with the Data Safety Monitoring Board. Informed consents were obtained from subjects in accordance with the University of Chicago Institutional Review Board and the National Institutes of Health guidelines. No subjects are under age 18 years. All participants provided informed, written consent prior to their study participation. All studies were performed in accordance with the Declaration of Helsinki.

Samples from the Lyon Reference laboratory of the University hospital were anonymized in this analysis. Testing in Colombia was performed in accordance with local Ethics Committees approvals and guidelines. Stored sera from Cincinnati was approved by the IRB for future testing for a wide range of pathogens.

## RESULTS

A more detailed version of the Results including Tabulated primary data and approaches in Figures are in the Supplement.

### Feasibility clinical trial study performed exactly as the test would be used in practice, 2020 to 2021 demonstrates feasibility, identifies false positive predicate test results and develops new paradigm to help to obviate that problem, Study 1

Individual results in the ICT and the predicate test in this ongoing clinical trial study are in **Figure 2A** and **Supplemental Table S1**. Between August 2020 and December, 2021 we performed a prospective clinical trial in which 43 seronegative and 15 seropositive persons were tested with the ICT using whole blood and sera. The 58 sera were also tested with the predicate test used by The University of Chicago Medicine Clinical Serology laboratory, the Biorad assay. Results of all readers of the ICT were uniformly concordant. Any positive results were tested in Reference Laboratories.

**Figure 2.**
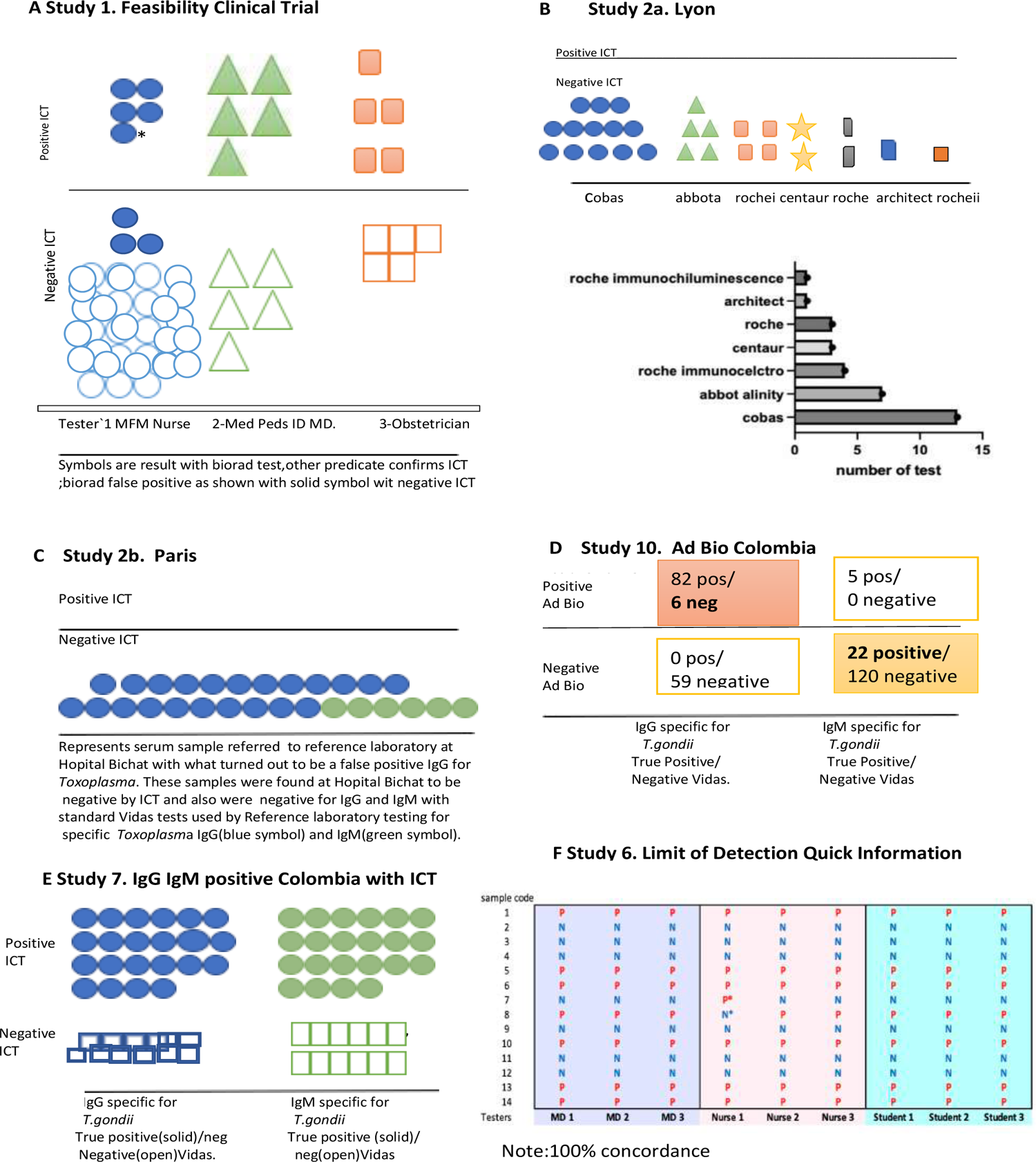
High performance of ICT. A. Clinical Feasibility Trial. Solid symbols represent positive predicate test, open circles represent negative predicate test.* False positive IgM predicate test for seropositive person. **B. ICT is negative with 32 false positive standard tests in Lyon. C. ICT is negative with false positive standard tests in Paris,** Blue IgG, green IgM. **D. ADBio, a USA test, suffers from false negative and positive IgM results. E. ICT detects acute infections with positive IgG and IgM in Colombia. F. Quick instructions** are able to teach 9 testers, 3 physicians, 3 medical students and 3 nurses to use test accurately with 7 samples that are negative and 7 samples at the limits of detection. Detailed methods, results and figures are in the Supplement for **Figure 2 A. Figure S1A-E, Table S1; B. Table S2; C.Table 3; D. Table S8; E.Table S7; F. Figure S4.**

Within testing of the initial 6 pregnant participants, we encountered false positive results for two participants in the IgM predicate test and three others later revealing a false positivity rate of 10%. Finding these frequent false positive predicate test results was disturbing for patient participants, providers, and investigators. Correcting the erroneous predicate test data with follow up gold-standard testing was time consuming, costly, and results in delays in care. Although the resource Reference laboratories in the USA and France provide one excellent solution to the problem of false positive results, that is of high quality results, the delays, cost and inconvenience were substantial difficulties. This occurred while we as investigators could promptly see the negative ICT result in whole blood and sera testing. We knew the results in the context of our earlier data with very high performance, sensitivity, and specificity of the ICT. Our earlier work had demonstrated negative results were accurate with the ICT for erroneous predicate test results considered as positive when samples were referred to the USA reference laboratory. This occurred when 33 patients with 60 sera were tested (**Tables 1 to 3**). We recognized that if we could not solve this problem of false positive predicate tests, this would have de-railed the research study and its longer-term goal of proper testing in systematic gestational screening programs for the USA and as a model for other countries. For the USA, along with their present high costs in the USA, false positive tests also would substantially harmfully influence the use of screening programs (**Figure 3, 4, S2.1, Tables 4, 5, S6**).

**Figure 3.**
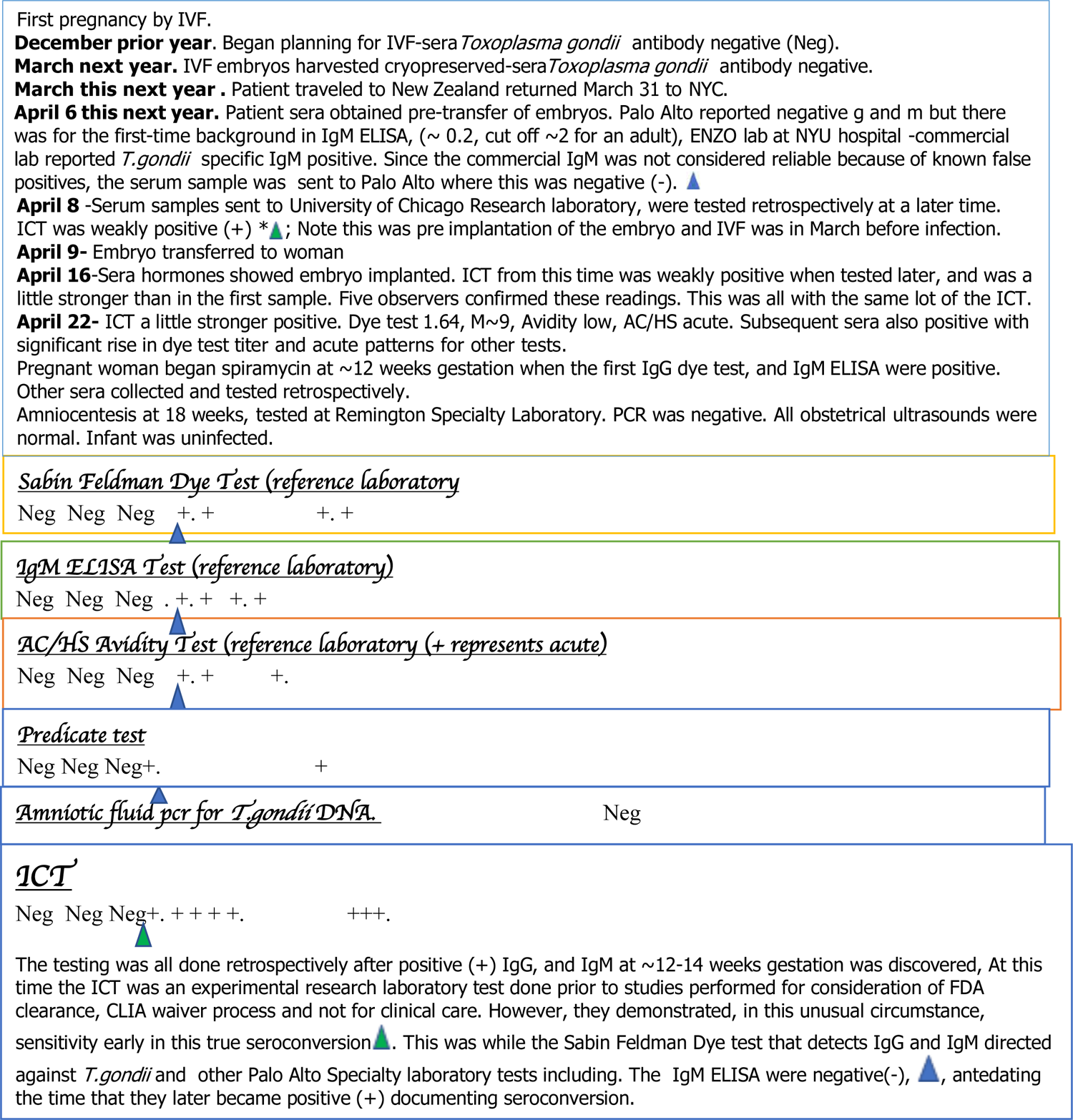
Representative example of ICT detecting very early seroconversion using sera originally stored for another purpose tested retrospectively, showing contrast with false positives. Blue triangle marks tests at time of seroconversion detected by ICT marked with green arrow. + represents positive result. There are a few examples of ICT positive with IgM only early and others G and M only early in seroconversion.

**Table 1.**
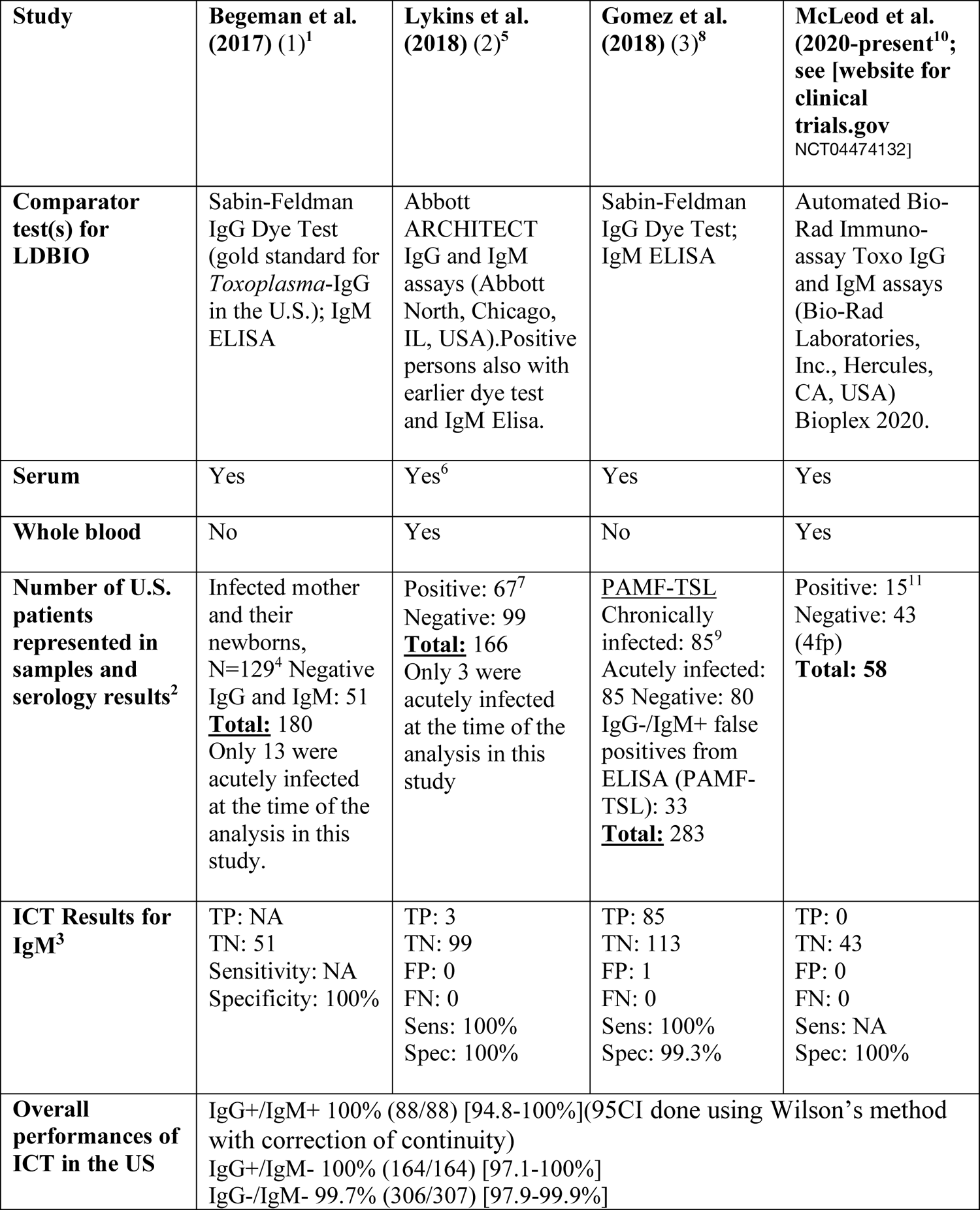
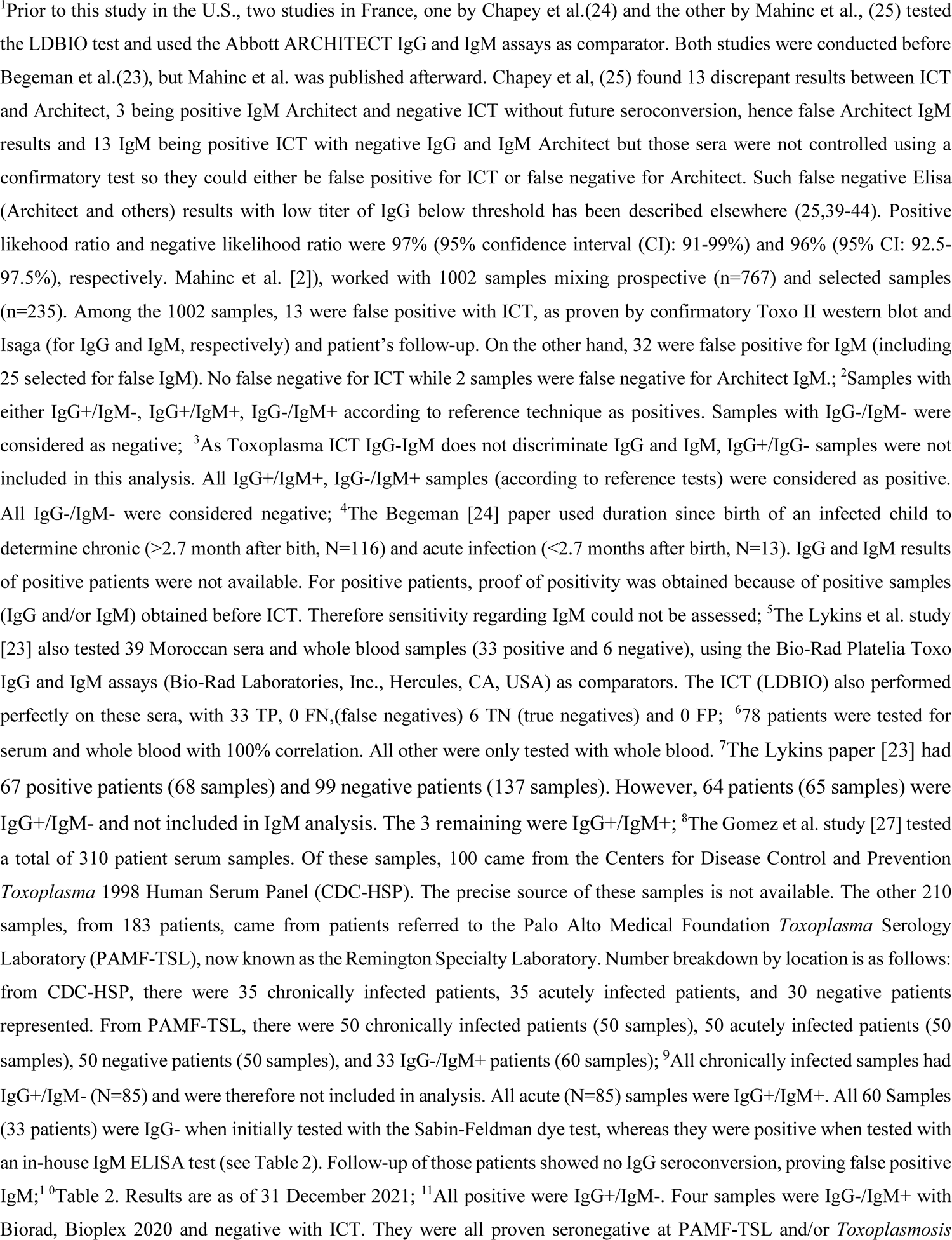
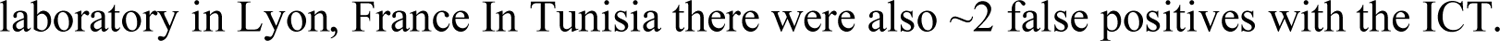
Summary of Studies that have employed the LDBIO *Toxoplasma* ICT IgG/IgM test (ICT) on U.S. sera or whole blood samples, with performance results

### New paradigm elucidated by this experience

Thus, as we tested the whole blood and sera with the ICT in parallel with testing the sera with the Biorad test, we developed an easy, inexpensive paradigm shifting approach to solve this problem of false positive tests for the USA. This paradigm (**Tables 4, 5, Figures S 2.I, S 3**) showed the ICT could help to eliminate the problem of false positives both in the clinic and the clinical laboratory. This paradigm was to have a method for diagnosis with a test that meets WHO ASSURED criteria available promptly at the time the test was performed and to have a first backup of positive results in serum rapidly in the local laboratory. Our experience shown demonstrates that this ICT performs properly in clinical practice and field studies. We noted that it could be used correctly by previously untrained observers, meeting WHO ASSURED criteria. We found that this also could help to obviate the difficulties caused when a commercial predicate test has false positive result. This was while introducing a novel test that could be low cost and easy to use in the clinic. It became clear from the experience and data presented that this novel test and paradigm could be a useful new method for the clinical laboratory to identify true positives rapidly for this emergent problem/disease. This could help to determine whether there was need for further screening. It could help to clarify whether there was need for emergent care with life, sight, cognition saving medicine should be initiated promptly while waiting for backup reference laboratory confirmation of a true positive test.

### Additional testing of erroneous false positive local predicate tests with ICT and gold-standard testing in reference laboratories demonstrates utility of the novel paradigm with ICT, Study 2

Then, Lyon and Paris Reference laboratories’ identified erroneous false positive results reported for samples referred for testing from local private laboratories that used commercial tests. Thirty-two samples that were referred to the Lyon reference laboratory from September 2021 to February 2022 from private laboratories because of the detection of isolated IgM in the course of monthly prenatal retesting, which the main system used in France in this context (**Figure 2B, Table S2**): The tests that had been used included Cobas Roche (n = 21), Abbott (Architect n = 1 or Alinity n=7), Siemens (n = 3) (**Figure 2B, Table S2**). None of the additional 32 samples gave positive results with ICT or in the reference laboratory with Abbott Architect despite the erroneous reports of positive IgM results (**Table S2**). Further, in Lyon France, none of the 32 false positive IgM tests with the predicate local laboratory tests used had false positive test results with the ICT or gold standard Western Blot (**Table S2**). This was using the same referred serum that was tested and reported to be positive from the local private laboratory. These results are included in **Tables 3 and S2, and Figure 2**.

### Placing US ICT in the context of other ongoing studies, including Study 3 and previously published studies, demonstrates high performance, Analysis 4

Data summarized in **Tables 1 to 3** place the results in the Clinical Trial and monthly screening acceptance studies (**Study 3**), in the context of other ongoing studies and our earlier published work. **Table 3** addresses details of studies in the USA and elsewhere. **Tables 1 to 3** also collate and address studies of false positive IgMs referred to reference laboratories in the USA and France. **Table 3** collates all these studies including those of other countries as a summary of all available results. Sensitivity, specificity, confidence intervals and details of studies are in **Tables 1 to 3**. Performance of ICT is high, sensitivity≥ 98.5%, specificity ≥98.9% (serum and/or blood).

Additional testing of a set of samples with Architect, Vidas and other tests was performed in an additional matrix analysis pertinent to consideration of false positive test results. Back up testing of another set of sera from a monthly screening and acceptability of the monthly screening program was performed in the Lyon France and Quindio Armenia Colombia Reference laboratories (**Table S6**). In these reference laboratory settings as well as in the Paris Hôpital Bichat reference laboratory [30] false positive test results were less problematic (**Figure 2 C, Table S6**) than in the clinical trial. There were no false positives in the Quindio laboratory Vidas testing and one patient with multiple consecutive IgG false positives in the Lyon Architect tests.

### Bibliographical search confirms high performance and that data analysis herein includes all published studies

As the ICT is now commercially available following CE Mark approval in Europe, we also used a bibliographic search to attempt to identify results with which we might have been unfamiliar or with inferior performance of the ICT. **Table 3** details all studies performed to date: Number of persons (N), Sensitivity (Se)/ Specificity (Sp), country of samples, N of false positive IgM, ICT results on false positive IgM and confidence intervals, testing for serum and whole blood are in this Table. As the ICT is now commercially available following CE mark approval in Europe we also used a bibliographic search. To date all studies have involved the authors of this manuscript. There were no additional studies identified that have been reported to date. A total of 4606 sera, 1401 positive and 3205 negative, and 1876 whole blood tests, 728 positive and 1148 negative tests have been performed, including all published, ongoing studies and those herein with high sensitivity and specificity (**Table 3**).

### Overall performance of ICT with NRL tests false positive IgM also is high, Study 2 a and b

**Overall,** including our own results herein, we found 137 samples with false positive IgM in at least one NRL technique also tested with ICT, among which 132 were found negative in ICT. The specificity of ICT for false positive IgM was 96.4%. Three samples were from IgG negative pregnant persons in Chicago. Two seropositive persons also had false positive NRL IgM was not found in Reference laboratory IgM tests. In addition 22 false positive IgG results were correctly identified at Bichat-Claude Bernard Hôpital, Laboratory of Parasitologie, Paris, France [30](**Figure 2C, Table 3**) and 5 false positive IgG identified in the predicate Abbot Architect in the University of Chicagomedicine samples tested in Institut des agents infectieux, Hôpital de la Croix-Rousse, Lyon, France (**Table 3**).

### Time, cost, comparing tests demonstrates time/cost savings and aids in eliminating delays, Study 5

An analysis of relative time and cost is in **Tables 5**. The ICT is substantially time and cost saving,

### Representative case summaries illustrate problems in care that false positive tests can cause and utility in identifying seroconversion in infection acquired prior to conception, called study 4b and “analysis 11” in methods)

**Table S8** discussed in **Supplement Commentary** has brief summaries of some consequences of false positive and negative results in ongoing cases in USA clinical practices. These provide further evidence of problems that false positive test results cause.

**Figure 3 (Study 4b)** shows utility in identifying infection prior to conception. The ICT detected seroconversion a day earlier than the Sabin Feldman Dye test and IgM ELISA in the Reference laboratory. There are a number of examples of patients who developed M alone then M and G [26]. In the Mahinc et al study [26, **Table 3**] there were 50 serum samples from 24 women for whom there were 17 samples with IgM only and 33 samples with IgM and IgG; ICT was positive for all samples except one that had a borderline IgM ISAGA of 5 for a patient who later was found to have acute *Toxoplasma* infection. There were also another 144 acutely infected persons identified in the USA, France, Morocco and Colombia all identified as positive with the ICT [24, 25, 27, 29, 30] **Table 3**. It was unusual, however, to watch seroconversion with as much precision in narrow time intervals so early in infection as shown in **Figure 3**.

**Figure 3**, compared with Figures S7 and 8 contrast current status and consequences of CT at earlier times and continuing to present in France and in the USA. This illustrates that the ICT and gold standard back up testing can solve a substantial health care problem. This is both in a historical context and at present, with potential spillover benefit for care for pregnant women and their families.

### Testing ability of written instructional materials to facilitate clinical use of the ICT by healthcare practitioners not skilled in using the ICT in a limit of detection quick information study per FDA and CLIA instructions demonstrates high performance, Study 6

Moving toward implementation, ability of written instructional material to be used in clinical practice with samples at limits of detection for positive whole blood samples and negative whole blood samples was found to have perfect performance. This perfect performance was for all the “blinded” readers and testers performance and reading after they read the Quick Information (QI) materials (**Figure 2F, S4**).

### Feasibility and acceptibility of monthly gestational screening with ICT is demonstrated in Study 3

Early in the work with the ICT in 2017 to 2018 we performed this study to determine whether monthly gestational screening would be feasible in a research study setting (**Figure 4, Table S6**). Results of the testing did not enter standard medical records or the EPIC system at that time and the testing took place earlier than study 1 but was completed after that study. This study was initiated before the clinical trial and led to the initial meeting with the FDA when a program officer from the Thrasher Foundation emphasized the importance of FDA clearance for the test to be useful to help patients in the USA. We also asked participants at its completion whether participants felt it was important and comfortable to have knowledge about *Toxoplasma* in pregnancy and whether they would want serologic testing and/or the finger stick point of care test in subsequent pregnancies if it were approved in the USA. The intent was to determine whether screening might be acceptable in standard academic obstetrical USA practice: Some parts of the study, e.g., the questionnaire and additional backup testing were performed in 2020. Results for the initial tests for the participants’ visits were included in the earlier 2018 publication [23]. Thus, numbers included for this study in the cumulative total of tests were subtracted from tests shown in **Figure 4**.

**Figure 4.**
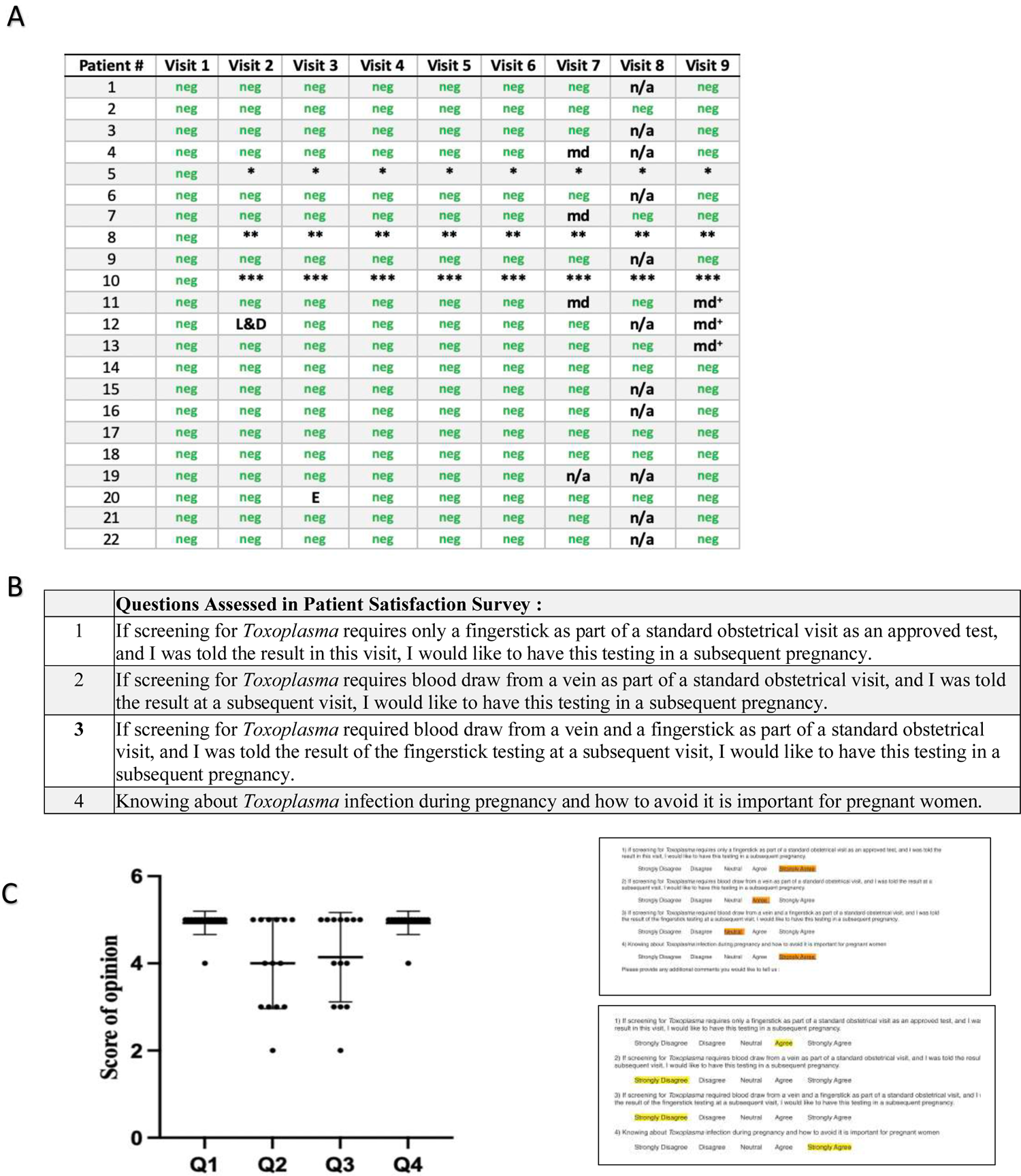
Feasibility and acceptability of monthly screening in U.S. Academic practice. **A. Times and results of monthly screening for each participant. B. Survey questions and Likert scale.** n the survey, number responses were considered as follows: strongly agree was 5, 4 was agree, 3 somewhat agree, 2 was somewhat disagree, 1 was strongly disagree **C. Results of satisfaction survey for 14 participants**. Each respondent’s answer is represented by the circles in the figure. The mean is indicated by the horizontal line, with error bars indicating standard deviation. In response to questions 1 and 4, 13/14 respondents indicated that they “strongly agree” that they would pursue testing for *T. gondii* in future pregnancies with POC testing, and that knowledge of *T. gondii* is important for pregnant women. Results were more mixed if testing required venipuncture, as indicated by the responses to questions 2 and 3, but most agreed that the test was important, would have it again in a subsequent pregnancy, and would recommend this to family and friends. It was noteworthy that other family members such as fathers of the fetus asked to be tested, relevant to the possibility of retinal disease. Right panel showed results after the time of the 6 week post partum visit. ‘We did not conduct a formal survey at the time; however, we began with two providers, and other providers in the practice asked to join with their patients. Providers continued in the further analysis of the ICT as it moved toward clearance and waiver.

Patients were all identified at between 8-12 weeks gestation. Patients had a median age of 31 years (range: 24-40 years). Seven of the 22 participants were nulliparous, while the remainder had been pregnant once or twice before. None reported having been tested for *T. gondii* infection in the past. Participants were enrolled in the study between September and November, 2017. The study initially concluded in September, 2018 with the birth of the last participant’s child. Because five mothers were missed by our study group at their 6-week postpartum visit, an anonymized questionnaire was provided in 2022 for those participants. This was considered separately in our analyses. Patients were tested at monthly intervals after their initial enrollment and tested until their 6-week postpartum visit. A small subset of patients (3/22) were withdrawn from the study: One individual underwent elective termination due to fetal anomalies. One participant suffered a spontaneous abortion. The third patient chose to withdraw from the study due to traditional beliefs about dangers associated with venipuncture. No patients (0/22) had evidence of prior infection with *T. gondii* upon their initial testing with the whole blood POC test, and none seroconverted during gestation.

One participant had a faint band suggesting the possibility of a positive test on one test, but this was only visible to the naked eye and could not be independently confirmed with photography. Per manufacturer instructions, this test was interpreted as negative. There was 100% concordance between testing interpretations of the POC test and confirmatory testing, including the ARCHITECT/Vidas/Western blot systems and the serum-based POC test variant, commercially available and now CE mark approved in France. The course of gestational screening for each participant is presented in **Figure 4, Table S6**.

Initial response of patients and their families to screening was noted. No patient declined and responses were enthusiastic from patients. There were even requests from patient participants about whether other pregnant and non-pregnant family members and friends could join. For example, even some fathers asked to be tested to know their own serologic status and if they might be at risk of retinal disease if infected. At the end of the consecutive screening tests, we administered the patient preferences survey to an available subset of the cohort (14 in total) at their 6-week postnatal visit. Those participating women who were missed completed the questionnaire in 2022. The POC testing and screening for acquisition of *T. gondii* in gestation was well received by all participants. There was not a formal questionnaire for providers. Rather, level of interest and enthusiasm was reflected by the following: All providers remained involved in the study with their patients. Additional providers in the practice noting the ongoing study with the initial providers asked to join. Those still practicing at the University of Chicago at the later time did continue to collaborate in the subsequent clinical trial study presented herein. These objective measures documented continued involvement rather than a formal survey. All providers found the finger-stick testing and monthly screening a constructive addition to their practice. The rapidity of obtaining the results was viewed positively.

### ICT detects early seroconversion and distinguishes additional seropositive and seronegative samples in USA, France and Colombia, Studies 4b, 7 and 8

We noted, as shown in **Figure 3,** the ability of the ICT to detect very early seroconversion in a study of sera obtained at narrow intervals to monitor hormone levels during *in vitro* fertilization (IVF), that happened to occur during very early seroconversion. This was a USA patient whose *in vitro* fertilization had occurred 6 months prior to implantation of their embryo at a time that she was seronegative. In the interval between *in vitro* fertilization and implantation she had traveled to New Zealand where she likely acquired *T.gondii* infection in the weeks before implantation as shown in **Figure 3**.

Our earlier studies (**Tables 1, 2, 3**) have all identified perfect performance in detecting sera from those with acute infection in the USA. It was, therefore, of interest to determine whether the same would be found in sera from patients with acute infection with the genetically different parasites found in Colombia. **Figure 2E, Table S7** shows perfect ability, sensitivity, and specificity of the ICT to also identify acute infection (IgG, IgM) in Colombia (n=22) and those who are seronegative (N=12), (p<0.0001)(sensitivity 100% and specificity 100%). This brought the total to 144, as above, Further, serologic status was correctly identified for additional NCCCTS participants tested with finger-stick whole blood and serum between March and December 2018 herein (N=20 positive chronically infected persons [times after infection years were known to be greater than 17 years for all except 3 persons, and 5 negative). As we found perfect correlation of testing of whole blood obtained by fingerstick and serum testing in the United States, herein, and almost perfect correlation in Morocco this ICT using whole blood accurately distinguishes seronegative and seropositive status as occurs in seroconversion. Indeed, in the study in which we tested whole blood (that contained serum that originally had 38 UI/ml of IgG and 63.89 ratio for IgM according to Roche *<u>Toxoplasma</u>* kits, diluted 1:89 in whole blood from a seronegative donor) at the limits of detection in the “QI study”, there was high accuracy in distinguishing positive and negative samples (**Figures 2F, S4**). There were N=63 negative and 63 positive, making a total of 126 tests of samples performed. There was 100% accuracy both with the cassette and with photographs read by the tester and two additional readers. All readings were congruent and consistent. All were blinded for 9 testers with 3 testers in each of three settings (physician office, nurse health care setting, and laboratory conference room], and with testers differing professional backgrounds (3 nursing, 3 medicine in training, 3 licensed physicians in practice previously unskilled in use of such a test) (**Figure 2F, detail in Figure S4)**,

**Table 2.**
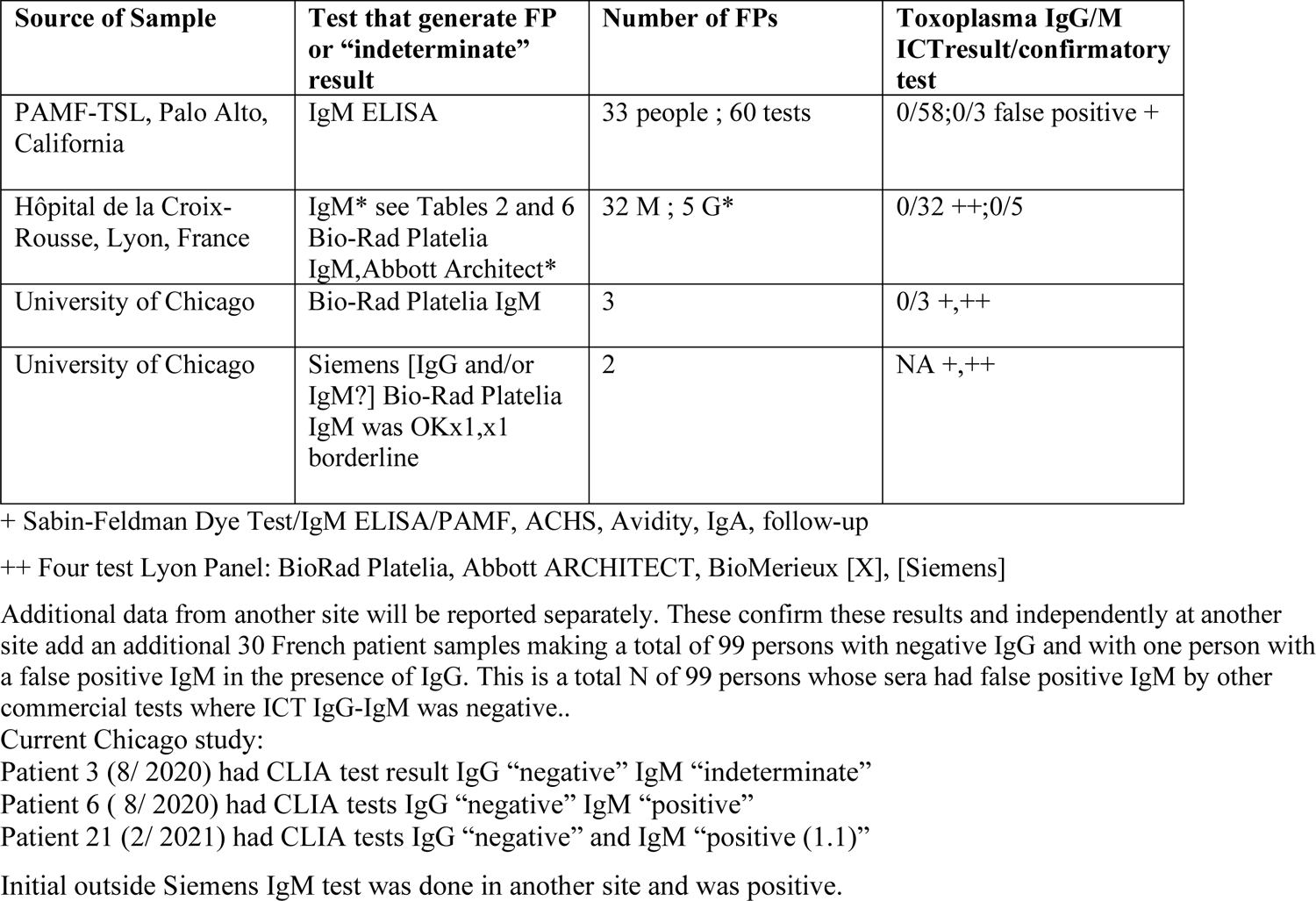
Comparison of *Toxoplasma* ICT and comparator predicate tests with false positive results

### Use of ICT for Cincinnati epidemiology study between 2017 and 2019 demonstrates that ICT is an efficient way to perform such studies and that prevalence is low in Cincinnati, Study 9

Of the 265 mothers tested, 8 (3%) had a positive IgG for *Toxoplasma* infection. None of these had a positive IgM. Variables of interest were available for 264 of the mothers including residential address (longitude and latitude), age, education, race, income and pet ownership as part of the original cohort study. There were no significant associations of testing positive for *Toxoplasma* infection and any of these variables (**Figure 5**).

**Figure 5.**
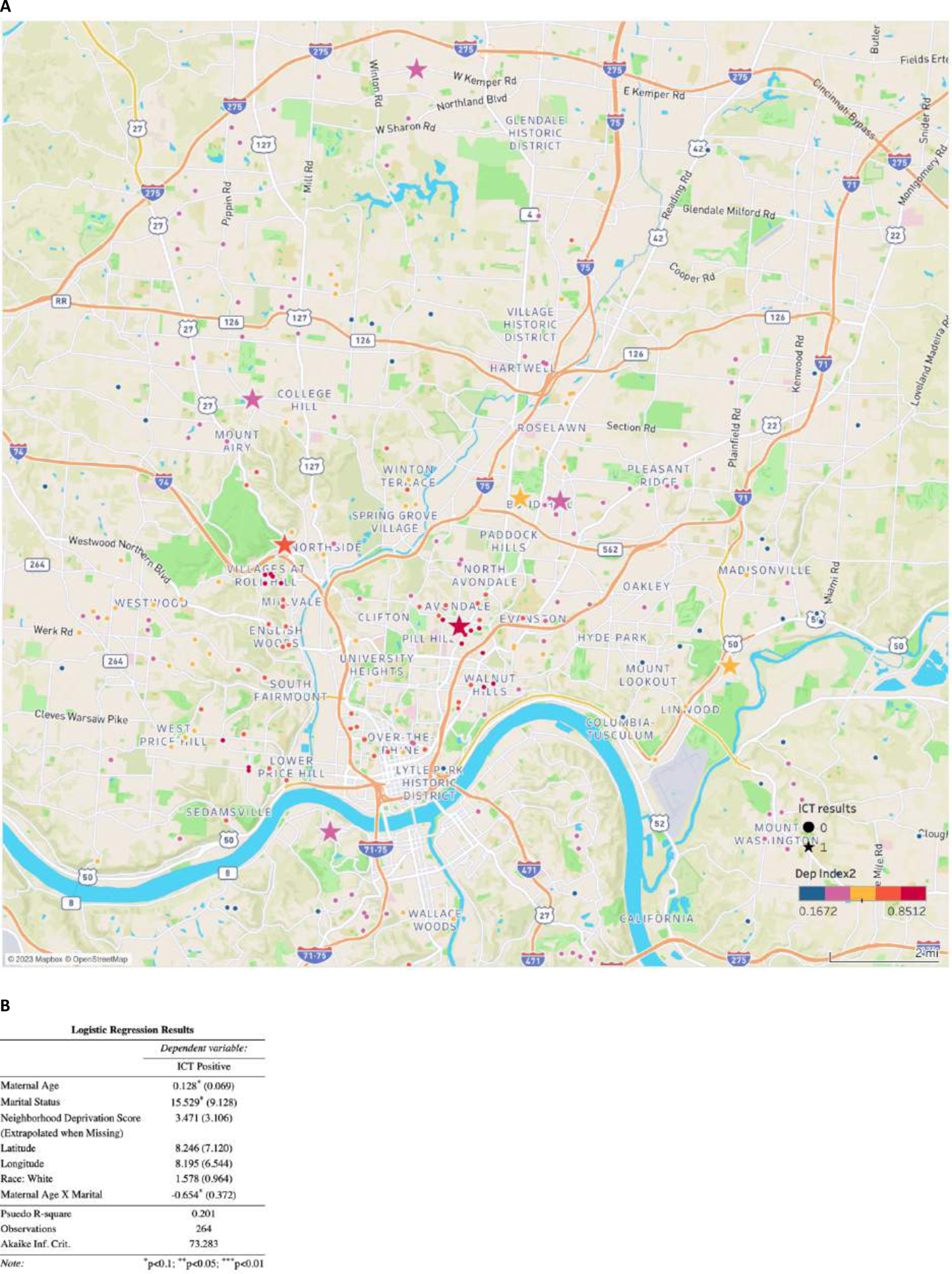
Location of seropositive persons in Cincinnati and associated demographic factors such as socioeconomic status, maximum educational level achieved, pet ownership, and ethnicity. Sera were collected between 2017 and 2019. The low prevalence of seropositivity did not allow testing for clustering by any known risk factors for infection, including proximity to watersheds associated with sewage water run-off. None of the sociodemographic parameters, neighborhood deprivation, nor residential latitude and longitude measures achieved statistical significance. The measures of neighborhood deprivation are indicated by the color of the symbols.

Results with ICT AdBio test (Onsite POC) that detects anti-*T. gondii* IgM and IgG separatelyhas both substantial false negatives and false positive IgG (9%) and false negative IgM(18.5 % true positives detected), Study 10.

We hoped that a test developed in the USA called ADBiothat is purported to distinguish IgG and IgM specific for Toxoplasma might be useful in a field setting. This test had a high proportion of False negative and substantial number of False positive results (**Figure 2 D, Table S8)**. Please also see detail in Supplemental.

## DISCUSSION (Also Expanded Introduction and Discussion are included as the Commentary in the Supplemental)

The results above demonstrate that the ICT has proven effective at identifying sera and whole blood samples of USA and non-USA patients with known *T. gondii* infection. It detects seroconversion early in infection. It is also was effective at identifying the false positive test results for *T.gondii* specific IgM of other, currently FDA cleared tests of sera when no *T. gondii* specific IgG is present. It was well-accepted in a monthly screening program that was shown to be feasible in a USA academic obstetrical practice. It also functioned with high precision while meeting WHO ASSURED criteria even in whole blood samples at the limit of detection of specific anti-*Toxoplasma* antibody. It was found to be straightforward for physicians, nurses and medical students and a medical resident to easily learn to use the ICT and accurately interpret the ICT results using the Quick Information in simple written instructions (**Figures 2F, S4**).

Up through and including the current stages of the clinical feasibility trial at the University of Chicago Medical Center, diagnostic sensitivity has exceeded 99% and specificity has stayed at 100% with all samples of U.S. patients. In addition, across several of these studies, this ICT has outperformed other screening tests. Herein, out of 99 IgM false positive sample results, across multiple consecutive different USA and French sets of data recently there have not been false positives or false negatives. In addition, in two countries (the USA and Morocco [29]), the ICT has not had false positive or borderline bands when testing the serum and/or whole blood. While it was already known that this test could perform accurately (**Tables 1-3, S1-7**), this present work also has evaluated the ability of ICT to correct the errors of other carefully tested, commercially available screening assays [27] using prospectively and retrospectively collected sera in the USA, France and Morocco. The high specificity is a particular strength for the ICT IgG-IgM device, especially when compared to other currently available commercial tests for anti-*Toxoplasma* IgM.

**Table 3.**
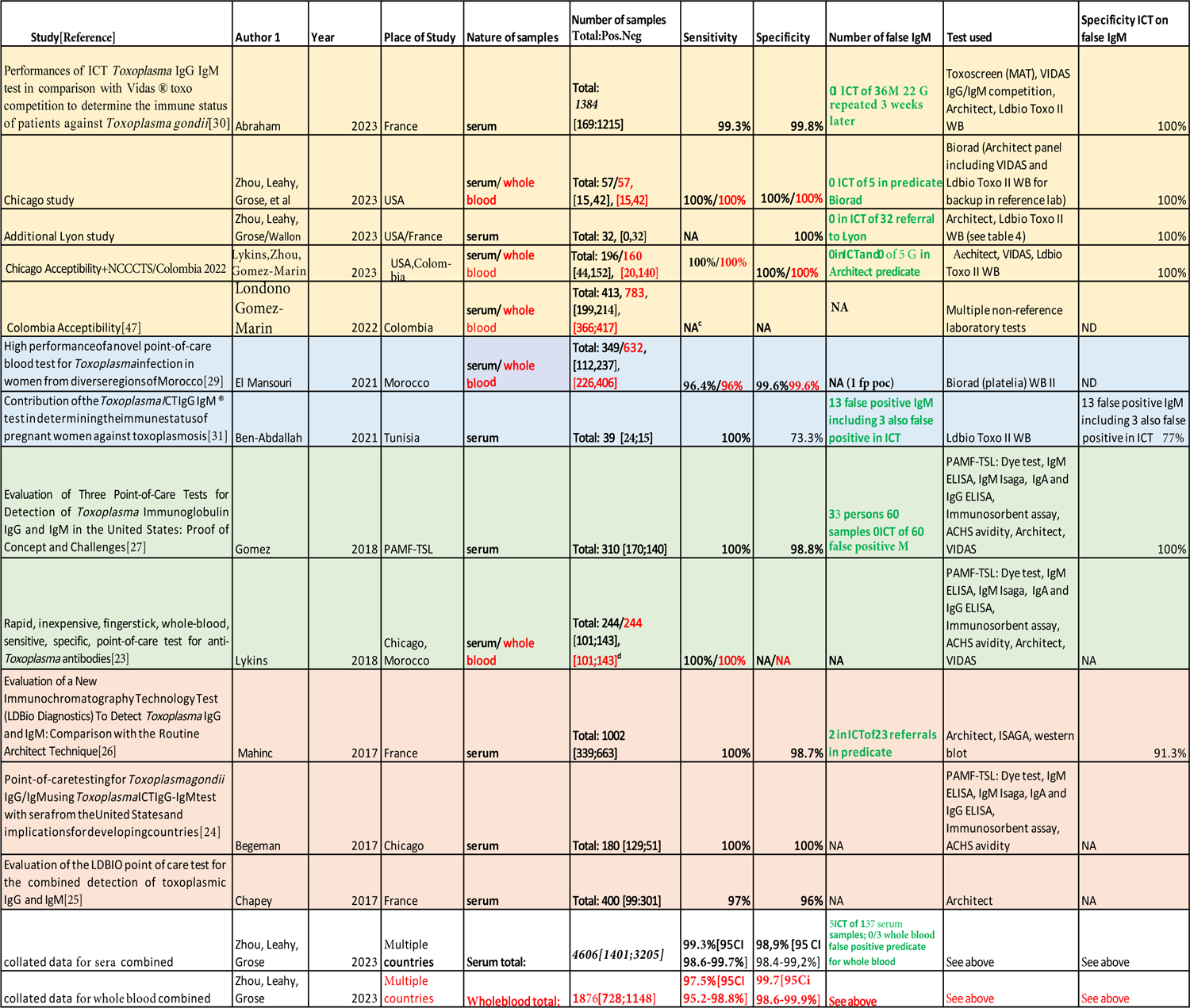
All studies with ICT

**Table 4.**
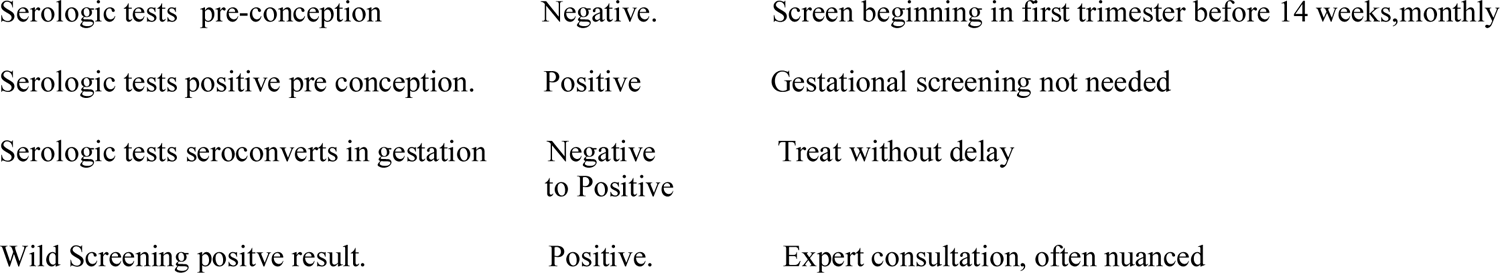
Approach to screening (also see Supplement Figure S2.I)

**Table 5.** Cost and Time saving just for fingerprick, all sample handling, reporting, billing for ICT at Point of Care verssu Predicate testing

| <u>Parameter</u> | <u>Point Of Care Test</u> | <u>Predicate</u> |
| --- | --- | --- |
| <u>Time for test.</u> | <u><2 minutes to perform 20 minutes to read.</u> | <u>Days</u> |
| <u>Location.</u> | <u>POC.</u> | <u>Requires Lab</u> |
| <u>Time to enter into the system and report.</u> | <u>Minutes.</u> | <u>Over days Multiple Levels</u> |
| <u>Cost</u> | <u><\$10 per test.</u> | <u>>\$500 per test in the USA</u> |
| <u>Participant Preference</u> | <u>Yes.</u> | <u>“Ok but POC Preferred by some”</u> |
| <u>False positive results</u> | <u>Very rare ; high sensitivity and specificity</u> | <u>Not infrequent ; problematic</u> |
| <u>Solves problem of false positives</u> | <u>Can be very helpful; also as back up test in lab</u> | <u>Can cause problems in clinical lab</u> |

The data from Houze et al (ECMID and manuscript in preparation [30] and Mahinc et al [26] increases the number up to 137 of such false positives IgM studied with the ICT. Mahinc et al also studied 23 false positive Architect and/or Biorad Platelia IgM [26]. In the Mahinc study [26], false positive IgM in the Biorad test were obviated by ICT testing 21/23 of the time. In Tunisia [31], recent results were similar adding additional data but with a higher proportion of false positives [31]. Ten of 13 false positives were negative in the ICT. Although there were no ICT false positives in these data sets in the US, the occasional false positives (5 of 36) in the work earlier in Marseilles and Tunisia emphasize the importance of confirmatory testing of positive results. The high quality performance of some of the Reference tests emphasize that some tests seem to perform better than others when used in Reference laboratories (**Figure 2E, Table S6, S7**).

Our studies, along with the earlier experience in the Palo Alto reference laboratory and collated recent results, demonstrate practical problems in the US with potential serious consequences for patient care [35] where the ICT can be helpful in a patient’s management. This has been confirmed in France making a total of 132 of 137 for IgM) and 27 of 27 times for IgG times that a false positive result could be corrected. False negatives are uncommon but would be detected by repeat testing in gestational screening programs. Any positive ICT during gestation would have confirmatory testing to differentiate IgG and IgM. The occasional false positives would be detected by back up testing in the reference laboratory in the USA or use of multiple tests including the Western blot in France. Reference laboratory gold-standard testing and certain commercially available test reagents have higher performance than testing in local laboratories as shown in **Figures 2B, 2C, 2E, Table 3, S1, 6**. The ICT has high precision with samples at the limit of detection. That the test is easy for medical students, a medical resident, practicing board certified physicians, nurse/nurse practitioners without familiarity with the ICT to perform and interpret (**Figure 2F, S4**) is congruent with a recent experience with 30 practitioners in Armenia, Colombia [47]. This experience was with patients infected with genetically distinctive Colombian parasites [47]. Acceptability in a Colombian patient and obstetrical practitioner group was high [47], similar to acceptability in our USA experience presented herein.

Colombian sera also were tested in Colombia [47–50] with a different lateral chromatography test made in the USA called the ADBio. This test differentiates IgG and IgM and has a USA sale price more than ten times that expected to be applied for the ICT. Unfortunately, the performance of the USA manufactured ADBio test was problematic (**Figure 2D, Table S8A-C**) when compared in the Quindio Reference laboratory with Vidas IgG and IgM reference tests [47–50]. This is similar to our earlier results with this test with French (unpublished) and USA [27] sera. For the Colombian sera specifically, there was a marked difference of the ICT and combined detection of IgG and IgM antibodies: The AdBio test resulted in lower sensitivity for IgM in stored samples from a biobank. ICT combined simultaneous detection of IgG and IgM can improve sensitivity for IgM because most of the IgM sera used for sensitivity analysis already have IgG [26, 27, 30} and the mechanism of the test with antigen coating the black bead reacting with both quadra/pentavalent IgM and bivalent IgG which react with the antigen placed in the line on the nitrocellulose. This combined detection of different isotypes also contributes to better specificity. The lysate antigen used in the ICT contains many proteins. The Western blot can accurately discriminate between and recognize IgG and IgM specific for *T.gondii*, as can the combination of other tests such as the Sabin Feldman Dye test which also detects IgG and IgM and the double sandwhich IgM ELISA or the IgM ISAGA. The IgM ISAGA is more sensitive and thus preferable for use for infants.

In the context of clinical protocols for prenatal *Toxoplasma* screening, the ICT insures that far fewer “false alarms” are generated and that less time and resources are spent on confirmatory testing for a pregnant woman who shows an isolated positive *Toxoplasma* IgM test. Risk that such sample may be a false negative IgM from the ICT test is very low, but cannot be excluded. To avoid any risk the patient should be retested for IgG and IgM 2-3 weeks later to ensure that IgG did not appear. This happens as part of a systematic gestational monthly screening program (**Figure 4, Table S6**). It should be emphasized again that POC tests for anti-*Toxoplasma* IgG and IgM, such as the ICT, are merely a first step toward diagnosis, given that IgM antibodies can persist for up to several years after acute infection. For any woman who receives a false positive IgM test result, the next step of an evaluation with other tests can involve weeks of waiting for a blood sample to be tested using technology that runs at much higher costs than the point of care test [1, 2, 6, 7, 27, 33, 35].

A potential limitation of the ICT might have been the lack of utility of the ICT using saliva (Peyron, unpublished). There is a nanogold Nirmidas test that was used with saliva, serum, and whole blood finding high sensitivity and specificity and dye test precision for the detection of IgG and IgM [37]. We had suggested earlier this might be an ideal test to use before conception. Although finger stick for glucose is standard, easy, and familiar in obstetrical practices, obtaining saliva may be viewed as less difficult than whole blood. Thus, some view saliva could be a potential advantage. However, the nanogold has required transport, associated delays to reach a clinical laboratory, and electricity and a sophisticated machine for testing. Recently manufacture of this nanogold test was discontinued. Nirmidas has also used a gold bead ICT for SARS CoVi2 but nothing like this has been produced for *Toxoplasma* to date. The diagnosis and management of *Toxoplasma* infection best involves knowledgeable health care provider input urgently making the advantage of home testing saliva less.

Testing before conception to identify seropositive persons and then testing regularly monthly through pregnancy for those who are IgG seronegative initially would be ideal as it helps to obviate problems of anxiety provoking delays that can result in irreversible fetal damage, as well as false positive test results. Such damage in congenital toxoplasmosis, as well as in ocular toxoplasmosis can occur in very short times of less than a week, making diagnosis and initiation of treatment urgent and emergent. Minimizing the likelihood of false positive IgM while maintaining maximum sensitivity is a top priority for any point of care test candidate.

The ICT also should be very useful in clinical laboratories testing with sera with a potential false positive IgM result without IgG as described herein. It could function as a second-line test to confirm or find IgM specific for *T. gondii* is not present before sending the sample to a reference center, while continuing to follow the patient while awaiting Reference laboratory results. This is a major advance as this will save time and reduce the need for gold standard tests. It can help reduce concern for patients and physicians.

When the ICT test is used initially with whole blood the only predicate test for confirmation needed will be if the whole blood test is positive. ICT not only helped to obviate the problems with false positives but also can result in detection of true positives and very early seroconversion as described herein and also recently acquired infections described elsewhere [9, 23, 24, 26, 27, 30]. We placed this work in the context of ongoing problems for healthcare (**Commentary Figures S7, S 8**) and potential for direct and spillover benefit for the care of pregnant women and their families (**Commentary Table S9,** [14]). We also placed these studies 1 to 12 herein in a historical context building parts of a toolbox working toward a role of screening using WHO ASSURED criteria compatible tests in the elimination of congenital toxoplasmosis (**Commentary Figure S7)**. There also are a variety of other clinical and epidemiologic circumstances where knowing *T.gondii* serologic antibody status can be of considerable clinical and public health utility and benefit [1, 2, 6, 7, 27, 32–9, 46–50]. Very high-quality, low-cost screening tests such as the ICT can improve infectious diseases care in gestation, help to eliminate perinatal infections with considerable spill over benefit for health care for pregnant women and in other clinical and research settings.

Congenital toxoplasmosis is a treatable and preventable disease, and physicians and other obstetrical providers now have the tools, in-hand, to improve outcomes and reduce patient and familial suffering. This screening, the standard of care in other countries, is now increasingly feasible in countries like the United States, where the primary argument against screening has been its economic burden. In the development of this test and other high-functioning point-of-care tests, there is potential for transformation in the provision of obstetrical care to improve maternal-child health. These benefits are amplified in subpopulation demographics in the USA[28] and regions of the world where the burden of disease is even higher. Examples of this occur in the Lancaster Amish population in the USA [8, 12, 32, 46], parts of Florida, are likely in other US subpopulations [28], and occur in Central [36] and South America [47], and parts of Africa [29].

Use of the ICT for the Cincinnati maternal cohort study found ICT to be efficient (**Study 9**). Due to the small proportion who were seropositive, we were unable to test for any clustering by known risk factors for exposure: none of the individual socio-economic or location factors in a regression analysis achieved statistical significance. Future analyses with a larger overall sample size will be needed to evaluate risk factors in this population. The reasons for the relatively low prevalence in Cincinnati in this cohort remain to be discovered. We have cared for children with congenital toxoplasmosis in Cincinnati, thus, even with the low prevalence found, it is likely still that gestational screening would be worthwhile.

Our recent study in Colombia also demonstrated high acceptability of a single use of the POC on a large scale of 783 women and 30 providers [47]. Although *Toxoplasma* infections occur in all demographics it was a particular problem in those who had lower education and socioeconomic status [31, 47]. To understand risk factors during gestation and develop programs to prevent such infection will require monthly screening in areas of high to low prevalence.

The implementation of this study in the clinical trial and the QI limit of detection study demonstrated that it should be easy to introduce this test into obstetrical or other practice with little time or inconvenience. For example, when patients are evaluated for vital signs, blood pressure, glucose including by fingerstick, by medical assistant or nurse, the cassette can also be brought to obstetrician or other health care practitioner for additional reading and entry into the medical record. Photography using an I phone for documentation could easily be included into the process for additional documentation made available to patient and in the medical record.

In France screening was mandated by law. In Austria those screened received additional health care benefits. In Colombia it was introduced through practice societies. In the USA those in advisory positions recommended that education, easy feasibility, low cost would result in those who would benefit choosing to have testing incorporated in medical practice and USA patient culture at many levels by personal preference. The acceptability study demonstrated that informed patients would want this and obstetricians could use this comfortably and without inconvenience in their practice. It could easily be introduced into family practice and adolescent pediatric care to identify seropositive patients at risk of this most common form of retina disease and loss of sight. Such screening in adolescence could also provide pre-pregnancy testing for young women to allow knowledge of who is seronegative and should be screened during pregnancy. Pre-marital/conception screening as initially occurred in France could also be helpful as families plan to have children. As *Toxoplasma* has been transmitted by organ donation and white blood cell transfusion and by sperm in domestic non-human animals, and can relapse with immune suppression and may be causative for epilepsy, and some neurodegeneration, there are a number of other medical settings where knowledge of *Toxoplasma* serologic status may be useful.

Obstetricians, nurse midwives, family practitioners, obstetrical nurses, and other obstetrical providers are uniquely positioned to intervene to prevent this disease, to improve the health of both mother and child. POC test-based monthly gestational screening of seronegative patients for *T. gondii* infection provides a valuable tool in the obstetric armamentarium to ensure maternal-child wellness. When such tests have undergone appropriate evaluation by the FDA and CLIA, as they have undergone in the CE-mark evaluation and approval in Europe, this testing can enable a paradigm shift in our management of the risks associated with exposure to *T. gondii*.

### Funding, Acknowledgements, Disclosures and Insuring Objectivity in Results

LDBio Diagnostics provided the ICT and Western Blots used in the studies. ANNAR Labs (Colombia) donated the AdBio kits. For the predicate test, costs for the comparison test for 58 persons for the feasibility, clinical trial the cost of performing the Biorad IgG and IgM tests was provided by the Susan and Richard family Kiphardt Seed Fund and The Thrasher Children’s Charity. At LDBio Diagnostics, Denis Limonne Pharm D. is the scientist and CEO share holder and Raphael Piarroux PharmD, PhD was the R&D Director Scientist until January 13, 2023. A patent application was submitted by D. Limonne with the scientists at the University of Chicago and in France in August 2018. This application is pending review in the United States in accordance with US Bayh Dole laws. This is for the development of the whole blood point of care test and the practical clinical utility of the ICT to guide treatment for gestational infection to prevent congenital toxoplasmosis. This is to insure its continued high-quality performance and reproducibility of the results described herein. It is pending in review at the US patent office.

In this collaborative work, the scientists D. Limonne and R. Piarroux (DL.RP) at LDBio provided insights and knowledge from their earlier work in creating the ICT, and collaboratively with RMc discussed FDA and CLIA requirements with RMc and the FDA during an IDE and “presubQ” phase of this study. In this phase, the FDA Program provided guidance for this academic /Biotek collaboration to prepare materials to allow FDA review for dual 510K clearance and CLIA waiver for use of the ICT in the USA. RP of LDBio performed the analysis of the French Blood bank serum to establish that the correct dilution required by CLIA instructions was 1:89. DL and RP designed the instruction sheet with input from FDA, CLIA, and RMc to be tested in the “QI at limits of detection study”. This was perfected in the “presubQ” process with advice from the FDA and CLIA as the FDA indicated that a 510K clearance and dual CLIA waiver might be the appropriate application mechanism. The scientists at LDBio did not interfere with the performing of the tests, the recording, interpretation of the results nor the reported conclusions of any work at any academic site. All these studies were performed independently in the academic centers. There was no payment to the scientists. At Hôpital Bichat, Paris and in Morocco studies were/are being reported separately. LDBio did provide resources to support operating expenses and reagents, but not in the USA or Colombia. RP and DL participated in editing initial and final drafts of the manuscript. The ICT tests of the Cincinnati samples and three western blots for Chicago samples were performed at LDBio. We gratefully acknowledge all participants in this work and those at the FDA who worked with us in the “Pre-Sub Q process” recognizing the substantial potential humanitarian benefit of the work toward obtaining FDA clearance and CLIA waiver that could allow this work and test to be used to help people and prevent suffering and loss of life, sight, cognition, and motor function, while saving costs for health care.

Additional thanks for funding from the Medical Student Award, the National Institute of Diabetes and Digestive and Kidney Diseases for their Grant #T35DK062719-30, the National Institutes of Health for their Division of Microbiology and Infectious Diseases Grant to RMc #R01 AI2753,RO1 16945, AI08749-01A1 BIOL-3, U01 AI77887, U01 AI082180, TMP R01-AI071319, the Thrasher Children’s Charity Research Fund for their E.W. “Al” Thrasher Award, the Kiphart Global-Local Health Seed Fund Award (to RMc), University of Chicago. We are grateful to Taking out Toxo, Network for Good, Toxoplasmosis Research Institute, The Cornwell Mann Family Foundation, The Rodriguez family, The Samuel family and Running for Fin, the Morel, Rooney, Mussalami, Kapnick, Taub, Engel, Harris, Drago,Longfellow/Van Dusen families, and the study participants. We thank testers E.McLeod, C.Weber, F.Goldenberg, C. Guinnette, R. Tennant, Z. Williams, H.Taylor, A. Beem, and S.Syed and photographers E.McLeod and K El Bissati for their reading and photography in the limit of detection, QI study. We thank all the participants in these studies of the ICT and those in the IRB office, members on the IRB, Office of Clinical Research, at the FDA and CLIA for their guidance and A. Ponsler at the Thrasher Foundation for emphasizing the importance of these studies on reviewing the initial data at a site visit at Stanford University and the Remington Serology laboratory with RMc.

## Author Contributions

The data were collected and analyzed at The University of Chicago and at the Institut des Agents Infectieux, Hôpital de la Croix-Rousse, Lyon, France, Hôpital Bichat in Paris France, and the Reference laboratory in Quindio, Armenia, Colombia, by all authors from these institutions. MAS, SC, NB and KW prepared parts of the manuscript addressing demographics and seroprevalence in the Cincinnati cohort. RMc, RP, AG, YZ, JL, JG, and M Wallon wrote the manuscript and all authors contributed to final editing of the manuscript.

**References are listed in the Supplemental**

## Supplemental

### Preface, Explanatory note

In this supplemental there are full methods to enable replication and thorough understanding of the reagents and conceptual and practical approaches, and detailed results with primary data to complement abbreviated main manuscript. There is also a commentary about historical perspective, current problems in care, and what the future could hold if the studies herein enable the programs our work supports. This small closing part of the Discussion is entitled: “Commentary: Where have we been, where are we now and what the future might/could be in prevention of congenital toxoplasmosis?” This is a minor modification of the original manuscript incorporating all suggested minor modifications to improve clarity and to make it briefer. To make context easily available in the Supplemental, we provide the same introduction and discussion as in the abbreviated main manuscript, but with an index and some additional detail, roadmaps to the studies, figures and tables.

The Reviewers’ excellent suggestions prompted the revised short, succinct manuscript, moving some figures and tables so they are only present in Supplemental, other than original figures 6 and 7, and former tables 3,4, and 5; adding roadmap and visual summary figure. A key shows renumbering of figures and tables in supplement and provides an index to parts of supplemental. Reviewers made these suggestions to make this important humanitarian work simple, clear to a broad audience of readers, and thereby more widely accessible. The Reviewers stated: *”Reviewer #1: The screening for Toxoplasma infection among pregnant women is of major concern regarding the health assistance and prevention of deleterious impact on the fetus/newborn. The main idea of the study presented here is of great importance, in the sense of searching for diagnostic methods that can avoid false positive/negative results allowing a better view of the infection occurrence among the population.)”. and “Reviewer #2: This study was deploying a POC lateral flow assay for Toxoplasma IgG/IgM antibody detection in clinic and as a second test in the setting of IgM positivity to rule out false positives. It seems like the LFA works well with very good sensitivity and specificity, and the proposal of a new workflow and how this fits into clinical care are good things to address. …The subject matter is incredibly important, and the overall data is good…”*

**Figure 6.**
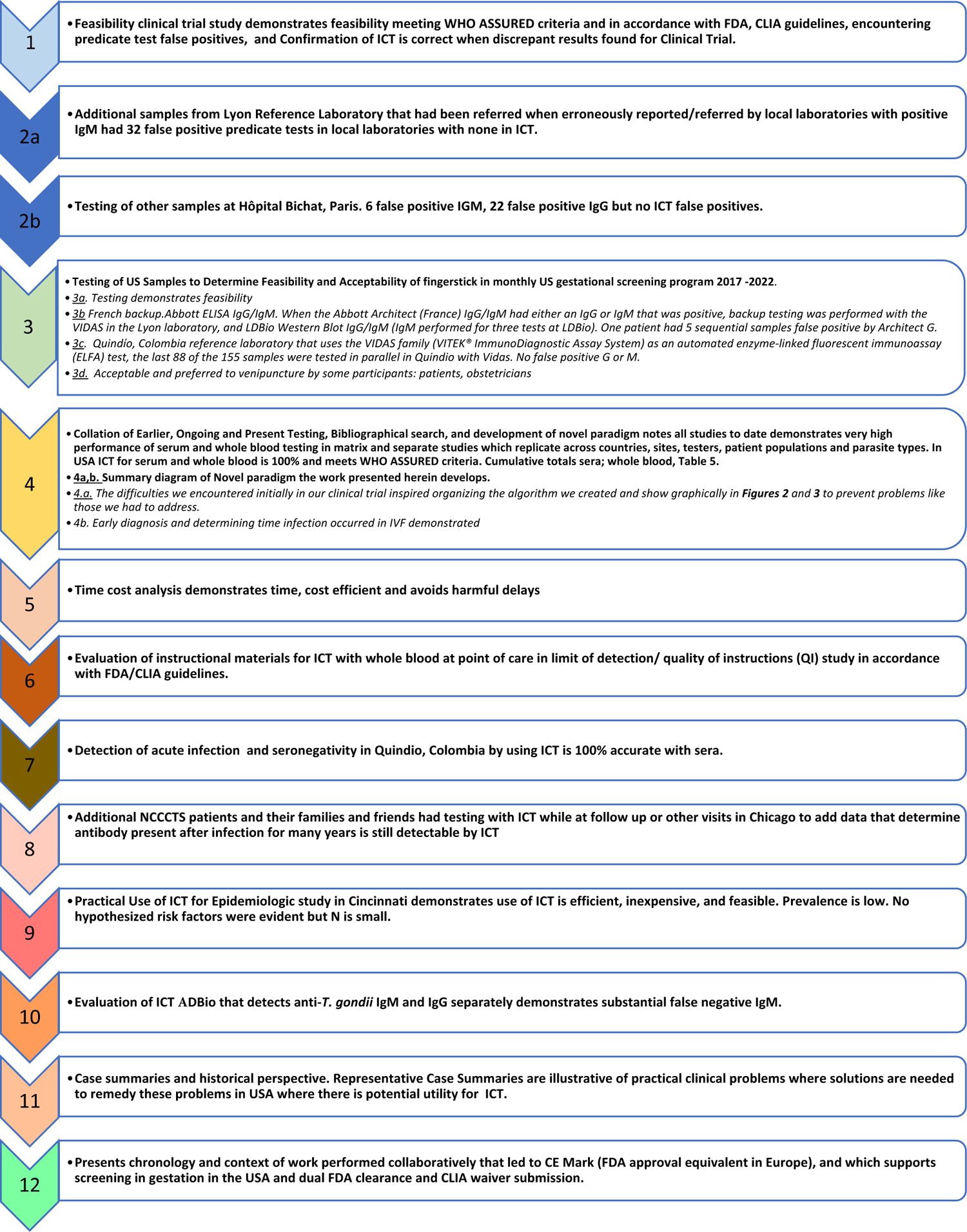
Summary Roadmap of Results of studies herein corresponding to initial Study Struccture /Design Roadmap.

We believe the suggested modifications incorporated into the more accessible, current abbreviated version are a wise approach. The Reviewers’ comments are incorporated into the Introduction in the final abbreviated manuscript. This approach also allows us to provide a “roadmap”, the experimental details, primary data, **Commentary**, historical context, and perspectives in a full supplement also part of the pre-submission process given to the FDA-CLIA program officials. This original manuscript also will be available publicly with submission of this present abbreviated manuscript and its supplement simultaneously deposited in Med-RXiv. This will allow this original manuscript to be part of the full presented to FDA and CLIA in their review anticipated soon. The goal of this work is to help enable that future where there are robust programs to prevent this debilitating and sometimes lethal/devastating disease.

## INTRODUCTION (Expanded with historical information in this Supplement, Main Manuscript is abbreviated)

It is estimated that a third to half of everyone on earth is infected with the apicomplexan parasite *Toxoplasma gondii*, with approximately 16 million people infected congenitally. Congenital toxoplasmosis (CT) occurs primarily when an expectant mother acquires the parasite for the first time during the gestation of the fetus. It can cause loss of life, chorioretinitis, loss of sight, or lifelong neurologic deficits sometimes due to hydrocephalus, and represents a considerable disease burden worldwide [1–5]. The 2013 World Health Organization (WHO) analysis of the disease estimates that there are up to 190,100 new cases of CT and up to 1.20 million disability-adjusted life years per year globally [4–7]. Disease is not evenly spread worldwide, with a concentration of disease burden in regions of the world such as Latin America and certain US populations with high exposure.

Almost all untreated congenitally infected persons have significant disease manifestations during their lives [1–17]. Considerable progress is being made toward definitive cure of *Toxoplasma* infection with novel potential medicines and vaccines to prevent toxoplasmosis [6, 7]. But, another critical part of the “toolbox” to eliminate the disease and to reduce the considerable global disease burden of CT requires prompt recognition of seroconversion and expeditious, early treatment of the acutely infected pregnant women with available, effective medicines [3,6–9]. Screening monthly, beginning before or near conception to one month post-partum for development of antibody to the parasite in previously seronegative women can enable treatment to prevent trans-placental transmission of newly acquired maternal *Toxoplasma* infection or treat the fetus to prevent sequelae [3,7–17]. France, Austria, and Slovenia have led the way in implementing nationwide, mandatory screening to this end. Colombia, Panama, Brazil, and other countries in Latin America, and Morocco are currently working to replicate efforts of these European countries [8,10, 11]. Were prevention and prompt treatment of CT a top priority of global health programs, it would lessen *Toxoplasma*’s impact on the world’s population to reduce the most severe effects this parasite can have on individuals and their communities throughout the world [8].

In the United States, there are no mandatory gestational screening protocols for CT, and treatment most often begins only once clinical signs are detected in a fetus or infant [3,6,7,8,12–14]. Of the many factors that explain this global disparity in CT screening, the cost-effectiveness of screening programs is one of the most substantial barriers to implementing screening protocols worldwide [1, 13–45]. A meeting with the Illinois legislature to introduce a limited bill to have obstetricians offer information about *Toxoplasma* to their patients [39] led to the Senator committee wanting to pass that legislation but the Department of Public Health societies and another attendee suggesting cost-benefit analyses and consideration of ways to lower costs for and means to facilitate a screening program and education programs besides or rather than legislation. Thus, several studies and cost-effectiveness models were developed and results were published over the last ten years [13–18]. These analyses all have found substantial cost savings and benefits with routine testing in France, Austria, and modeling in the U.S [14–17]. For example, Prusa et al demonstrated clear cost benefit with 14-fold cost savings even in countries with low prevalence such as Austria [15] and considerably improved outcomes with screening in gestation compared with screening only at birth demonstrated in France, Colombia and on a very limited scale even in the USA [7–17]. Recently, in France recent cost /benefit for pre-natal screening and treatment compared to no screening was found [17]. Nonetheless, the current American Academy of Pediatrics guidelines had a comment that national routine gestational CT screening might still be financially unrealistic [18]. In the U.S., and other countries without robust screening, introducing prenatal screening tests that fulfill WHO ASSURED criteria (Affordable, Sensitive, Specific, User-friendly, Rapid, Robust, Equipment-free, Deliverable) will be paramount for developing a widely used paradigm to improve prevention and care for CT [13–18].

Meanwhile, a particular problem hampering testing and development of ASSURED testing is high frequency of false positive results using currently available commercial test kits for anti-*Toxoplasma* IgM [19–22]. Commercial IgM tests for *T. gondii* antibody have had an unacceptably high prevalence of false positive results, leading to a FDA advisory about these tests [19–22, 38]. Blood samples that have reports of isolated, positive IgM results typically prompt repeat-testing by more definitive tests in a Reference laboratory and in some instances, testing for *T. gondii* specific IgG weeks later in order to determine whether IgG antibodies emerge within two to three weeks. In any case, in the United States, the FDA stated that a positive result for acute infection (IgM) with a non-reference laboratory (NRL) test should be confirmed at the Palo Alto Medical Foundation /Remington Specialty *Toxoplasma* Serology Laboratory (PAMF-TSL) [22]. This referral of the samples showing positive results and its associated delays, concern for patients and their physicians, and substantial costs (more than $800 per panel of tests in the USA with additional problems for this exceptional testing from insurance denials and capitation of obstetrical health care), has been one argument against screening programs [14]. Therefore, NRL tests with high specificity and low cost are needed. The recently developed *Toxoplasma* ICT IgG-IgM test (LDBIO Diagnostic, Lyon, France, hereafter called ICT) is a promising candidate for a NRL test that satisfies ASSURED criteria [23–5].

To begin to implement USA gestational screening programs, which has currently and historically been problematic (please isee Commentary below showing those problems for care), a formal clinical trial feasibility study has taken place at the University of Chicago Medical Center (UCMC) beginning in July 2020. This clinical trial is being performed with the goal of evaluating a sufficient number of verifiable ICT results that could be used to complete the U.S. Food and Drug Administration (FDA) 510(k) clearance and Clinical Laboratory Improvement Amendments (CLIA) regulations waiver process. This study involves comparing results of the ICT to an already cleared serum test, also called predicate test (Bio-Rad Platelia Toxo Enzyme Immunoassay). When we encountered difficulties with false positive IgM results with the predicate comparator, but not the ICT, we were faced with the unanticipated constraint of cost of positive confirmation of a number of tests at PAMF-TSL, and recognition that this type of cost from the frequent false positive IgM results could de-rail screening programs in the United States. Given the true negative with ICT in our setting, we queried whether the ICT could be part of a paradigm to rule-out false positive IgM results with NRL test, both at the point-of-care and in the hospital clinical laboratory. Further, we tested samples with the ICT that were suspected false positives from local laboratories that had been referred to reference laboratories. We placed these data in the context of practical clinical problems we encountered and collated our results with ongoing and reported similar studies to define whether this could be a paradigm helpful in addressing false positive predicate NRL test results. Solving this problem emphasized how the ICT can be used in screening programs to benefit pregnant women and their families, creating a new paradigm to approach the problem of need for accurate screening and of false positive tests. This highly accurate test may help enable screening for acquisition of *T. gondii* in gestation [23–39] and thereby contribute to saving sight, cognition, motor function and lives and improve quality of life [1-10, 12-15, 17, 24, 35].

### DETAILED METHODS (expanded from Main Manuscript Methods) Hypothesis

Our hypothesis was that a lateral immunochromatography test (ICT) that we had previously found to be sensitive and specific could meet criteria specified by the US Food and Drug Administration and Clinical Laboratory Improvement Amendments (CLIA) to document that this test is useful for serologic and whole blood point of care testing to detect *Toxoplasma* infection.

### Roadmap to overall structure and design of the work in **Figure 1**

We performed a series of studies (**Figure 1**) to test this hypothesis in the USA, France, and Colombia. In so doing, we discovered paradigm shifting approaches and utility, proving and extending beyond our original hypothesis, in studies using the methods that follow. **Figure 1** lists and provides a roadmap to the 12 studies and analyses that comprise this work:

### Study 1: Feasibility clinical trial study

The design of this Study 1 (Clinical_Trials.gov number NCT04474132) and how it is related to earlier work is shown in

### Supplemental Figure S1A-E

Serologic samples for the UCMC feasibility study (ongoing as of December 2022) were obtained from 41 pregnant women, 40 undergoing regular prenatal appointments at the UCMC (23 in first trimester, 12 in second trimester, four ithird-trimester) and from seventeen non-pregnant volunteers. Each subject’s whole blood and sera were tested with ICT; subjects’ sera were also tested at the UCMC’s CLIA-approved Clinical Laboratory, which uses a Bio-Rad Platelia Toxo Enzyme Immunoassay as its FDA-cleared standard predicate test for IgG and IgM *Toxoplasma* antibodies.

As the FDA guidelines specified, this was carried out to determine whether this new ICT could be used successfully within formal, usually structured, clinical care systems in a University outpatient obstetrical and in-field practice settings. It was compared with the simultaneously performed FDA cleared, standard, predicate test. The ICT with whole blood was read by three readers independently and with sera by two independent readers also at the same times. Three different healthcare providers performed the point of care (POC) test. Each provider tested a minimum of five sero-positive persons. Results were entered into the University hospital EPIC medical record, standard clinical laboratory, and Clinical Research Redcap systems of the University of Chicago Hospitals with ICT results also documented by smart phone photograph.

This was tested to determine whether this novel, simple test could perform well even during a time with the constraints posed by the SARS CoVi2 pandemic. As the FDA clearance, CLIA waiver and Institutional Review Board (IRB) processes specified must occur, this was a formal Clinical Trial included in Clinical_Trials.gov NCT04474132, that would mimic ways the test would be used to identify serologic positivity or seroconversion during pregnancy in the University of ChicagoMedicine practice or in other clinical settings that would be suitable to apply for dual FDA clearance and CLIA waiver. This was to formally determine whether this test could perform well and be feasible and easy to use within a clinical trial in a standard obstetrical and other care and research settings in accordance with FDA and CLIA requirements. We were expected to use a standard, comparator, “predicate” test that the FDA had already cleared for use with serum in the USA to detect and confirm presence of antibodies to *T.gondii*. There were only three such cleared tests in the USA and we selected the test our clinical laboratory utilized regularly, herein designated “predicate comparator test” so it could exactly replicate the academic practice setting in accordance with FDA/CLIA guidelines. The test cost was lowered for the clinical trial from $650 for the IgG and IgM clinical laboratory test to $13 to facilitate the study. The cost of the predicate testing was paid for by the Seed Fund Award from the Susan and Richard Kiphart Family Foundation, the A.K. Thrasher Children’s Charity and other charitable sources of funds. US Reference laboratory confirmation of test results could not be subsidized and remained over $1600 for two consecutive tests with a full “adult panel” when a positive IgM test was identified. Back up testing was performed for two patients in the Remington *Toxoplasma* Specialty Reference Laboratory and also was performed in the Lyon reference laboratory without charge using the Abbot ELISA IgM/IgG. In August 2022 these Abbott reagents also became FDA cleared.

### Whole blood sample testing protocol for ICT

For each ICT, ∼30 µL of whole blood were collected using a 60µl glass micro hematocrit tube filled to half full (by visual estimate) and placed into the well of the device. After placing four drops of diluent buffer in the well and waiting for 20 minutes, each ICT test yielded results that were interpreted by the tester and two additional readers independently reading photographs of the results. All individuals who interpreted results were blinded to the Bio-Rad IgG and IgM test results. Test interpreters determined whether the ICT result was positive (black, positive line and blue, positive control line) or negative (absence of the black, positive line and presence of blue, positive control line). According to the test Instructions for Users (IFUs) its performances are 97.5% sensitivity and 99.7% specificity when used with whole blood.

### Predicate test protocol

Platelia *Toxoplasma* IgG or IgM (Bio-Rad) (Bioplex 2020 automated assay) detects IgG or IgM antibodies against *T.gondii* via capture of IgM in solid phase. Microplate wells coated with anti-human antibody chains, a mixture of antigens and monoclonal anti-*T. gondii* antibody labeled with peroxidase, the conjugate, are used. Values ranging from 0.8 to 1 were “equivocal”, values > 1 were considered positive for IgM. These tests were used in accordance with the manufacturer’s instructions. When the first false positive IgM tests results were detected Biorad validated proper functioning of the test materials and automated machine at UCMC and re-calibrated and re-standardized the machine. This did not eliminate the false positive results which seemed to be associated with the reagents and system intrinsically.

### Confirmation of discrepant results for Clinical Trial

All discrepant results between ICT and predicate were sent to Remington Specialty Laboratory-PAMF-TSF or Lyon Reference laboratory for confirmation immediately using a panel of tests described elsewhere [27, 37].

### Study 2a: Additional samples from Lyon Reference Laboratory that had been referred when erroneously reported/referred by local laboratories with positive IgM

A set of 32 samples obtained at the Parasitology Laboratory of the University Hospital of Lyon, France (Institut des agents infectieux, Hôpital de la Croix-Rousse, Lyon, France) were selected for being reported as false positive with at least one first-line, NRL automated assay and confirmed to be negative by a panel of additional tests in the laboratory and were additionally tested at LDbio Diagnostics using ICT and WB ToxoII IgG and IgM [40–45].

### Study 2b. Testing of other samples at Hôpital Bichat, Paris

A total of 558 US serum samples that would otherwise be discarded were tested at Hôpital Bichat.,Paris (Bichat-Claude Bernard Hôpital, Laboratory of Parasitologie, Paris, France [30]). Another set of samples also was tested at Hôpital Bichat in Paris. Results are being presented in detail in a separate report describing a variety of tests from Hôpital Bichat (Abraham et al, 30, and in submission 2023).

### Study 3: Testing of US Samples in a study to Determine Feasibility and Acceptability of fingerstick in monthly US gestational screening program 2017-2022

#### Practice Setting and Patient Recruitment

This separate study was to determine whether this ICT testing could be performed monthly for pregnant women in an academic obstetrical setting in the USA and whether it would be acceptable for patients and their physicians. This study took place in the outpatient Obstetrics and Gynecology Practice at an urban academic medical center between September, 2017 and September 2018. Patients were identified at their first outpatient obstetrical visit, between 8-12 weeks gestation, by their primary obstetrical care provider. Patients not infrequently attended their obstetrical visit with their partners.They were provided an educationl pamphlet [33] and were able to ask any questions. Patients then were offered an opportunity to participate in the monthly screening study and if they wished to do so to sign an informed consent. If the patient indicated interest in participating in the study, voluntary consent was obtained by the research team. All patients who were asked expressed interest and willingness to participate. The original intent of the study was to follow 20 women to term with monthly testing through the sixth week post-partum obstetrical visit.

#### Testing

Each month, at the patients’ regularly scheduled appointment or shortly thereafter the patient was tested with the whole blood-variant *Toxoplasma* ICT IgG-IgM POC test. Methods for testing have been discussed in our previous work [23] and above. Briefly, the patient underwent finger prick and 30 µL of whole blood was collected via capillary tube. This sample equivalent to half the capillary tube volume was applied to a test kit, followed by 4 drops of provided eluent. The test was interpreted between 20-30 minutes after the sample was applied, and photographed for later, blinded interpretation. At the time of sampling via fingerprick, the patient also provided a sample of blood in a red-top tube for serum separation obtained via venipuncture. This was at the time of venipuncture for routine obstetrical care or when no such sampling occurred it was performed only for this study. Serum was tested with another high-functioning test, i.e.,with the ARCHITECT-, and /or VIDAS (VITEK® ImmunoDiagnostic Assay System) as an automated enzyme-linked fluorescent immunoassay (ELFA) and/or Western Blot-Toxo-IgG and IgM systems (LDBio diagnostics) performed in Lyon, France and/or Quindio Colombia Reference Laboratories[10, 25]. Testing was conducted on a monthly basis throughout gestation after enrollment and was completed with a final screening at the 6-week postpartum visit. A video of test performance is available at Youtube link [23 supplement]. We also tested an additional 25 participants in the National Collaborative Congenital Toxoplasmosis Study (NCCCTS) and our other studies during this time frame who wanted to participate.

### Provider Participation and Patient Satisfaction Surveys

Providers joined the study as collaborators following an Obstetrics Department Grand Rounds and Obstetrics Sectional Educational informational meeting for those who missed the Grand Rounds. Both informational meetings were presented by RMc. Providers were provided the same educational pamphlet that their patients also received. All had the opportunity to ask questions of RMc. As described above under “Patient Recruitment”, providers then mentioned the study to their patients. At the initial and subsequent visits the medical student (JL), Maternal Fetal Medicine Nurse (KL) or PI (RMc) obtained the samples after coordinating with the patient and practitioner at the time of a subsequent monthly obstetrical visit. Providers were told the results they could discuss with their patients.

Surveys, designed to assess patient satisfaction with the gestational screening program were created to use at the end of the study. Responses were based on a 5-point Likert scale, ranging from “strongly disagree” to “strongly agree.” There was also an opportunity for free response regarding strengths and potential areas of improvement for the screening program. The detail of questions is in a figure in the results section. Surveys were provided by the research nurse or others working in the study to the study participants at the 6-week postpartum visit or shortly before this visit. Contact with provision of the brief questionnaire was missed for five study participants at the 6-week postpartum visit. All five were asked and one of those five completed the questionnaire at a later time. Surveys were anonymized, so correlation of survey data to individual respondents was not possible.

In addition, as part of this separate study examining feasibility and acceptability of the ICT in this monthly screening program, we also tested these USA sera we knew to be negative with ICT with additional tests in reference laboratories in Lyon France and Quindio, Colombia. As part of this intent-to-study, as above, we had enrolled 22 pregnant women, and 19 continued monthly. In the Lyon, France reference laboratory, the 155 samples were tested with the Abbott ELISA IgG/IgM. When the Abbott Architect (France) IgG/IgM had either an IgG or IgM that was positive, backup testing was performed with the VIDAS in the Lyon laboratory, and LDBio Western Blot IgG/IgM (IgM performed for three tests at LDBio). In the Quindío, Colombia reference laboratory that uses the VIDAS family (VITEK® ImmunoDiagnostic Assay System) as an automated enzyme-linked fluorescent immunoassay (ELFA) test, the last 88 of the 155 samples were tested in parallel in Quindio with Vidas.

### Study 4. Collation of Earlier Testing, Bibliographical search, and development of novel paradigm

We collated results of all of our earlier work, both published already [23-27, 29-31, 35] and other separate studies ongoing at present on related topics that are being separately submitted for publication currently [Abraham et al, in submission 2023; 30], and our current work herein.

Now that the ICT is CE marked and available in a Europe, to determine whether we had overlooked any other study we were not otherwise aware of, we performed a bibliographical search on Pubmed using terms for evaluations of the *Toxoplasma* ICT IgG-IgM test. This was to make certain that we had included all reported tests in our analysis. Only English literature was reviewed. For evaluations found, full text was retrieved and searched for potential false positive samples.

Additionally, results of evaluations presented in congress that were known by the authors including those in submission to other journals were added to the totals in this analysis.

The earlier and ongoing studies and reports were arranged chronologically and the total collated data are reported herein. [For Reviewers and Editors manuscripts in submission that add to this cumulative total are provided to confirm entries in **Table S4** but they will be published elsewhere].

### Summary diagram of Novel paradigm the work presented herein develops

The difficulties we encountered initially in our clinical trial inspired organizing the algorithm we created and show graphically in **Figures S2.I** and **3** to prevent problems like those we had to address.

### Study/Analysis 5: Time cost analysis

We found a number of approaches including reference laboratory tests with varying costs, ease of use, and considered whether they meet WHO ASSURED criteria. Advantages and disadvantages of those approaches are summarized in tabular format including a time cost analysis.

### Study 6: Evaluation of instructional materials for ICT with whole blood at point of care in limit of detection/ quality of instructions (QI) study in accordance with FDA/CLIA guidelines

The following study was performed to determine the precision of the ICT with samples at the limit of detection and whether never experienced testers could read, understand, perform and interpret instructional material for use of the ICT with whole blood. Samples were prepared in accordance with FDA/CLIA requirements and guidance for instructional material for CLIA waiver for a point of care test (Recommendations for Clinical Laboratory Improvement Amendments of 1988 [CLIA] Waiver Applications for Manufacturers of *In Vitro* Diagnostic Devices ([version of January 30, 2008 – in force and updated in 2020]). The following limits of detection, “Quick Instructions” (“QI”) study was then performed as follows: Nine testers were identified. Testers were three practicing physicians, three nurse/nurse practitioners, two medical students and one medical resident. They were not experienced with this type of ICT using whole blood or this cassette. None worked as a certified laboratory technician. They were selected to reflect categories of potential users of this test who were unfamiliar with and unskilled with this test. Each tester took the University of Chicago blood-borne pathogens and universal precautions training to work with whole blood, and their competence in understanding and using this material currently was formally documented for the study, as was IRB-required.

A serum sample that had *T.gondii* antibody, that was positive at the limits of detection for the *Toxoplasma* ICT IgG-IgM test was used as positive sample. The sample was prepared according to CLSI EP12-A2 User Protocol for Evaluation of Qualitative Test Performance guideline, with the objective of obtaining a sample that would be positive around 95% of the time (defined as positive between 36 to 39 times out of 40 trials, as per guideline instruction). The sample used was a citrate-based plasma obtained from the *Etablissement Français du Sang* (the French national blood bank) with both IgG and IgM for Toxoplasmosis (38 UI/ml of IgG and 63.89 ratio for IgM according to Roche *Toxoplasma* kits) prior dilution into seronegative heparinized blood obtained from venipuncture of a known seronegative person the day prior to dilution. The selected dilution was the 90^th^ (1:89 ratio) as the test was read positive 37 times by an untrained user and 39 by an experienced user at that dilution (LDBIO Diagnostics, data not shown). Assuming linearity of dosage for the Roche kit, the IgG titer of the final sample was therefore 0.42 UI/ml, and 0.71 ratio for IgM, both below the threshold of 3 and 1 for the Roche IgG and IgM kit, respectively.

Knowing that this limit of detection was determined to occur when this positive serum was diluted 1:89 with whole blood, serum or plasma from a seronegative donor, the study proceeded as follows: Prior to being tested by the nine testers, the whole blood from another seronegative donor in Chicago was confirmed to be negative with ICT from fingerstick. Thirty ml of whole blood was drawn from this second seronegative donor by venipuncture and placed into three ten ml tubes with EDTA anticoagulant and gently mixed by repeatedly inverting the tubes. This blood was divided into two parts, one part was to be used to create the negative samples and the second part was to be used to create the “limits of detection positive” sample. Half the blood from the seronegative donor was diluted 89:1 with the previously tested positive serum and gently mixed to create the “positive at the limits of detection whole blood sample”. The ability of an experienced tester to distinguish the negative and positive samples was tested with the ICT and was confirmed. Two hundred microliters of the negative whole blood was placed into each of sixty-three 1.5 ml Eppendorf tubes used in this study for the negative samples. Two hundred microliters of the “positive at the limits of detection whole blood sample” was placed in each of sixty-three 1.5 ml Eppendorf tubes. The samples were assigned and labeled with random numbers between 1 and 14. The code was known only to the scientist who diluted the samples and prepared the labeled tubes and created the unknowns to be tested. A set of unknown blood samples labeled from #1 to #14 was prepared for each of the nine testers along with ICT cassettes labeled with the number between 1 and 14. Each set had the testers initials on each cassette. Each tester had the same samples labeled 1 to 14. The testers were asked to read and sign the informed consent if they concurred. They were asked to read the information (Quick information, QI) document and complete the top of the data sheet documenting they had read the QI and indicate if they believed they understood its content. They also answered questions about their educational and other ICT experience. The cassettes with initials and numbers along with capillary tubes, laboratory coat, gloves, and data sheet were provided in a work space separate from other testers and with guidance from the written instructions. Informed consent, and signature into a study log were obtained.

The ability of three groups of testers with different clinical roles to read the instructions, to perform the test in accordance with the instruction, and to distinguish negative and positive results at the pre-established limits of detection were evaluated. This was done to determine whether the instructional material was adequate to teach unskilled users in three different practice type settings. Familiarity with universal precautions and biosafety instruction precautions was documented as described above. The fourteen samples, half positive and half negative were tested by each tester in a blinded manner. The tester and one other reader read the tests. The testers read the cassette at 20-30 minutes after sample followed by buffer were applied to the cassette, and recorded their interpretations on a data sheet. Photographs of the cassettes were taken using smart phones with photography in an area with some natural light. The photographs were provided to the tester to read in a blinded manner. One of the testers was a second reader for 7 of the photograph sets and a laboratory scientist was the photograph reader for the remaining two tester’s photographs including the tester who was the photographer and second reader for the first 7 testers/readers. A skilled physician scientist with familiarity with the test was a third independent photograph reader. The tester, another unskilled reader (other than the laboratory reader who was experienced), and experienced reader all independently read the photographs of the cassette in a blinded manner and their results were recorded. Data from the data sheet then were analyzed.

### Study 7. Detection of acute infection and seronegatvity in Quindio, Colombia by using ICT

Sera from 24 patients who had recently acquired their *T.gondii* infection in Quindio Colombia and 12 patients who were seronegative had sera that were tested with Vidas ELIFA IgG, IgM and with ICT.

### Study 8. Additional NCCCTS patients and their families had testing with ICT while at follow up visits in Chicago to add data determine antibody present for many years is still detectable by ICT

Between March and December 2018 20 participants in the NCCCTS whose serologic status was known from earlier reference laboratory testing and family members traveled to Chicago. While participating in their NCCCTS visit they had a fingerstick sample of whole blood, and this was applied to an ICT. The number of tests were 20 seropositive and 8 of the family members or other controls for separate studies such as multimodal neuroimaging studies were found to be seronegative but did not have other reference laboratory testing. The time from acquisition of infection was noted, in a similar approach to the earlier studies of Begeman and Lykins.

### Study 9. Use of ICT for Epidemiologic study in Cincinnati

Sera and demographic data from a maternal-infant cohort in Cincinnati were available for 265 women; 264 had data on the variables of interest. Variables of interest included residential address (longitude and latitude), age, education, race, income and pet ownership as part of the original cohort study. Sera were tested with ICT and if positive then were tested with IgM and IgG western blots at LDBio.

A logistic regression model on the results for the 264 samples was used to estimate the ICT *Toxoplasma* infection positive status by including independent variables such as, maternal age, marital status, Neighborhood Deprivation Score (a higher value means more deprived and missing values are extrapolated from 5 nearest neighbors), latitude, longitude, race, i.e., White or not, and an interaction term between maternal age and Marital status.

### Study 10. Evaluation of lateral chromatography test AdBio that detects anti-*T. gondii* IgM and IgG separately

To determine whether a USA made immunochromatography test (called ADBio) would function as well as the ICT or whether samples from Colombia that had very high performance with ICT would function as well with a different USA made test, an additional set of known positive IgM positive or IgM negative samples was tested. The tests that were used were the VIDAS (Quindío, Colombia) test and another commercially available but not FDA cleared or CLIA waived test that detects *Toxoplasma* specific IgM in the Colombian Reference laboratory. A total of 147 serum samples were included, selected from the biobank of past studies at the University of Quindío in Armenia, Quindío, Colombia. All samples were previously tested using the reference test VIDAS (VIDAS Toxo-IgG Avidity kit; bioMérieux, Marcy-l’Etoile, France). Samples were divided into the following three groups as defined by VIDAS testing: (1) IgG negative and IgM negative (n=65), (2) IgG positive and IgM negative (n=55), and (3) IgG positive and IgM positive (n=27). These groups corresponded to seronegative samples, chronic *Toxoplasma* infection, and acute *Toxoplasma* infection, respectively. VIDAS® TOXO IgM (TXM) assay, is an enzyme-linked fluorescent immunoassay-ELFA-(Biomerieux, Marcy-l’Étoile, France) performed in an automated instrument. In VIDAS IgM test, a value is generated for each sample by forming a ratio from the relative fluorescence of the sample to that of the calibrator or a stored calibrator result (“stored standard”). Test values from patient and control samples are compared to a set of thresholds stored in the computer. The thresholds and interpretations are given as follows according to the values of the cutoff index (COI): <0,8 COI = non-reactive; 0,8≤ COI <1,0 = gray zone and ≥1,0 COI = reactive. The AdBio Test (CTK-Biotech, San Diego, CA) uses lateral-flow immunochromatography with a test and control band and goes further with separate bands to differentiate between *Toxoplasma*-IgG and *Toxoplasma-*IgM antibodies. Tests were performed and results were read according to manufacturer guidelines and confirmed by three investigators. Sensitivity, specificity, positive predicted values (PPVs), and negative predicted values (NPVs) were calculated against VIDAS IgG and IgM test results, respectively, using the Vassar Stats Clinical Calculator (http://vassarstats.net/clin1.html).

### Analysis 11: Representative Case Summaries illustrative of practical clinical problems where solutions are needed and potential utility of ICT

Representative case vignettes with concepts they illustrate were collected and were summarized to illustrate impact and need of this paradigm and its historical context (called “**Study 10”**). These brief case summaries are from The National Collaborative Chicago-Based Congenital Toxoplasmosis Study and Consultations to the Toxoplasmosis Center and Toxoplasmosis Research Institute. They illustrate representative, frequent clinical problems incurred from false positive IgM tests. Representative examples of benefit and novel utility of ICT in addressing this problem are also presented. This was to illustrate the substantive problems false positive *Toxoplasma* antibody serologies continue to cause in an ongoing manner in local laboratories in the USA, and to place the present study in the context of practical problems in clinical care. They are presented to illustrate the difficulties and grave consequences of erroneously false positive and negative results. A case summary also presents use of ICT for early detection confirming pre-conception infection when sequential samples obtained in the context of *in vitro* fertilization (IVF) were available. **Commentaries** about screening programs and their absence in the USA were also identified to further place our findings in an historical perspective.

#### Ethics

The ongoing UCMC study, under the name “Prevention of CT: Feasibility of prenatal screening using point-of-care tests,” is conducted with ethical standards for human experimentation established in the Declaration of Helsinki. Research received approval from the University of Chicago Institutional Review Board (University of Chicago IRB Protocols 20-0442, 19-0505, 21-1446, 8793, 8794, 8795, 8796, 8797, 8798, 16708A and met the standards of the Health Insurance Portability and Accountability Act. Results were or will be reviewed by/discussed with the Data Safety Monitoring Board. Informed consents were obtained from subjects in accordance with the University of Chicago Institutional Review Board and the National Institutes of Health guidelines. No subjects are under the age of 18. All participants provided informed, written consent prior to their participation in the study. All studies were performed in accordance with the Declaration of Helsinki.

Samples from the Lyon Reference laboratory of the University hospital were anonymized in this analysis. Testing in Colombia was performed in accordance with local Ethics Committees approvals and guidelines. Stored sera from Cincinnati was approved by the IRB for future testing for a wide range of pathogens.

## RESULTS (including details and tabulated primary data and Key to link Figure and Tables in Main manuscript and Supplement)

Key showing Correspondence of Main Manuscript and Supplemental Figures and Tables with study numbers are in the Key below. In this version of the supplemental all figures are included when cited, both main manuscript and supplemental tables and figures.:

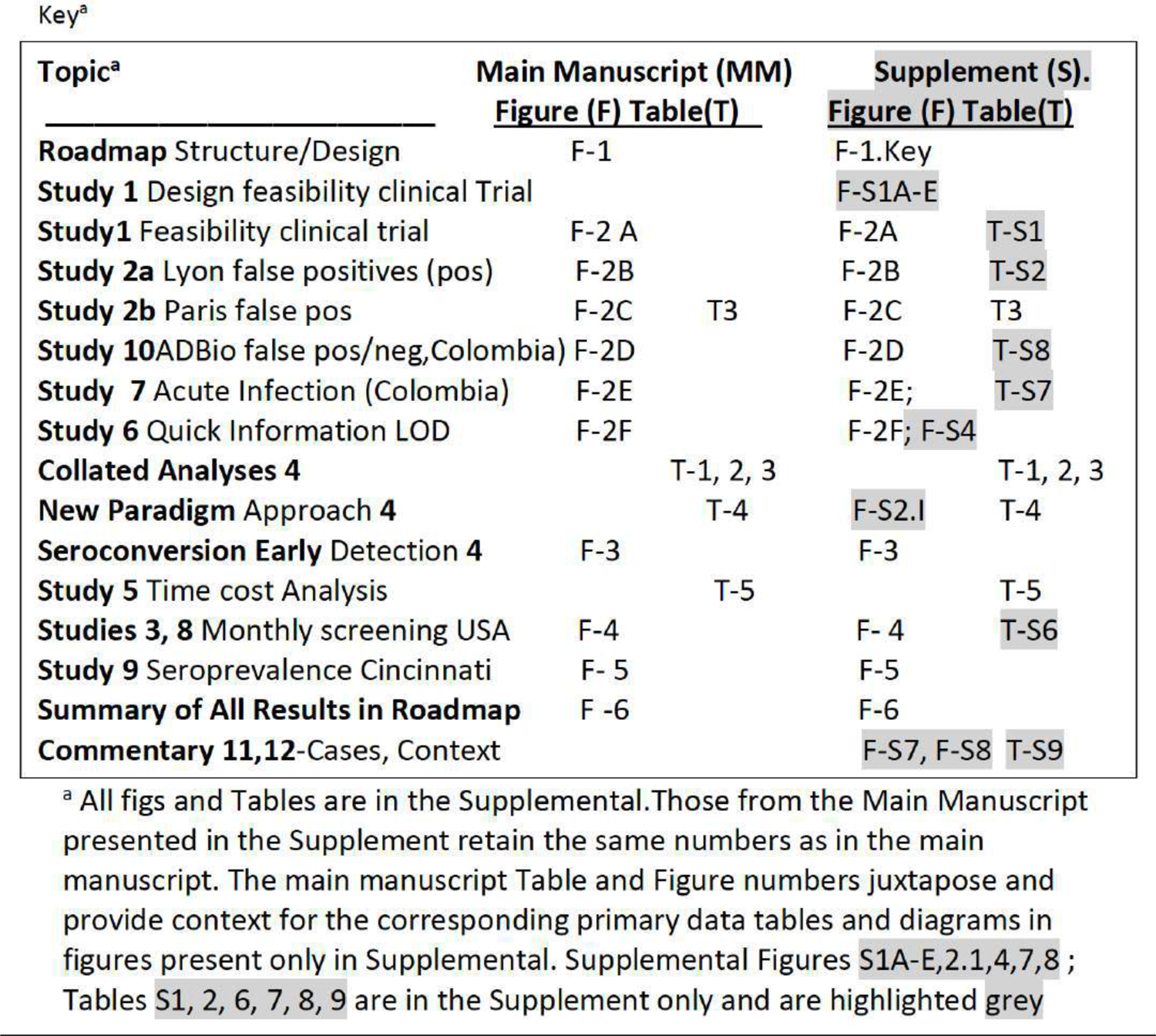

### Study 1.Feasibility clinical trial study performed exactly as the test would be used in practice, 2020 to 2021 demonstrates feasibility, identifies false positive predicate test results and develops new paradigm to help to obviate that problem

Individual results in the ICT and the predicate test in this ongoing clinical trial study are in **Table S1**. Between August 2020 and December, 2021 we performed the Clinical Trial study described herein. For this prospective clinical trial, 43 seronegative and 15 seropositive persons were tested with the ICT using whole blood and sera. The 58 sera were also tested with the predicate test used by The University of Chicago Medicine Clinical Serology laboratory, the Biorad assay. Results of the ICT always had at least two independent observers to insure objectivity in their interpretation as well as to avoid any possible bias. Readings were documented by photographs 20-30 minutes after applying the sample and the buffer immediately after the sample. RMc always read the results independently after initial readers and never influenced the readings of the initial readers. Results of all readers were uniformly concordant. Any positive results were re-tested in Reference Laboratories.

As specified by the FDA the three testers of different professional backgrounds who might be among the categories of users of the test included an obstetrician, an infectious diseases specialist, and an obstetrics maternal fetal medicine nurse. All who tested participants were blinded to the participants serologic status. Each tester tested at least 5 persons who turned out to be seropositive and at least 5 persons who turned out to be seronegative. Testing in Chicago took place in the nurses examining area, the obstetricians examining area/office and sometimes in outdoor field settings during the SARS CoVi pandemic. Testing of 265 sera obtained in a study of influenza in Cincinnati between 2017 and 2019 demonstrates that it is easy and efficient to use the ICT for epidemiologic studies. Feasibility, accuracy, and one hundred percent correlation between the whole blood point of care test, the ICT with serum performed by a laboratory member, and a predicate and when needed gold standard test were identified.

Within testing of the initial 6 pregnant participants, we encountered false positive results for two participants in the IgM predicate test and three others later revealing a false positivity rate of 10%. Finding these frequent false positive predicate test results was disturbing for patient participants, providers, and investigators. Correcting the erroneous predicate test data with follow up gold-standard testing was time consuming, costly, and results in delays in care. Although the Reference laboratories in the USA and France provide one excellent solution to the problem of false positive results, that is of high quality results, the delays, cost and inconvenience were substantial difficulties. This occurred while we as investigators could promptly see the negative ICT result in whole blood and sera testing. We knew the results in the context of our earlier data with very high performance, sensitivity, and specificity of the ICT. Our earlier work had demonstrated negative results were accurate with the ICT for erroneous predicate test results considered as positive when samples were referred to the USA reference laboratory. This occurred when 33 patients with 60 sera were tested). We recognized that if we could not solve this problem of false positive predicate tests (**Figure 2A, 2B, 2C,2D, Tables 1-3, S1, 2, 7, 8)**, this would have de-railed the research study and its longer-term goal of proper testing in systematic gestational screening programs for the USA and as a model for other countries. For the USA, along with their present high costs in the USA, false positive tests also would substantially harmfully influence the use of screening programs (**Figure 2, S2.1**).

### New paradigm elucidated by this experience

Thus, as we tested the whole blood and sera with the ICT in parallel with testing the sera with the Biorad test, we developed an easy, inexpensive paradigm shifting approach to solve this problem of false positive tests for the USA. This paradigm (**Figure 2.I**) showed the ICT could help to eliminate the problem of false positives both in the clinic and the clinical laboratory. This paradigm was to have a method for diagnosis with a test that meets WHO ASSURED criteria available promptly at the time the test was performed and to have a first backup of positive results in serum rapidly in the local laboratory. Our experience shown by **Tables 1-3, S8** demonstrates that this ICT performs properly in clinical practice and field studies. We noted that it could be used correctly by previously untrained observers, meeting WHO ASSURED criteria. This problem of false positive predicate test results occurred unexpectedly because of the frequent false positives with the predicate test we were using while preparing for FDA clearance and CLIA waiver. This was unexpected because we had earlier used US and French reference laboratory backup testing. We had incorrectly considered that the FDA clearance of the predicate test that we were required to use for this clinical trial would have addressed and solved the problem of false positives.

As we worked in this study, it was apparent that the ICT used in our research laboratory setting with the same sera also provided evidence that this test was potentially useful for clinical laboratories when we simultaneously tested sera with the ICT along with the performance of the predicate test in the clinical laboratory **(Fig. 2, Table S1**). We found that this also could help to obviate the difficulties caused when a commercial predicate test has false positive results (**Fig 2, Table S1**). This was while introducing a novel test that could be low cost and easy to use in the clinic.

It became clear from the experience and data presented in **Fig 2, S2.I, Table S1** that this novel test and paradigm could be a useful new method for the clinical laboratory to identify true positives rapidly for this emergent problem/disease.

This could help to determine whether there was need for further screening. It could help to clarify whether there was need for emergent care with life, sight, cognition saving medicine should be initiated promptly while waiting for backup reference laboratory confirmation of a true positive test (**Figure 2, S2.I**).

### Study 2.Additional testing of erroneous false positive local predicate tests with ICT and gold-standard testing in reference laboratories demonstrates utility of the novel paradigm with ICT

Then, Lyon and Paris Reference laboratories’ identified erroneous false positive results reported for samples referred for testing from local private laboratories that used commercial tests. This occurred at least once a day in each laboratory. Thirty-two samples that were referred to the Lyon reference laboratory from September 2021 to February 2022 from private laboratories because of the detection of isolated IgM in the course of monthly prenatal retesting, which the main system used in France in this context (**Table S2**): The tests that had been used included Cobas Roche (n = 21), Abbott (Architect n = 1 or Alinity n=7), Siemens (n = 3) (**Table S2**). None of the additional 32 samples gave positive results with ICT or in the reference laboratory with Abbott Architect despite the erroneous reports of positive IgM results (**Table S2**). Further, in Lyon France, none of the 32 false positive IgM tests with the predicate local laboratory tests used had false positive test results with the ICT or gold standard Western Blot (**Table S2**). This was using the same referred serum that was tested and reported to be positive from the local private laboratory. These results are included in **Tables 2** and **3.**

### False positive referrals Croix Roux, Lyon, France with ICT, Western blot IgM negative

| Patient number | Sampling date | Gestation when tested (week) | ICT results | IgM | Test kit | WB IgM results | University hospital Lyon la Croix Rousse conclusion |
| --- | --- | --- | --- | --- | --- | --- | --- |
| 1 | 9/21 | unknown | Negative | 3.84 | Cobas | Negative | Negative |
| 2 | 9/21 | 33 | Negative | 0.876 | Cobas | Negative | Negative |
| 3 | 12/21 | 34 | Negative | 3.26 | Cobas | Negative | Negative |
| 4 | 4/21 | 9 | Negative | 1.35 | Cobas | Negative | Negative |
| 5 | 12/21 | 32 | Negative | 1.19 | Roche Immunoelctro | Negative | Negative |
| 6 | 2/22 | 7 | Negative | Equivocal | Roche Electrochimiluminescence | Negative | Negative |
| 7 | 8/21 | 4 | Negative | 0.74 | Abbot Alinity | Negative | Negative |
| 8 | 9/21 | 4 | Negative | 1.3 | Roche Immunoelctro | Negative | Negative |
| 9 | 11/21 | unknown | Negative | 0.54 | Abbot Alinity | Negative | Negative |
| 10 | 12/21 | 11 | Negative | 0.98 | Cobas | Negative | Negative |
| 11 | 1/22 | 23 | Negative | 1.45 | Roche Immunoelctro | Negative | Negative |
| 12 | 9/21 | unknown | Negative | Positive | Cobas | Negative | Negative |
| 13 | 9/21 | 16 | Negative | Positive | Cobas | Negative | Negative |
| 14 | 4/21 | 11 | Negative | 0.72 | Abbot Alinity | Negative | Negative |
| 15 | 10/21 | 36 | Negative | 0.92 | Abbot Alinity | Negative | Negative |
| 16 | 2/22 | 9 | Negative | 1.44 | Roche Immunoelctro | Negative | Negative |
| 17 | 10/21 | 15 | Negative | 1.11 | Centaur | Negative | Negative |
| 18 | 10/21 | 15 | Negative | 1.68 | Cobas | Negative | Negative |
| 19 | 10/21 | 14 | Negative | 1 | Centaur | Negative | Negative |
| 20 | 10/21 | 10 | Negative | 0.867 | Cobas | Negative | Negative |
| 21 | 9/21 | 7 | Negative | 11.67 | Roche | Negative | Negative |
| 22 | 7/21 | 32 | Negative | 0.66 | Abbot Alinity | Negative | Negative |
| 23 | 8/21 | 10 | Negative | Positive | Cobas | Negative | Negative |
| 24 | 9/21 | unknown | Negative | 1.48 | Cobas | Negative | Negative |
| 25 | 8/21 | 3 | Negative | 0.6 | Abbot Alinity | Negative | Negative |
| 26 | 4/21 | 30 | Negative | 0.6 | Architect | Negative | Negative |
| 27 | 11/21 | given birth | Negative | 0.9 | Cobas | Negative | Negative |
| 28 | 8/21 | 3 | Negative | 4.14 | Cobas | Negative | Negative |
| 29 | 7/21 | post birth | Negative | 2.68 | Centaur | Negative | Negative |
| 30 | 10/21 | 17 | Negative | 1.01 | Abbot Alinity | Negative | Negative |
| 31 | 5/21 | 3 | Negative | 1.35 | Roche | Negative | Negative |
| 32 | 10/21 | 29 | Negative | Equivocal | Roche | Negative | Negative |

### Analysis 4. Placing US ICT in the context of other ongoing studies including Study 3 and previously published studies demonstrates high performance

Data summarized in **Tables 1, 2, 3** place the results in the Clinical Trial and monthly screening acceptance studies (Study 3), in the context of other ongoing studies and our earlier published work. **Table 1, 2** address details of studies in the USA. **Tables 1-3 and S2** also collate and address studies of false positive IgMs referred to reference laboratories in the USA and France. **Table 3** collates all these studies including those of other countries as a summary of all available results. Sensitivity, specificity, confidence intervals and details of studies are in **Tables 1 to 3**. Performance of the ICT is high, sensitivity ≥98.5%, specificity 98.9%(serum and/or blood). This high performance provided a foundation for a new paradigm and approach (**Table 4, Figure S2.1**).

Additional testing of a set of samples with Architect, Vidas and other tests was performed in an additional matrix analysis pertinent to consideration of false positive test results and part of Study shown in **Figure 5**. Back up testing of another set of sera from a monthly screening and acceptability of the monthly screening program was performed in the Lyon France and Quindio Armenia Colombia Reference laboratories (**Table S6**). In these reference laboratory settings as well as in the Paris Hôpital Bichat reference laboratory [30] false positive test results were less problematic (**Table S6**) than in the clinical trial. There were no false positives in the Quindio laboratory Vidas testing and one patient with multiple consecutive IgG false positives in the Lyon Architect tests.

### Bibliographical search confirms high performance and that data analysis herein includes all published studies

As the ICT is now commercially available following CE Mark approval in Europe, we also used a bibliographic search to attempt to identify results with which we might have been unfamiliar or with inferior performance of the ICT. **Table 3** details all studies performed to date: Number of persons (N), Sensitivity (Se)/ Specificity (Sp), country of samples, N of false positive IgM, ICT results on false positive IgM and confidence intervals, testing for serum and whole blood are in this Table. As the ICT is now commercially available following CE mark approval in Europe we also used a bibliographic search. To date all studies have involved the authors of this manuscript. There were no additional studies identified that have been reported to date. A total of 4606 sera, 1401 positive and 3205 negative, and 1876 whole blood tests, 728 positive and 1148 negative tests have been performed, including all published, ongoing studies and those herein with high sensitivity and specificity (**Table 3),** foundational for a novel approach for gestational screening to diagnose primary infection during hestation and therbeby enable treatment of the pregnant woman to prevent congenital toxoplasmosis **(Table 4. Figure S2.1)**.

### Overall performance of ICT with NRL tests false positive IgM also is high including in Study 2 a and b

Overall, including our own results herein, we found 137 samples with false positive IgM in at least one NRL technique also tested with ICT, among which 132 were found negative in ICT. The specificity of ICT for false positive IgM was 96.4%. Three samples were from IgG negative pregnant persons in Chicago. Two seropositive persons also had false positive NRL IgM was not found in Reference laboratory IgM tests. In addition 22 false positive IgG results were correctly identified at Bichat-Claude Bernard Hôpital, Laboratory of Parasitologie, Paris, France [30] and 5 false positive IgG identified in the predicate Abbot Architect in the University of Chicagomedicine samples tested in Institut des agents infectieux, Hôpital de la Croix-Rousse, Lyon, France.

### Study 5. Time, cost, comparing tests demonstrates time/cost savings and aids in eliminating delays

An analysis of relative time and cost is in **Table 5**. The ICT is substantially time and cost saving,

### Representative case summaries illustrate problems in care that false positive tests can cause and utility in identifying seroconversion in infection acquired prior to conception, called study 4b and “analysis 11” in methods)

**Table S9 in the Commentary in the Discussion** has brief summaries of some consequences of false positive and negative results in ongoing cases in USA clinical practices. These provide further evidence of problems that false positive test results can cause.. This contrasts the current status and consequences of CT at earlier times and continuing to the present in France and in the USA (**Analysis 11** in Commentary with Discussion). This illustrates that the ICT and gold standard back up testing can solve a substantial health care problem. This is both in a historical context and at present, with potential spillover benefit for care for pregnant women and their families.

**Study 4b (Figure 3)** shows utility in identifying infection prior to conception the approach seen in **Table 4, S7** and **Figure S2. I**. This approach presents our novel paradigm. The ICT detected seroconversion a day earlier than the Sabin Feldman Dye test and IgM ELISA in the Reference laboratory. There are a number of examples of patients who developed M alone then M and G [26]. In the Mahinc et al study [26, **Table 3**] there were 50 serum samples from 24 women for whom there were 17 samples with IgM only and 33 samples with IgM and IgG; ICT was positive for all samples except one that had a borderline IgM ISAGA of 5 for a patient who later was found to have acute *Toxoplasma* infection. There were also another 144 acutely infected persons identified in the USA, France, Morocco and Colombia all identified as positive with ICT [24, 25, 27, 29, 30] **Table 1-3, S7**. It is unusual, however, to watch seroconversion with as much precision in narrow time intervals so early in infection as in **Figure 3.**

Study 6. Testing ability of written instructional materials to facilitate clinical use of the ICT by healthcare practitioners not skilled in using the ICT in a limit of detection quick information study per FDA and CLIA instructions demonstrates high performance, Moving toward implementation, ability of written instructional material to be used in clinical practice with samples at limits of detection for positive whole blood samples and negative whole blood samples was found to have perfect performance. This perfect performance was for all the “blinded” readers and testers performance and reading after they read the Quick Information (QI) materials (**Figure 2F**, S4).

### Study 3. Feasibility and acceptibility of monthly gestational screening with ICT is demonstrated

Early in the work with the ICT in 2017 to 2018 we performed this study to determine whether monthly gestational screening would be feasible in a research study setting (**Figure 4**). Results of the testing did not enter standard medical records or the EPIC system at that time and the testing took place earlier than study 1 but was completed after that study. This study was initiated before the clinical trial and led to the initial meeting with the FDA when a program officer from the Thrasher Foundation emphasized the importance of FDA clearance for the test to be useful to help patients in the USA. We also asked participants at its completion whether participants felt it was important and comfortable to have knowledge about *Toxoplasma* in pregnancy and whether they would want serologic testing and/or the finger stick point of care test in subsequent pregnancies if it were approved in the USA. The intent was to determine whether screening might be acceptable in standard academic obstetrical USA practice: Some parts of the study, e.g., the questionnaire and additional backup testing were performed in 2020. Results for the initial tests for the participants’ visits were included in the earlier 2018 publication [23]. Thus, the numbers included for this study in the cumulative total of tests were subtracted from the tests shown in **Figure 4**.

### Patient Characteristics

Patients were all identified at between 8-12 weeks gestation. Patients had a median age of 31 years (range: 24-40 years). Seven of the 22 participants were nulliparous, while the remainder had been pregnant once or twice before. None reported having been tested for *T. gondii* infection in the past. Participants were enrolled in the study between September and November, 2017. The study initially concluded in September, 2018 with the birth of the last participant’s child. Because five mothers were missed by our study group at their 6-week postpartum visit, an anonymized questionnaire was provided in 2022 for those participants. This was considered separately in our analyses.

### Testing Details

Patients were tested at monthly intervals after their initial enrollment and tested until their 6-week postpartum visit. A small subset of patients (3/22) were withdrawn from the study: One individual underwent elective termination due to fetal anomalies. One participant suffered a spontaneous abortion. The third patient chose to withdraw from the study due to traditional beliefs about dangers associated with venipuncture. No patients (0/22) had evidence of prior infection with *T. gondii* upon their initial testing with the whole blood POC test, and none seroconverted during gestation. One participant had a faint band suggesting the possibility of a positive test on one test, but this was only visible to the naked eye and could not be independently confirmed with photography. Per manufacturer instructions, this test was interpreted as negative. There was 100% concordance between testing interpretations of the POC test and confirmatory testing, including the ARCHITECT/Vidas/Western blot systems and the serum-based POC test variant, commercially available and now CE mark approved in France. The course of gestational screening for each participant is presented in **Figure 4A**.

### Patient Perceptions of POC Gestational Screening are favorable

Initial response of patients and their families to screening was noted. No patient declined and responses were enthusiastic from patients. There were even requests from patient participants about whether other pregnant and non-pregnant family members and friends could join. For example, even some fathers asked to be tested to know their own serologic status and if they might be at risk of retinal disease if infected.

At the end of the consecutive screening tests, we administered the patient preferences survey to an available subset of the cohort (14 in total) at their 6-week postnatal visit. Those participating women who were missed completed the questionnaire in 2022. The POC testing and screening for acquisition of *T. gondii* in gestation was well received by all participants. The remaining participants 6-week visits were missed due to the lack of availability of study personnel to provide the survey at the time of the 6-week visit, but the questionnaire was provided in 2022, one participant responded and these data were considered separately because of the time elapsed. On average, patients appeared to prefer finger-stick POC screening to venipuncture, (POC-4.93 ± 0.27 vs venipuncture-4.00 ± 1.04) (**Figures 4B, 4C**). More than 90% (13/14) of enrolled patients “strongly agreed” that they would have POC testing in a future pregnancy if it did not require monthly venipuncture, and the same percentage of patients “strongly agreed” that knowledge of *T. gondii* infection and ways to avoid exposure were important for pregnant women. It should be noted that 70% (9/13) of patients either “strongly agreed” or “agreed” that they would pursue screening for *T*. *gondii* infection even if it meant monthly venipuncture. The participants who responded after the time elapsed in 2022 also agreed that screening was worthwhile but only would want to do this by fingerstick and not venipuncture.(**Figure 4C right panel**)

### Provider Perceptions of POC Gestational Screening are favorable

There was not a formal questionnaire for providers. Rather, level of interest and enthusiasm was reflected by the following: All providers remained involved in the study with their patients. Additional providers in the practice noting the ongoing study with the initial providers asked to join. Those still practicing at the University of Chicago at the later time did continue to collaborate in the subsequent clinical trial study presented herein. These objective measures documented continued involvement rather than a formal survey. All providers found the finger-stick testing and monthly screening a constructive addition to their practice. The rapidity of obtaining the results was viewed positively.

### Studies 4b, 7 and 8. ICT detects early seroconversion and distinguishes additional seropositive and seronegative samples in USA, France and Colombia

We noted, as shown in **Figure 3,** the ability of the ICT to detect very early seroconversion in a study of sera obtained at narrow intervals to monitor hormone levels during *in vitro* fertilization (IVF), that happened to occur during very early seroconversion. This was a USA patient whose *in vitro* fertilization had occurred 6 months prior to implantation of their embryo at a time that she was seronegative. In the interval between *in vitro* fertilization and implantation she had traveled to New Zealand where she likely acquired *T.gondii* infection in the weeks before implantation as shown in **Figure 3**. Our earlier studies (**Tables 1, 2** and **3**) have all identified perfect performance in detecting sera from those with acute infection. It was, therefore, of interest to determine whether the same would be found in sera from patients with acute infection with the genetically different parasites found in Colombia. **Table S7** shows perfect ability, sensitivity, and specificity of the ICT to also identify acute infection (IgG, IgM) in Colombia (n=22) and those who are seronegative (N=12), (p<0.0001)(sensitivity 100% and specificity 100%). This brought the total to 144, as above, Further, serologic status was correctly identified for additional NCCCTS participants tested with finger-stick whole blood and serum between March and December 2018 herein (N=20 positive chronically infected persons [times after infection years were known to be greater than 17 years for all except 3 persons, and 5 negative)

As we found perfect correlation of testing of whole blood obtained by fingerstick and serum testing in the United States, herein, and almost perfect correlation in Morocco this ICT using whole blood accurately distinguishes seronegative and seropositive status as occurs in seroconversion. Indeed, in the study in which we tested whole blood (that contained serum that originally had 38 UI/ml of IgG and 63.89 ratio for IgM according to Roche *<u>Toxoplasma</u>*kits, diluted 1:89 in whole blood from a seronegative donor) at the limits of detection in the “QI study”, there was high accuracy in distinguishing positive and negative samples. There were N=63 negative and 63 positive, making a total of 126 tests of samples performed. There was 100% accuracy both with the cassette and with photographs read by the tester and two additional readers. All readings were congruent and consistent. All were blinded for 9 testers with 3 testers in each of three settings (physician office, nurse health care setting, and laboratory conference room], and with testers differing professional backgrounds (3 nursing, 3 medicine in training, 3 licensed physicians in practice previously unskilled in use of such a test) (**Figure S4)**,

### Study 9. Use of ICT for Cincinnati epidemiology study between 2017 and 2019 demonstrates that ICT is an efficient way to perform such studies and that prevalence is low in Cincinnati

Of the 265 mothers tested, 8 (3%) had a positive IgG for *Toxoplasma* infection. None of these had a positive IgM. Variables of interest were available for 264 of the mothers including residential address (longitude and latitude), age, education, race, income and pet ownership as part of the original cohort study. There were no significant associations of testing positive for *Toxoplasma* infection and any of these variables (**Figure 5**).

### Study 10. Results with ICT AdBio test (Onsite POC) that detects anti-*T. gondii* IgM and IgG separately has both substantial false negatives and false positive IgG (9%) and false negative IgM(18.5 % true positives detected)

A total of 147 samples were tested using the Onsite POC test (**Figure 2D, Table S8A-C**) in Quindio, Colombia. By using *Toxoplasma*-IgG detection by VIDAS as reference test (**Figure 2D, Table S8A**), the sensitivity of Adbio for *Toxoplasma*-IgG detection was 100 % (95% confidence interval [CI], 94.4%–100%), and specificity for *Toxoplasma*-IgG detection was 90.8% (95% confidence interval [CI], 80.3%–96.2%). Of note, in 6 of 65 (9%) samples, the Onsite POC kit tested positive for IgG but were negative by VIDAS IgG. In 5 of these sera the bands observed in AdBio were visible. One serum with positive result for IgG in the AdBio but negative in the IgG VIDAS, was obtained from a boy with confirmed congenital toxoplasmosis who was being treated when the sample was obtained. For diagnostic accuracy of IgM detection, sensitivity for *Toxoplasma*-IgM detection was 18.5% (95% confidence interval [CI], 7.0%–38.7%; 5 of 27), and specificity for *Toxoplasma*-IgM detection was 100% (95% confidence interval [CI], 96.1%–100%; 120 of 120). Summary of the diagnostic properties found in this group of sera are shown in **Table S8C**, including NPV and PPV values.

## Summaries of Results

Considering the complexity of this decades long work that occurred in stepwise studies presented with detail in this Supplement, results/data are summarized. **Figure 6** summarizes the results of all the studies above verbally in the format that corresponds with the initial format of the Design and “Roadmap” shown in **Figure 1**. As noted above in the individual analyses of the studies with detailed results, **Figure 2** presents visual summaries of data in the results of the following **Studies**: **1** (**Figure 2A** USA feasibility Clinical Trial Study in accordance with FDA guidelines and predicate test), **2a** (**Figure 2B** Lyon False positives vs ICT), **2b Figure 2C** Paris false positives vs ICT, (**Figure 2D, Table** S**8** AdBio false negatives and positives)vs predicate test), (**Figure 2E, Table S7,** Quindio acute), **Study 6 (Figure 2F, Figure S4, QI-LOD**), **Table 3** summarizes all studies of ICT with **Table 2** that summarizes acute, IgM, in US with ICT. Others as presented separately, **Study 3 (Figure 4**, monthly feasibility acceptability, note Study 6 also addresses reliability of ICT vs other predicate tests in France and Quindio, **Table S6), Analyses 4 and 5 (Tables 1-7), Study 9 Epidemiologic study in Cincinnati (Figure 5)**.

## DISCUSSION

The results above (summarized in **Figure 6, Table 3** and also **Figure S8 in the Commentary below**) demonstrate that the ICT has proven effective at identifying sera and whole blood samples of USA and non-USA patients with known *T. gondii* infection. It detects seroconversion early in infection. It is also was effective at identifying the false positive test results for *T.gondii* specific IgM of other, currently FDA cleared tests of sera when no *T. gondii* specific IgG is present. It was well-accepted in a monthly screening program that was shown to be feasible in a USA academic obstetrical practice. It also functioned with high precision while meeting WHO ASSURED criteria even in whole blood samples at the limit of detection of specific anti-*Toxoplasma* antibody. It was found to be straightforward for physicians, nurses and medical students and a medical resident to easily learn to use the ICT and accurately interpret the ICT results using the Quick Information in simple written instructions (**Figures 2F, S4**).

Up through and including the current stages of the clinical feasibility trial at the University of Chicago Medical Center, diagnostic sensitivity has exceeded 99% and specificity has stayed at 100% with all samples of U.S. patients. In addition, across several of these studies, this ICT has outperformed other screening tests. Herein, out of 99 IgM false positive sample results, across multiple consecutive different USA and French sets of data recently there have not been false positives or false negatives. In addition, in two countries (the USA and Morocco [29]), the ICT has not had false positive or borderline bands when testing the serum and/or whole blood. While it was already known that this test could perform accurately this present work, (**Tables S1-3, S7**), also has evaluated the ability of ICT to correct the errors of other carefully tested, commercially available screening assays [27] using prospectively and retrospectively collected sera in the USA, France and Morocco. The high specificity is a particular strength for the ICT IgG-IgM device, especially when compared to other currently available commercial tests for anti-*Toxoplasma* IgM.

The data from Houze et al (ECMID and manuscript in preparation [30] and Mahinc et al [26] increases the number up to 137 of such false positives IgM studied with the ICT. Mahinc et al also studied 23 false positive Architect and/or Biorad Platelia IgM [26]. In the Mahinc study [26], false positive IgM in the Biorad test were obviated by ICT testing 21/23 of the time. In Tunisia [31], recent results were similar adding additional data but with a higher proportion of false positives [31]. Ten of 13 false positives were negative in the ICT. Although there were no ICT false positives in these data sets in the US, the occasional false positives (5 of 36) in the work earlier in Marseilles and Tunisia emphasize the importance of confirmatory testing of positive results. The high quality performance of some of the Reference tests emphasize that some tests seem to perform better than others when used in Reference laboratories (**Table S6**).

Our studies, along with the earlier experience in the Palo Alto reference laboratory and collated recent results, demonstrate practical problems in the US with potential serious consequences for patient care [35] where the ICT can be helpful in a patient’s management. This has been confirmed in France making a total of 132 of 137 for IgM) and 27 of 27 times for IgG times that a false positive result could be corrected. False negatives are uncommon but would be detected by repeat testing in gestational screening programs. Any positive ICT during gestation would have confirmatory testing to differentiate IgG and IgM. The occasional false positives would be detected by back up testing in the reference laboratory in the USA or use of multiple tests including the Western blot in France. Reference laboratory gold-standard testing and certain commercially available test reagents have higher performance than testing in local laboratories as shown in **Figure 2A-C, Tables 1-3, S6**. The ICT has high precision with samples at the limit of detection. That the test is easy for medical students, a medical resident, practicing board certified physicians, nurse/nurse practitioners without familiarity with the ICT to perform and interpret (**Figure S4**) is congruent with a recent experience with 30 practitioners in Armenia, Colombia [47]. This experience was with patients infected with genetically distinctive Colombian parasites [47]. Acceptability in a Colombian patient and obstetrical practitioner group was high [47], similar to acceptability in our experience presented herein in the USA.

Colombian sera also were tested in Colombia [47–50] with a different lateral chromatography test made in the USA called the ADBio. This test differentiates IgG and IgM and has a USA sale price more than ten times that expected to be applied for the ICT. Unfortunately, the performance of the ADBio test was problematic (**Fig 2D, Table S8 A-C**) when compared in the Quindio Reference laboratory with Vidas IgG and IgM reference tests [47–50].This is similar to our earlier results with this test with French (unpublished) and USA [27] sera. For the Colombian sera specifically, the difference of the ICT test for combined detection of IgG and IgM antibodies the AdBio test resulted in lower sensitivity for IgM in stored samples from a biobank. ICT combined simultaneous detection of IgG and IgM can improve sensitivity for IgM because most of the IgM sera used for sensitivity analysis already have IgG [26, 27, 30} and the mechanism of the test with antigen coating the black bead reacting with both quadra/pentavalent IgM and bivalent IgG which react with the antigen placed in the line on the nitrocellulose. This combined detection of different isotypes also contributes to better specificity. The lysate antigen used in the ICT contains many proteins. The Western blot can accurately discriminate between and recognize IgG and IgM specific for *T.gondii*, as can the combination of other tests such as the Sabin Feldman Dye test which also detects IgG and IgM and the double sandwhich IgM ELISA or the IgM ISAGA. The IgM ISAGA is more sensitive and thus preferable for use for infants.

In the context of clinical protocols for prenatal *Toxoplasma* screening, the ICT insures that far fewer “false alarms” are generated and that less time and resources are spent on confirmatory testing for a pregnant woman who shows an isolated positive *Toxoplasma* IgM test. Risk that such sample may be a false negative IgM from the ICT test is very low, but cannot be excluded. To avoid any risk the patient should be retested for IgG and IgM 2-3 weeks later to ensure that IgG did not appear. This happens as part of a systematic gestational monthly screening program (**Figures S2.I, 4, Table 4**). It should be emphasized again that POC tests for anti-*Toxoplasma* IgG and IgM, such as the ICT, are merely a first step toward diagnosis, given that IgM antibodies can persist for up to several years after acute infection. For any woman who receives a false positive IgM test result, the next step of an evaluation with other tests can involve weeks of waiting for a blood sample to be tested using technology that runs at much higher costs than the point of care test [1, 2, 6, 7, 27, 33, 35].

A potential limitation of the ICT might have been the lack of utility of the ICT using saliva (Peyron, unpublished). There is a nanogold Nirmidas test that was used with saliva, serum, and whole blood finding high sensitivity and specificity and dye test precision for the detection of IgG and IgM [37]. We had suggested earlier this might be an ideal test to use before conception. Although finger stick for glucose is standard, easy, and familiar in obstetrical practices, obtaining saliva may be viewed as less difficult than whole blood. Thus, some view saliva could be a potential advantage. However, the nanogold has required transport, associated delays to reach a clinical laboratory, and electricity and a sophisticated machine for testing. Recently manufacture of this nanogold test was discontinued. Nirmidas has also used a gold bead ICT for SARS CoVi2 but nothing like this has been produced for *Toxoplasma* to date. The diagnosis and management of *Toxoplasma* infection best involves knowledgeable health care provider input urgently making the advantage of home testing saliva less.

Testing before conception to identify seropositive persons and then testing regularly monthly through pregnancy for those who are IgG seronegative initially would be ideal as it helps to obviate problems of anxiety provoking delays that can result in irreversible fetal damage, as well as false positive test results. Such damage in congenital toxoplasmosis, as well as in ocular toxoplasmosis can occur in very short times of less than a week, making diagnosis and initiation of treatment urgent and emergent. Minimizing the likelihood of false positive IgM while maintaining maximum sensitivity is a top priority for any point of care test candidate.

The ICT also should be very useful in clinical laboratories testing with sera with a potential false positive IgM result without IgG as described herein. It could function as a second-line test to confirm or find IgM specific for *T. gondii* is not present before sending the sample to a reference center, while continuing to follow the patient while awaiting Reference laboratory results. This is a major advance as this will save time and reduce the need for gold standard tests. It can help reduce concern for patients and physicians. When the ICT test is used initially with whole blood the only predicate test for confirmation needed will be if the whole blood test is positive. ICT not only helped to obviate the problems with false positives but also can result in detection of true positives and very early seroconversion as described herein and also recently acquired infections described elsewhere [9, 23, 24, 26, 27, 30]. We placed this work in the context of ongoing problems for healthcare (**Table S9, Figures S7, S8**) and potential for direct and spillover benefit for the care of pregnant women and their families (,[14]). We also placed these studies 1 to 12 herein in a historical context building parts of a toolbox working toward a role of screening using WHO ASSURED criteria compatible tests in the elimination of congenital toxoplasmosis (**Figures S7, S8)**. There also are a variety of other clinical and epidemiologic circumstances where knowing *T.gondii* serologic antibody status can be of considerable clinical and public health utility and benefit [1, 2, 6, 7, 27, 32–9, 46–50]. Very high-quality, low-cost screening tests such as the ICT can improve infectious diseases care in gestation, help to eliminate perinatal infections with considerable spill over benefit for health care for pregnant women and in other clinical and research settings.

Congenital toxoplasmosis is a treatable and preventable disease, and physicians and other obstetrical providers now have the tools, in-hand, to improve outcomes and reduce patient and familial suffering. This screening, the standard of care in other countries, is now increasingly feasible in countries like the United States, where the primary argument against screening has been its economic burden. In the development of this test and other high-functioning point-of-care tests, there is potential for transformation in the provision of obstetrical care to improve maternal-child health. These benefits are amplified in subpopulation demographics in the USA[28] and regions of the world where the burden of disease is even higher. Examples of this occur in the Lancaster Amish population in the USA [8, 12, 32, 46], parts of Florida, are likely in other US subpopulations [28], and occur in Central [36] and South America [47], and parts of Africa [29].

Use of the ICT for the Cincinnati maternal cohort study found ICT to be efficient (**Study 9, Figure 5**). Due to the small proportion who were seropositive, we were unable to test for any clustering by known risk factors for exposure: none of the individual socio-economic or location factors in a regression analysis achieved statistical significance. Future analyses with a larger overall sample size will be needed to evaluate risk factors in this population. The reasons for the relatively low prevalence in Cincinnati in this cohort remain to be discovered. We have cared for children with congenital toxoplasmosis in Cincinnati, thus, even with the low prevalence found, it is likely still that gestational screening would be worthwhile.

Our recent study in Colombia also demonstrated high acceptability of a single use of the POC on a large scale of 783 women and 30 providers [47]. Although *Toxoplasma* infections occur in all demographics it was a particular problem in those who had lower education and socioeconomic status [31, 47]. To understand risk factors during gestation and develop programs to prevent such infection will require monthly screening in areas of high to low prevalence.

Implementation of this study in the clinical trial and QI limit of detection study demonstrated that it should be easy to introduce this test into obstetrical or other practice with little time or inconvenience. For example, when patients are evaluated for vital signs, blood pressure, glucose including by fingerstick, by medical assistant or nurse, cassettes can also be brought to obstetricians or other health care practitioner for additional reading and entry into medical records. Photography using an I phone for documentation could easily be included into processes for additional documentation made available to patient and records.

## COMMENTARY AND CONLUSIONS

### Where have we come from, where are we now, and where might we be in the future?

This **Commentary** addresses where approaches to prevention and treatment of congenital toxoplasmosis have been as the studies herein were developed and carried out and our goal that the studies herein provide a foundation to move beyond the current status of prevention and care in the US. We provide a Table and two figures to summarize and to provide historical context. We present cases representative of those that we encounter regularly (**Table S9**), an historical overview with a timeline (**Figure S7**) and a Figure showing what we encountered for many decades before carrying out the studies herein (**Figure S8**). Our goal in performing the studies described herein is to move beyond the harm we have observed from untreated congenital toxoplasmosis.

In France screening was mandated by law. In Austria those screened received additional health care benefits. In Colombia it was introduced through practice societies. In the USA those in advisory positions recommended that education, easy feasibility, low cost would result in those who would benefit choosing to have testing incorporated in medical practice and USA patient culture at many levels by personal preference. The acceptability study demonstrated that informed patients would want this and obstetricians could use this comfortably and without inconvenience in their practice. It could easily be introduced into family practice and adolescent pediatric care to identify seropositive patients at risk of this most common form of retina disease due to infection and loss of sight. Such screening in adolescence could also provide pre-pregnancy testing for young women to allow knowledge of who is seronegative and should be screened during pregnancy. Pre-marital/conception screening as initially occurred in France could also be helpful as families plan to have children. As *Toxoplasma* has been transmitted by organ donation and white blood cell transfusion and by sperm in domestic non-human animals, and can relapse with immune suppression and may be causative for epilepsy, and some neurodegeneration, has caused epidemics in North, Central, and South America, and has been categorized by NIH as a potential bioterrorism pathogen, and a neglected tropical disease, there are a number of other medical settings where knowledge of *Toxoplasma* serologic status may be useful.

Obstetricians, nurse midwives, family practitioners, obstetrical nurses, and other obstetrical providers are uniquely positioned to intervene to prevent this disease, to improve the health of both mother and child. POC test-based monthly gestational screening of seronegative patients for *T. gondii* infection provides a valuable tool in the obstetric armamentarium to ensure maternal-child wellness. When such tests have undergone appropriate evaluation by the FDA and CLIA, as they have undergone in the CE-mark evaluation and approval in Europe, this testing can enable a paradigm shift in our management of the risks associated with exposure to *T. gondii*.

### Funding, Acknowledgements, Disclosures and Insuring Objectivity in Results

LDBio Diagnostics provided the ICT and Western Blots used in the studies. ANNAR Labs (Colombia) donated the AdBio kits. For the predicate test, costs for the comparison test for 58 persons for the feasibility, clinical trial the cost of performing the Biorad IgG and IgM tests was provided by the Susan and Richard family Kiphardt Seed Fund and The Thrasher Children’s Charity. At LDBio Diagnostics, Denis Limonne Pharm D. is the scientist and CEO share holder and Raphael Piarroux PharmD, PhD was the R&D Director Scientist until January 13, 2023. A patent application was submitted by D. Limonne with the scientists at the University of Chicago and in France in August 2018. This application is pending review in the United States in accordance with US Bayh Dole laws. This is for the development of the whole blood point of care test and the practical clinical utility of the ICT to guide treatment for gestational infection to prevent congenital toxoplasmosis. This is to insure its continued high-quality performance and reproducibility of the results described herein. It is pending in review at the US patent office.

In this collaborative work, the scientists D. Limonne and R. Piarroux (DL.RP) at LDBio provided insights and knowledge from their earlier work in creating the ICT, and collaboratively with RMc discussed FDA and CLIA requirements with RMc and the FDA during an IDE and “presubQ” phase of this study. In this phase, the FDA Program provided guidance for this academic/Biotek collaboration to prepare materials to allow FDA review for dual 510K clearance and CLIA waiver for use of the ICT in the USA. RP of LDBio performed the analysis of the French Blood bank serum to establish that the correct dilution required by CLIA instructions was 1:89. DL and RP designed the instruction sheet with input from FDA, CLIA, and RMc to be tested in the “QI at limits of detection study”. This was perfected in the “presubQ” process with advice from the FDA and CLIA as the FDA indicated that a 510K clearance and dual CLIA waiver might be the appropriate application mechanism. The scientists at LDBio did not interfere with the performing of the tests, the recording, interpretation of the results nor the reported conclusions of any work at any academic site. All these studies were performed independently in the academic centers. There was no payment to the scientists. At Hôpital Bichat, Paris and in Morocco studies were/are being reported separately. LDBio did provide resources to support operating expenses and reagents, but not in the USA or Colombia. RP and DL participated in editing initial and final drafts of the manuscript. The ICT tests of the Cincinnati samples and three western blots for Chicago samples were performed at LDBio. We gratefully acknowledge all participants in this work and those at the FDA who worked with us in the “Pre-Sub Q process” recognizing the substantial potential humanitarian benefit of the work toward obtaining FDA clearance and CLIA waiver that could allow this work and test to be used to help people and prevent suffering and loss of life, sight, cognition, and motor function, while saving costs for health care.

Additional thanks for funding from the Medical Student Award, the National Institute of Diabetes and Digestive and Kidney Diseases for their Grant #T35DK062719-30, the National Institutes of Health for their Division of Microbiology and Infectious Diseases Grant to RMc #R01 AI2753,RO1 16945, AI08749-01A1 BIOL-3, U01 AI77887, U01 AI082180, TMP R01-AI071319, the Thrasher Children’s Charity Research Fund for their E.W. “Al” Thrasher Award, the Kiphart Global-Local Health Seed Fund Award (to RMc), University of Chicago. We are grateful to Taking out Toxo, Network for Good, Toxoplasmosis Research Institute, The Cornwell Mann Family Foundation, The Rodriguez family, The Samuel family and Running for Fin, the Morel, Rooney, Mussalami, Kapnick, Taub, Engel, Harris, Drago,Longfellow/Van Dusen families, and the study participants. We thank testers E.McLeod, C.Weber, F.Goldenberg, C. Guinnette, R. Tennant, Z. Williams, H.Taylor, A. Beem, and S.Syed and photographers E.McLeod and K El Bissati for their reading and photography in the limit of detection, QI study. We thank all the participants in these studies of the ICT and those in the IRB office, members on the IRB, Office of Clinical Research, at the FDA and CLIA for their guidance and A. Ponsler at the Thrasher Foundation for emphasizing the importance of these studies on reviewing the initial data at a site visit at Stanford University and the Remington Serology laboratory with RMc.

## Data Availability

All data are included in the manuscript and its supplement.

---

**Figure S1A-E.**
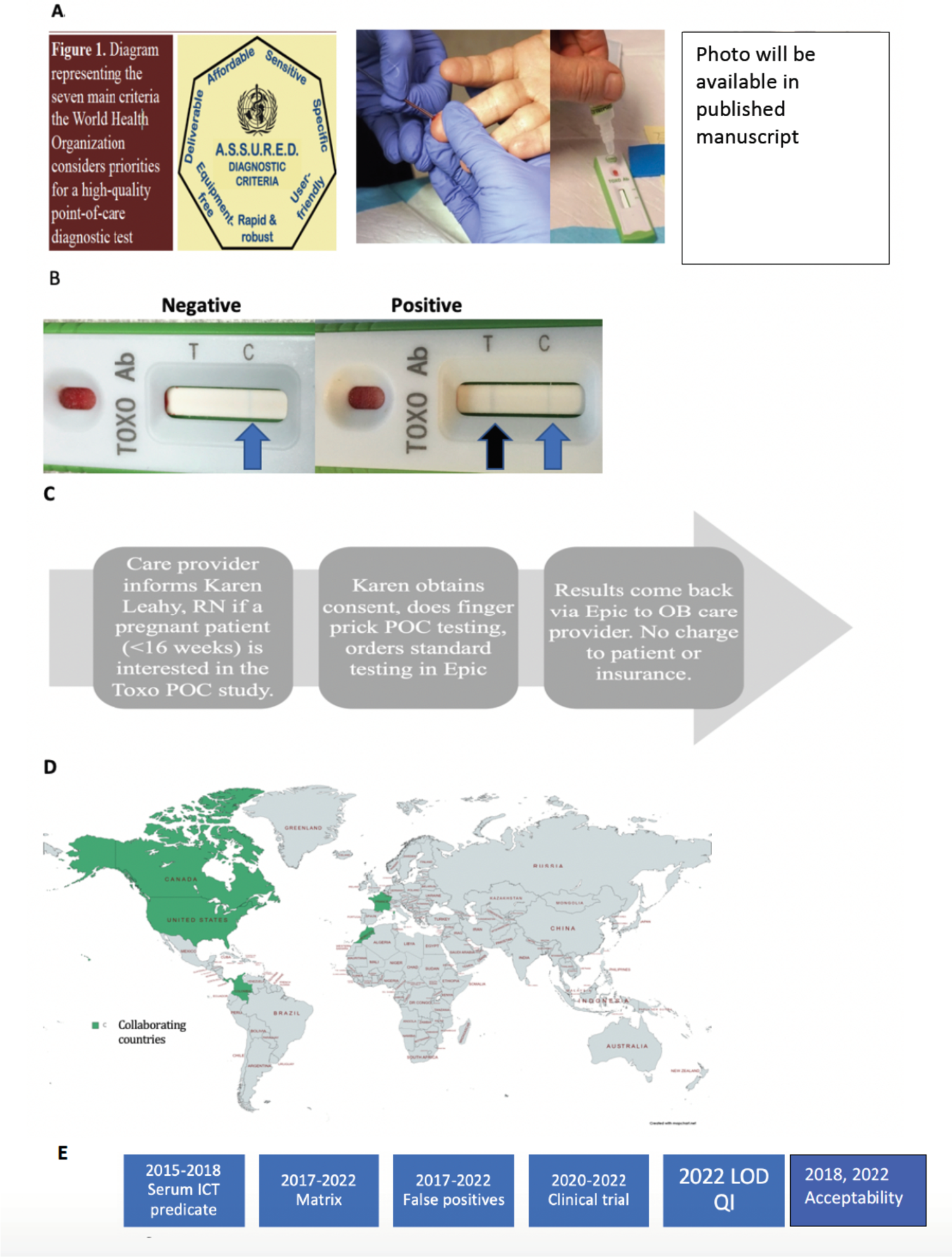
Performing *Toxoplasma* ICT IgM-IgG. A, B. Testing and Reading ICT. Finger prick and blood collection, dispensing blood, then 4 drops of buffer, waiting 20 minute to read as shown in B, Blue arrow indicates blue control band and black arrow indicates black *Toxoplasma* antibody band. Review can occur with patient in twenty to thirty minutes as shown. C. Design of clinical trial operationally in the clinical setting. Sample then goes to clinical lab with request within the EPIC system. Results are available to the ordering physician and available in patient’s chart. D. Areas of the world that this ICT has been tested referred to in this manuscript. E. Present acceptance studies in the context of earlier work chronologically. (Some images in A,B adapted from Easton J, U Chicago Press office. With permission.)

**Figure 2.**
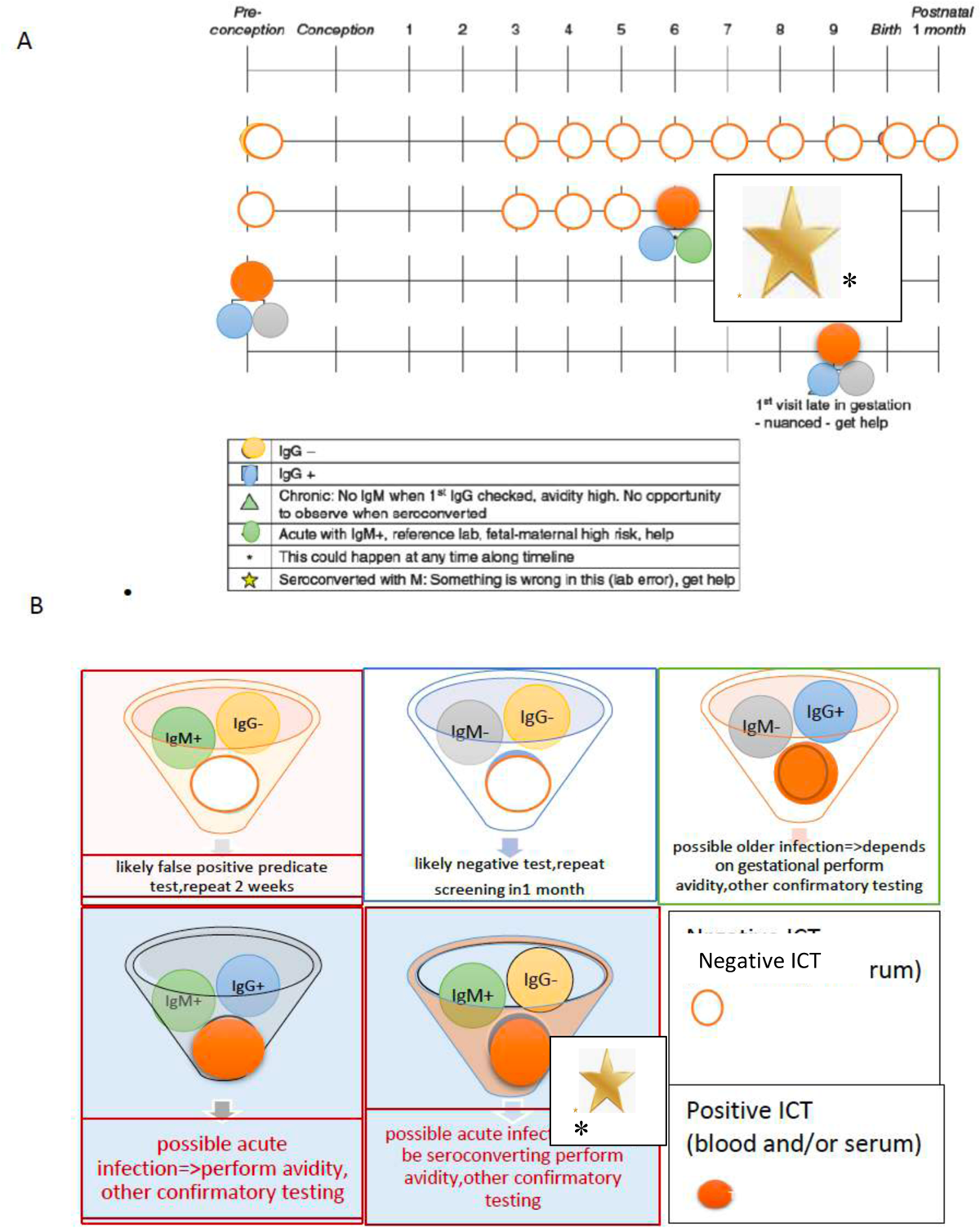
I Approach to detection of seroconversion using monthly screening, also see Table 4, S7A, Figure 4

**Figure S4.**
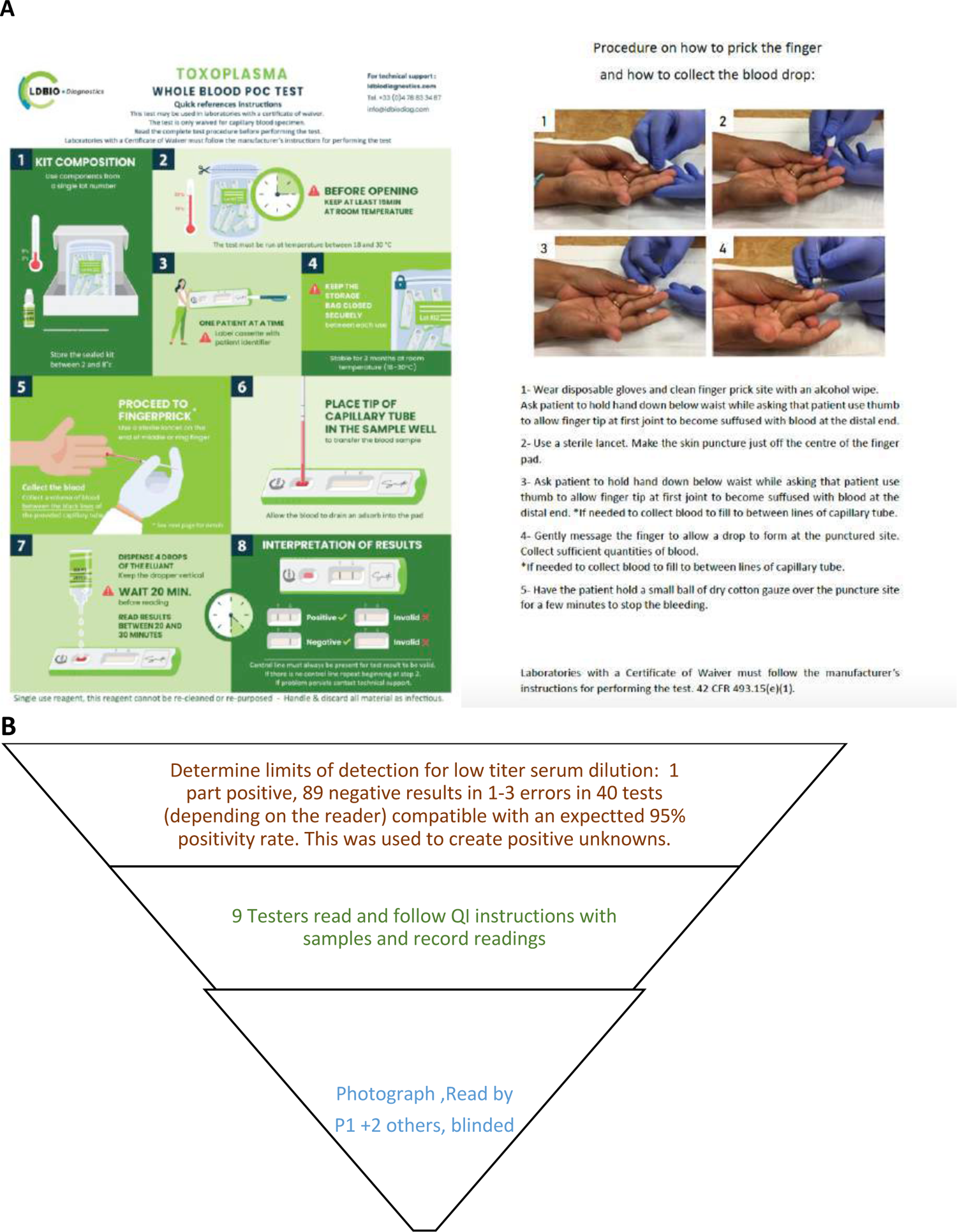
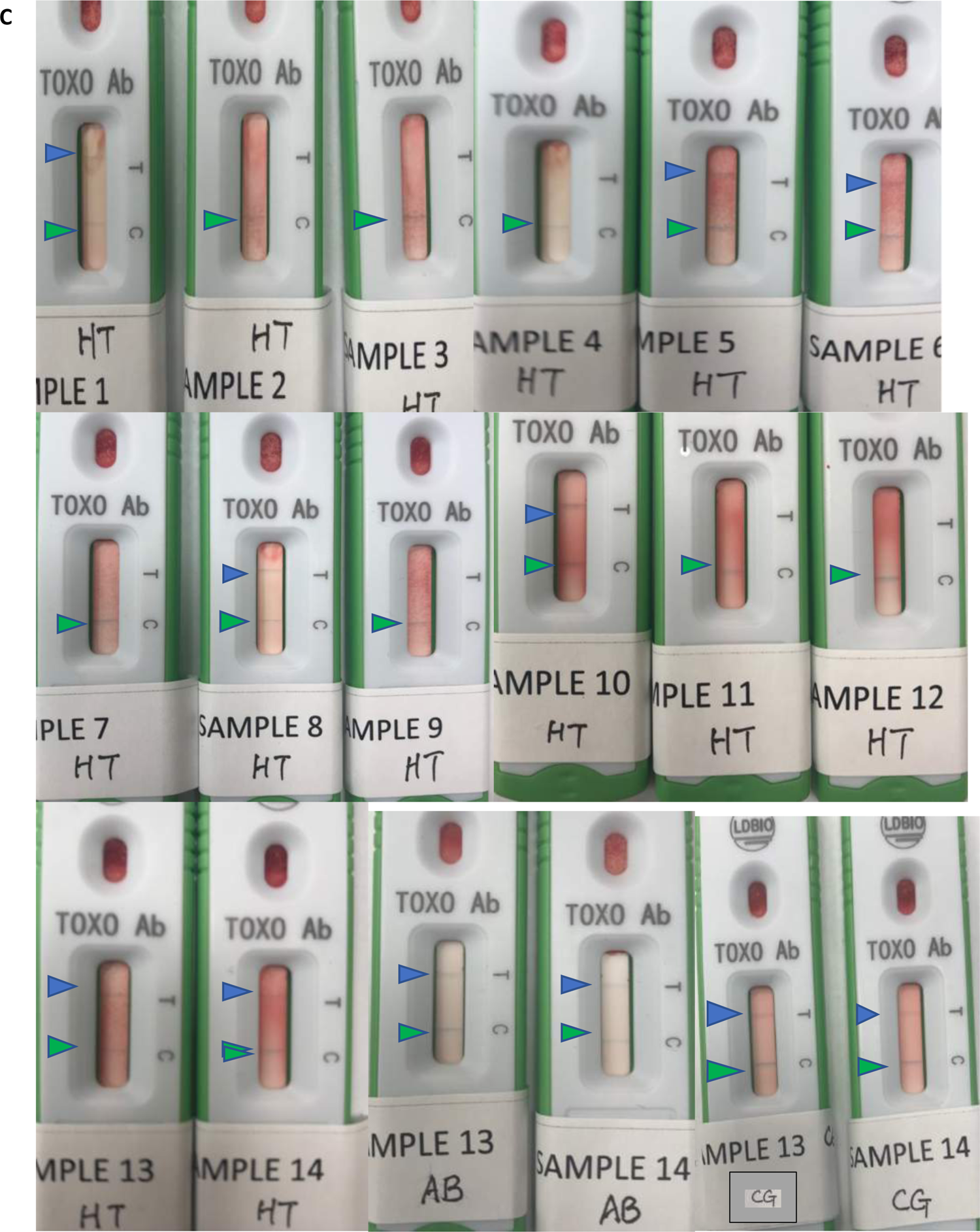
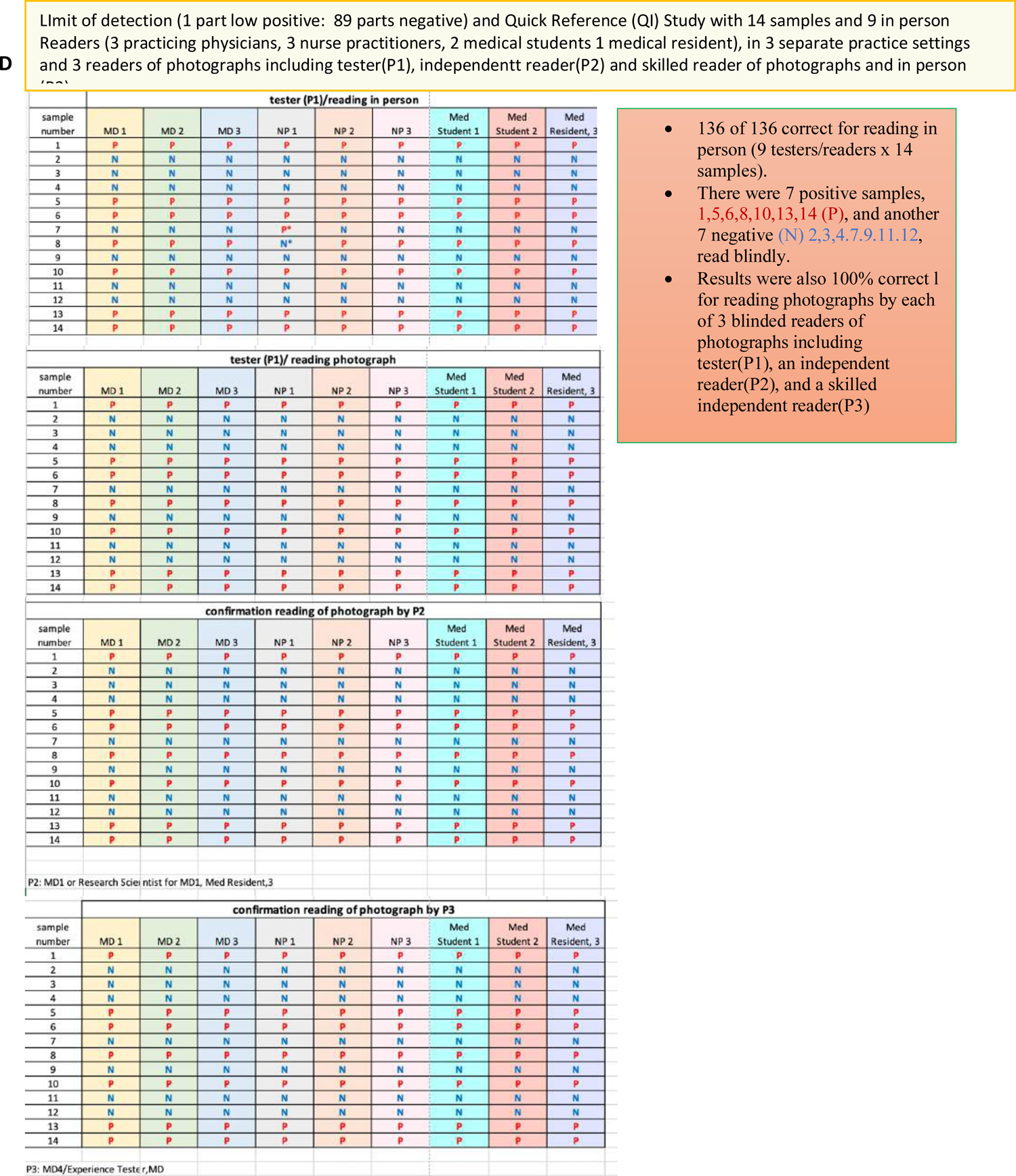
Limits of detection and quick reference study. A. Instructional material provide called Quick Information (QI). **B. Design of study**. **C**. **Representative examples of casettes.** Tester HT, samples 1 to 14, Tester AB samples, 13,14; Tester CG, samples 13,14. **D. Summary of readings** of results at the time of test by readers performing the test, and another reader, and of photographs by each reader and two additional readers. All readings were blinded as to whether the samples were positive or negative and the code for the samples. Concordance of in person readers and reading of the photographs by all readers blinded and independent of one another was 100%. Also shown as panel in summary **Figure 2F**

**Figure S7.**
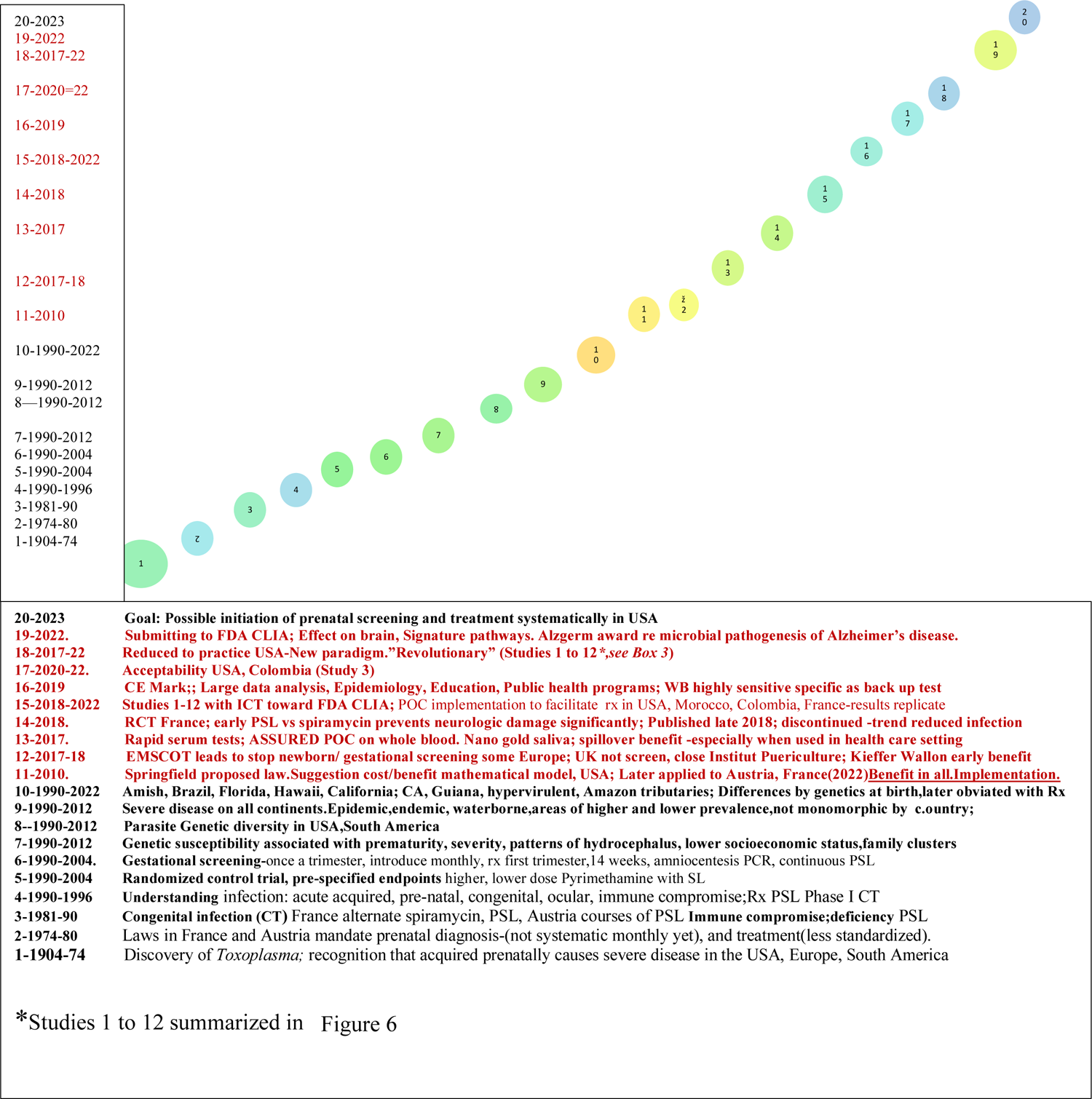
Overview Summary of studies herein in context of other work. Top presents context of current studies toward introducing screening in a global initiative. Red font shows work herein. Figure S7 summarizes the studies herein.

**Figure S8:**
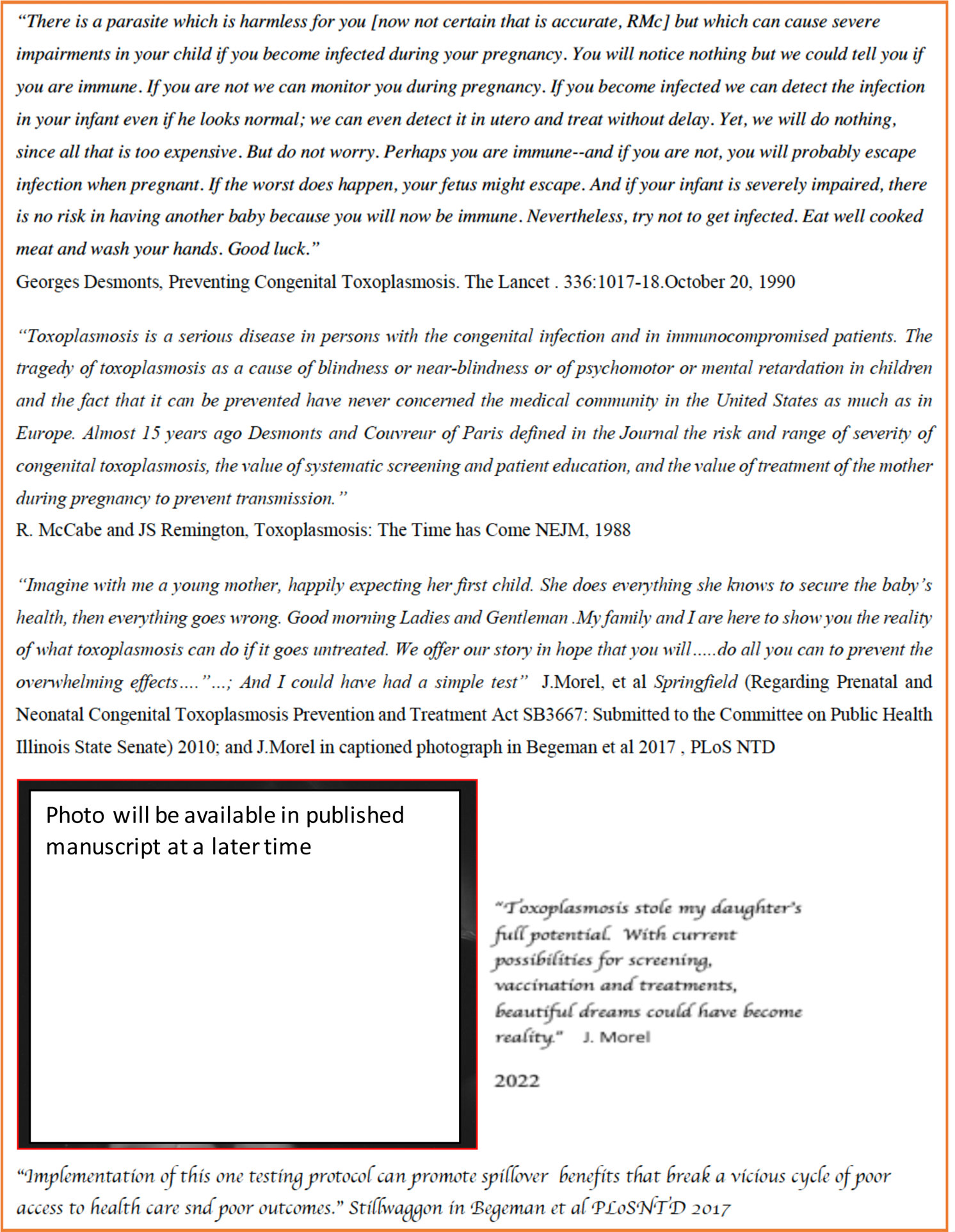
Historical perspectives on screening and treatment of *Toxoplasma gondii* acquired in Gestation in France and the USA. This figure is to provide context of where we have come from to studies herein with considerable spillover benefit for patient care and well-being, with the goal that studies herein will provide a foundation for improvement of prevention and care for congenital toxoplasmosis.

**Table S1.**
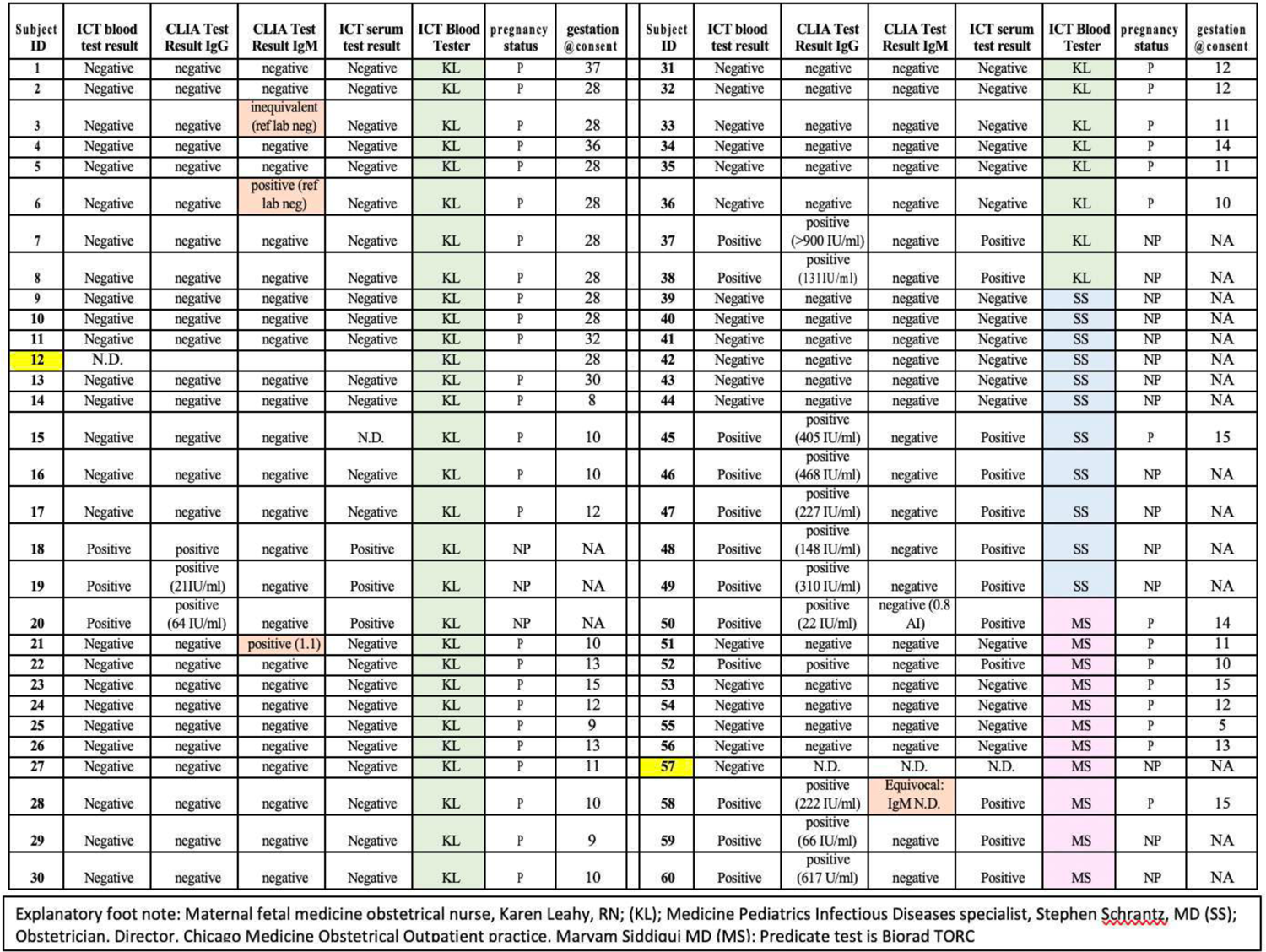
Chicago Clinical Feasibility Trial 2020-2021.

**Table S6.**
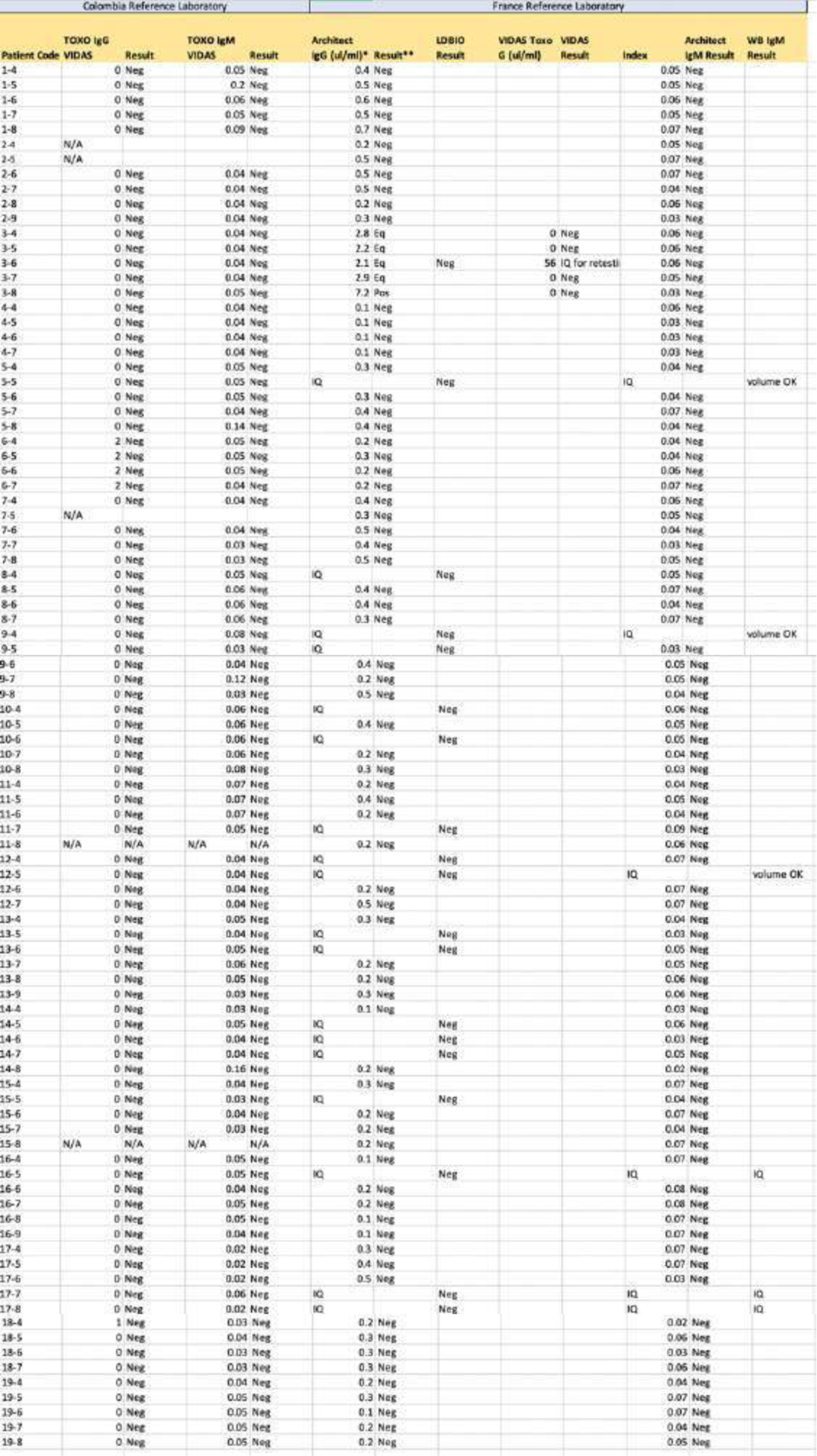
Analysis of 164 ICT negative samples with Other tests in Colombia and French reference laboratories

**Table S7.**
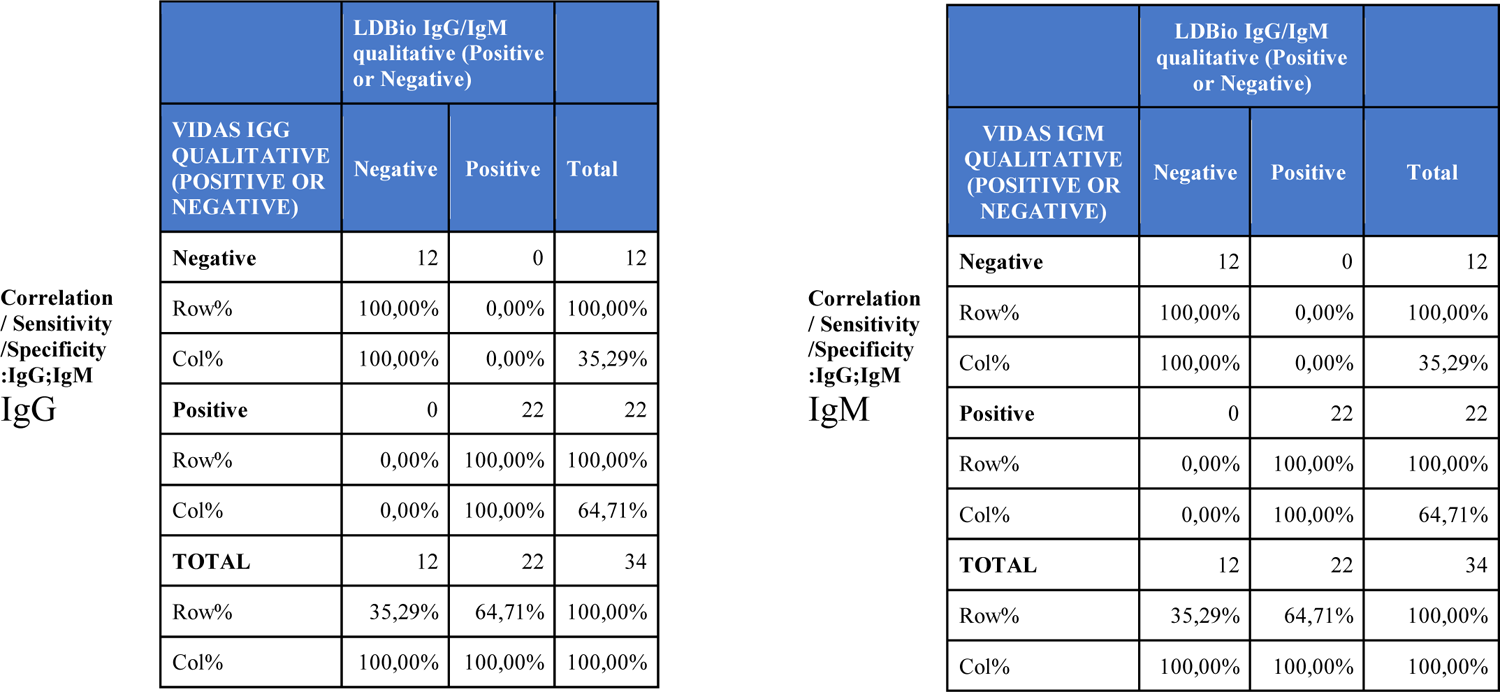
Correlation between positive IgG and IgM and seronegative samples in Colombian patients measured in Vidas and ICT results in these patients

**Table S8.** Determining whether the USA manufactured ADBio could perform as well as ICT with Colombia samples or whether it would introduce the same problem of false negative IgM results as found in an earlier study.

|  | VIDAS IgG result |  |  |
| --- | --- | --- | --- |
| AdBio IgG Result | Positive | Negative | Total |
| Positive | 82 | 6 | 88 |
| Negative | 0 | 59 | 59 |
| TOTAL | 82 | 65 | 147 |

|  | VIDAS IgM result |  |  |
| --- | --- | --- | --- |
| AdBio IgM Result | Positive | Negative | Total |
| Positive | 5 | 0 | 5 |
| Negative | 22 | 120 | 142 |
| TOTAL | 27 | 120 | 147 |

C. Diagnostic properties for *Toxoplasma* IgG and IgM detection of AdBio POC Test compared to the result of VIDAS
| POC Kit | Toxoplasma Antibodies | Sensitivity (%) (95% CI) | Specificity (%) (95% CI) | NPV (%) (95% CI) | PPV (%) (95% CI) |
| --- | --- | --- | --- | --- | --- |
| Onsite | IgG | 100 (94.4-100) | 90.8 (80.3-96.2) | 100 (92.4-100) | 93.2 (85.2-97.2) |
|  | IgM | 18.5 (7.0-38.7) | 100 (96.1-100) | 84.5 (77.3-89.8) | 100 (46.3-100) |

**Table S9.**
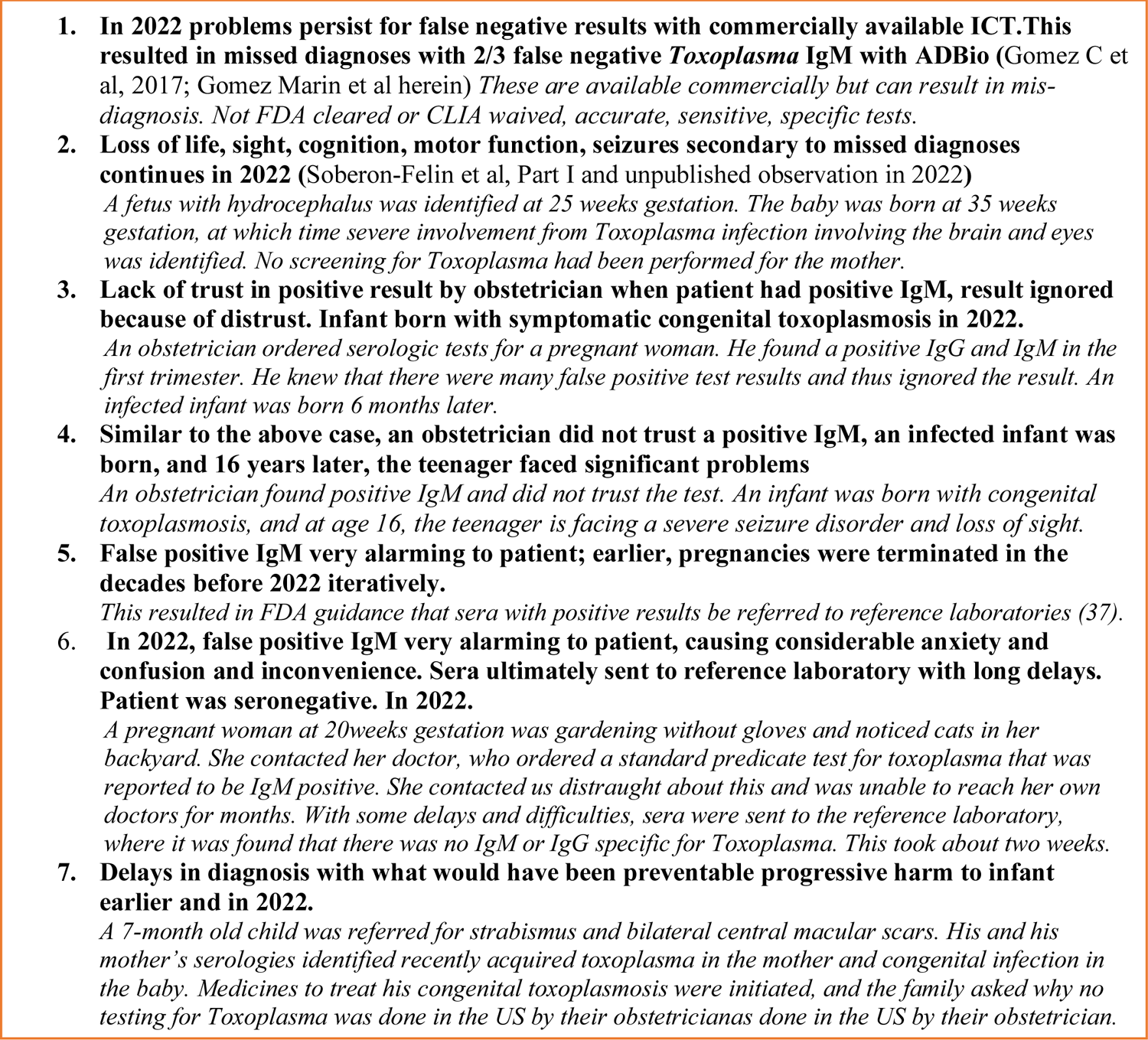
Case vignettes provide representative practical examples from false negative and false positive *Toxoplasma gondii* IgM tests in the USA that harm patients and patient care

